# Measuring High-Priority Outcomes in Autistic Adults: Initial psychometric assessment of the instruments in the AASPIRE Measurement Toolkit

**DOI:** 10.64898/2026.03.23.26349108

**Authors:** Christina Nicolaidis, Dora M. Raymaker, Mary Baker-Ericzén, Shannon des Roches Rosa, Todd Edwards, Emanuel Frowner, Willi Horner-Johnson, Andrea Joyce, Steven K. Kapp, Clarissa Kripke, Julie Lounds Taylor, Julia Love, Rachel Ludwig-Kripke, Joelle Maslak, Katherine McDonald, Ian Moura, Mirah Scharer, Zack Siddeek, Ivanova Smith, Joseph Vera, Anna Wallington, Liu-Qin Yang, KJ Flores, N Chaiyachakorn

## Abstract

**Background:** Adult autism services research is hampered by a lack of accessible self-reported outcome measures. The AASPIRE Outcomes Project used a community-based participatory research (CBPR) approach to create and test the AASPIRE Measurement Toolkit, a set of accessible survey instruments for use in real-world settings. The core toolkit contains 12 characteristics modules and 19 outcome measures, each with self-reported and caregiver-reported versions.

**Methods:** We used our CBPR-nested Delphi process, our collaborative instrument adaptation/creation process, and cognitive interviews to ensure accessibility and content validity. We then conducted a longitudinal survey to validate the outcome measures in a pragmatic sample of 870 autistic adults from two healthcare systems, two disability service systems, and the larger autistic community in the United States. Participants completed surveys at 3 time points (baseline and follow-ups at 5-11 months and 12-18 months). A 15% random subset completed an additional retest survey. We assessed 1) accessibility using completion rates and perceived ease of use; 2) internal consistency using Cronbach’s alphas; 3) convergent validity using Pearson’s correlations; 4) two-week test-retest reliability using intraclass correlation coefficients; and 5) six-month responsiveness to change by comparing self-perceived change with change in scores.

**Results:** Over 90% of participants reported the survey items were easy to understand. The outcome measures and their pre-determined subscales demonstrated strong content validity, internal consistency reliability, test-retest reliability, convergent and discriminant validity, and responsiveness to change.

**Conclusion:** The study provides strong evidence supporting the psychometric properties of the direct report versions of 19 outcome measures in the AASPIRE Measurement Toolkit. We will report additional in-depth assessments separately for each measure. Further research is needed to validate the caregiver report versions. All instruments are available for free and can help clinicians, service providers, advocacy organizations, and researchers assess the effectiveness of interventions and follow changes in outcomes over time.

**Community Brief:** *Why is this an important issue?:* Autistic adults have worse health and social outcomes than the general population. To learn how to improve outcomes, we need an accurate way to measure them. Most existing survey measures are hard for autistic people to use, not tested with autistic people, or can’t measure if the outcome has changed.

*What was the purpose of this study?:* The AASPIRE Measurement Toolkit is a set of accessible questionnaires (also known as “survey instruments”) that can be used to measure the outcomes most important to autistic adults. The purpose of this study was to test how well these survey instruments work.

*What did the researchers do?:* We used a community-based participatory research (CBPR) approach, where scientists and autistic community members worked together as equal partners. We collaboratively selected, adapted, and created the instruments in the Measurement Toolkit. We made a version for autistic people and a version for caregivers. The 19 survey instruments that measured outcomes focused on quality of life, health outcomes (overall health, flourishing, depression, anxiety, autistic burnout), social outcomes (community participation, self-determination, choices and decisions, employment), and supports and services (social support, barriers to communication, healthcare services, disability services, and adequacy of supports and services). Twelve other survey instruments asked for additional information describing participants. We used these 31 instruments with a diverse group of 870 autistic adults and caregivers. They took the survey three times over about a year. We used statistics and feedback to test how well the instruments worked.

*What were the results and conclusions of the study?:* The survey instruments tested very well:

1) They were accessible. Most people answered all the survey questions and said it was easy to use.
2) They were “internally consistent.” That means that people gave similar responses to different questions about the same idea.
3) They had good “test-retest reliability.” That means that, as expected, responses didn’t change much over a two-week period.
4) They had good “convergent validity.” That means that they related to each other in expected ways.
5) They were “responsive to change.” That means that changes in scores over 6 months matched participants’ own perceptions of change.

Researchers and service providers should use our survey instruments. We feel they work well because autistic people were involved at every step.

*What is new or controversial about these findings?:* Previously, there weren’t studies to see if survey instruments measuring these outcomes worked for autistic adults and could detect change. This study furthers what we know about adapting and co-creating accessible, accurate survey instruments for autistic adults using CBPR.

*What are potential weaknesses in the study?:* Not enough caregivers took part to test their version of the survey instruments. We think these instruments would work for other people too, but we only tested them with autistic adults.

*How will these findings help autistic adults now or in the future?:* These survey instruments will help researchers and service providers know if what they are doing to improve outcomes is working. That will help them create services that can do a better job improving autistic peoples’ lives.

## Introduction

Research suggests that, compared to the general population, autistic adults have worse quality of healthcare services^1–7^ and widespread gaps in social services,^8–11^ with worse health and social outcomes,^12^ including in mental health,^13,14^ physical health,^15–17^ educational attainment^18–20^, employment,^21–23^ and community participation.^24,25^ and increased mortality.^26–29^ While there are many calls for improved services,^30–37^ we still do not know which services and interventions are effective in improving outcomes.^38,39^

Studying the effectiveness of services or interventions to improve outcomes may require different tools and methods than those to document disparities. Intervention effectiveness studies often rely on data collected using survey instruments. However, few such instruments have been designed for or rigorously tested in populations of autistic adults.^40^ Psychometric data from studies testing survey instruments in the general population may not apply to autistic adults, including but not limited to those with pragmatic language challenges, intellectual disability, or other characteristics that make it difficult to respond to surveys.^40^ Autistic adults find many existing instruments inaccessible, offensive, or anxiety-provoking; or report that they cause them to provide inaccurate responses, skip items, or stop participating in studies.^41^ The lack of accessible, reliable and valid data collection instruments jeopardizes the validity and generalizability of results from studies using surveys of autistic adults.^40^ It also may contribute to over-reliance on proxy reporting, which raises validity concerns and ethical implications around excluding autistic voices.^42,43^

Even where validated instruments exist, many have only been tested in cross-sectional samples^44^ or were not designed to assess changes over time. Thus we may not know if they can adequately assess the effectiveness of interventions or accurately identify predictors of change over time.^45,46^ For example, many of the most widely used, well-validated instruments to measure quality of life or physical functioning in general populations may have poor responsiveness to change.^45^

Self-reported instruments specifically designed to measure outcomes—often referred to as patient-reported outcome measures (PROMs) or “outcome measures”—are critical for evaluating individual and system-level interventions and services, particularly when taking person-centered approaches. Consensus panels have established standards for developing and assessing PROMs ^47^ and initiatives like the Patient Reported Outcomes Measurement Information System (PROMIS) ^48^ and NIH Toolbox® ^49^ have produced comprehensive PROM packages. While many PROMs measure the health and social outcomes that are important to the autistic community, few are validated for use with autistic adults.^40^

### The Current Study

The Academic Autism Spectrum Partnership in Research and Education (AASPIRE) Outcomes Project is community-based participatory research (CBPR) project which aims to create and validate the AASPIRE Measurement Toolkit - a set of accessible, psychometrically sound outcome measures - and to explore changes in outcomes over time. Our academic and community partners co-created all instruments in the Toolkit, which are available in one version for autistic adults (with or without support) and another for caregivers when direct data collection is not possible, even with accommodations and supports. The project began in 2020; we hope to collect and analyze data from additional time points indefinitely. All instruments are freely available for use in clinical or research settings (http://aaspire.org/measurement).

The project had three phases. In the first phase, we used a CBPR-nested Delphi process with 53 autistic community leaders, representatives of family-focused advocacy organizations, service providers, and academics to identify the highest priority outcomes to measure when assessing the effectiveness of services for autistic adults.^50^ Panelists and team members also reviewed existing instruments to assess their potential use with autistic adults or their caregivers. In the second phase, we used our CBPR adaptation process^41^ to adapt or create 19 outcome measures. We then used an iterative cognitive interview process with 37 autistic adults and caregivers to further refine these instruments. Measures focus on quality of life, health outcomes (overall health, flourishing, depression, anxiety, and autistic burnout), social outcomes (community participation, self-determination, choices and decisions, overall employment satisfaction, and organization-specific job satisfaction), and quality of supports and services (social support, barriers to communication, unmet healthcare needs, barriers to healthcare, patient-provider communication, healthcare self-efficacy, satisfaction with disability services, and adequacy of services and supports). We also co-created or adapted 12 modules to assess a variety of demographic, social, health, and disability-related characteristics. We present detailed information about how and why we adapted or created the instruments separately (in preparation).

In the third phase, we conducted a longitudinal cohort study whose goals were to psychometrically test the instruments in the AASPIRE Measurement Toolkit and explore potential predictors of change in outcomes over time. Data collection for the cohort study took place between April 2023 and December 2024. We have presented a detailed description of our longitudinal study methods and the characteristics of our sample separately.^51^

The aim of *this paper* is to assess the initial psychometric characteristics of the outcome measures we collaboratively created or adapted. This information can allow researchers, clinicians, or service providers to decide whether to use the instruments in their own research or practice.

Separate papers will describe in-depth assessments of the construct validity, structural validity, and measurement invariance for each measure (e.g., Choices and Decisions,^52^ Autistic Burnout^53^) and identify predictors of change over time.

## Methods

### Community Academic Partnership and Participatory Approach

AASPIRE (www.aaspire.org) has used a CBPR approach with the autistic community since 2006.^54^ You can see example materials and detailed descriptions of our collaboration guidelines,^55^ methods, and processes in the AASPIRE Inclusion Toolkit (https://aaspire.org/inclusion-toolkit/).

The Outcomes Project team consists of a non-autistic, neurodivergent principal investigator (PI); an autistic community project lead; 10 autistic community partners, including non-speaking autistic people, people with intellectual disability, and people with varying levels of support needs; a leader from the parent advocacy community; a representative from a partnering governmental developmental disability services agencies; and other autistic, neurodivergent, and neurotypical co-investigators, consultants, students, trainees, and research staff from multiple universities. Greater detail on our Outcomes Project team exists elsewhere.^50^ Community members took part in all phases of the research project. The full Outcomes Project team made all high-level decisions together using a consensus process.

### Instrument Selection, Adaptation, or Creation

We used results from our CBPR-nested Delphi process to prioritize which constructs to include in the study, identify existing instruments measuring the selected constructs, and begin understanding concerns with existing instruments.^50^ We then spent two years adapting or creating instruments, conducting cognitive interviews and refining instruments, and assembling the full survey for the longitudinal study.

We created small workgroups, each of which included the PI and at least one autistic team member, to focus on each construct. Workgroups conducted literature searches, reviewed instrument-specific data from the CBPR-nested Delphi process, and made their own detailed assessment of existing measures. If possible, they selected an existing instrument and adapted it to address the identified concerns. Once they created an initial draft, they presented it in a full-team meeting. Team members identified persistent or new problems and offered suggestions for further improvements. In many cases, the team was able to reach consensus on the adaptations during the meeting. If they couldn’t, the workgroup continued adapting. The full team typically reached consensus on the additional adaptations via email; otherwise, we held additional full-team meetings.

While we intended to adapt existing instruments, we sometimes could not identify one to adapt, or we could not reach consensus on adaptations that would address all the group’s concerns. As such, we created four instruments and one additional subscale de novo. For each one, the workgroup created an initial draft based on Delphi panelist comments, literature review, and lived experience. We then used the same iterative process as above for revisions.

Once the team reached consensus on the direct report (DR) instruments, the caregiver report (CR) workgroup (including a parent of an autistic adult who would need to participate via CR) created CR versions of each instrument. They changed items to use 3^rd^-person pronouns for the autistic participant and 1^st^ or 2^nd^-person pronouns for the caregiver. The workgroup added qualifiers such as, “I think that they are satisfied with…” to items when they felt a caregiver could not fully know the autistic adult’s internal state; and they added items related to the caregiver’s confidence in their answer and how they know the information. We sought consultation from a non-speaking autistic adult who communicates independently via augmentative and alternative communication (AAC) but could remember the time before she obtained a robust form of communication as a young adult. She reviewed the package of instruments and suggested minor edits

Another workgroup then conducted 37 cognitive interviews (with 31 autistic adults, 5 caregivers, and 1 pair). Cognitive interviews are a structured qualitative method to evaluate and strengthen content validity. Participants review items and respond to standardized probes evaluating comprehension, relevance, and comprehensiveness.^56^ They are particularly important in the development of outcome measures for autistic adults.^40^ We conducted interviews via Zoom, focusing on 4-10 instruments per participant to allow for in-depth discussion within 60-120 minutes. The interviewer shared items via screen and used concurrent verbal probing, asking participants to paraphrase what they felt items were asking, explain why they answered the way they did, note anything that they found confusing, discuss the importance of the instrument’s topic, assess if important concepts were missing, and offer ways to make instruments clearer. (See Supplement A1 for interview guide.) We used an iterative process whereby we would make any necessary edits and show the revised version to the next participant. Once we refined all instruments, we brought them back to the full team, who came to consensus on the final versions.

All instruments use Plain Language and include a variety of accessibility features. A detailed description of the concerns with each instrument, the specific adaptations we made, and the accessibility strategies we used is available at www.aaspire.org/measurement.

### Longitudinal Study Procedures

The Institutional Review Board (IRB) at Portland State University (PSU) approved the project and served as the single IRB (protocol # 227674-18). The IRBs for our other study sites ceded review to the PSU IRB. Details about our accessible informed consent and authorization process^57^ and data sharing processes^58^ are available elsewhere.

We recruited three subcohorts to increase the heterogeneity of our sample and to enable us to validate the measures in different real-world settings. We recruited the Healthcare (HC) subcohort from Oregon Health & Science University (OHSU) in Portland, Oregon and Vanderbilt University Medical Center (VUMC) in Nashville, Tennessee; we recruited the Disability Services (DS) subcohort from two governmental disability services systems in Multnomah County, Oregon, and San Diego, California; and we recruited the Community subcohort from the broader U.S. autism community.

Participants had to identify as autistic, be at least 18 years old, and reside in the United States. HC and DS subcohort participants also needed a diagnosis of autism in their medical record or service records, respectively, and had to have received healthcare or disability services from one of the partnering systems in the past three years.

We recruited participants separately from each of the five study sites, matching recruitment methods to site needs. The HC sites identified eligible participants via electronic medical records and one of the DS sites identified them through a centralized database. They then invited them to take part in the study, oversampling people of color. The other DS site could not systematically identify potential participants, but offered information about the study to people receiving their services. We recruited the Community subcohort via social media, community organizations, presentations, our team’s website, and word of mouth.

Autistic participants could take part in the study directly with or without support, or via a caregiver proxy in situations where direct participation was not possible. We used an electronic survey system for screening, consent, data collection, and participant management. We used an extensive protocol to identify and prevent fraudulent activity.

We present a significantly more detailed description of the methods (including procedures for recruitment, screening, consent, data collection, fraud prevention, and data integrity) elsewhere.^51^

### Data Collection

We collected data at three time points. Participants took the second survey 5-11 months after baseline and the third survey 12-18 months after baseline. A random 15% sample of participants (N=132, oversampled for those who took part via a caregiver) re-took the survey approximately two weeks after Time 2.

Participants could choose to complete surveys online or meet with a research assistant over the telephone, videoconference, or (where feasible) in person. Participants could also get help from their own supporters. We collected and managed data using REDCap (Research Electronic Data Capture) electronic data capture tools hosted at OHSU.^59,60^ The REDCap survey allowed us to include multiple accessibility features.

**Table 1** describes the 19 outcome measures, including their origins, length, response options, and predetermined subscales.

**Table 1:**
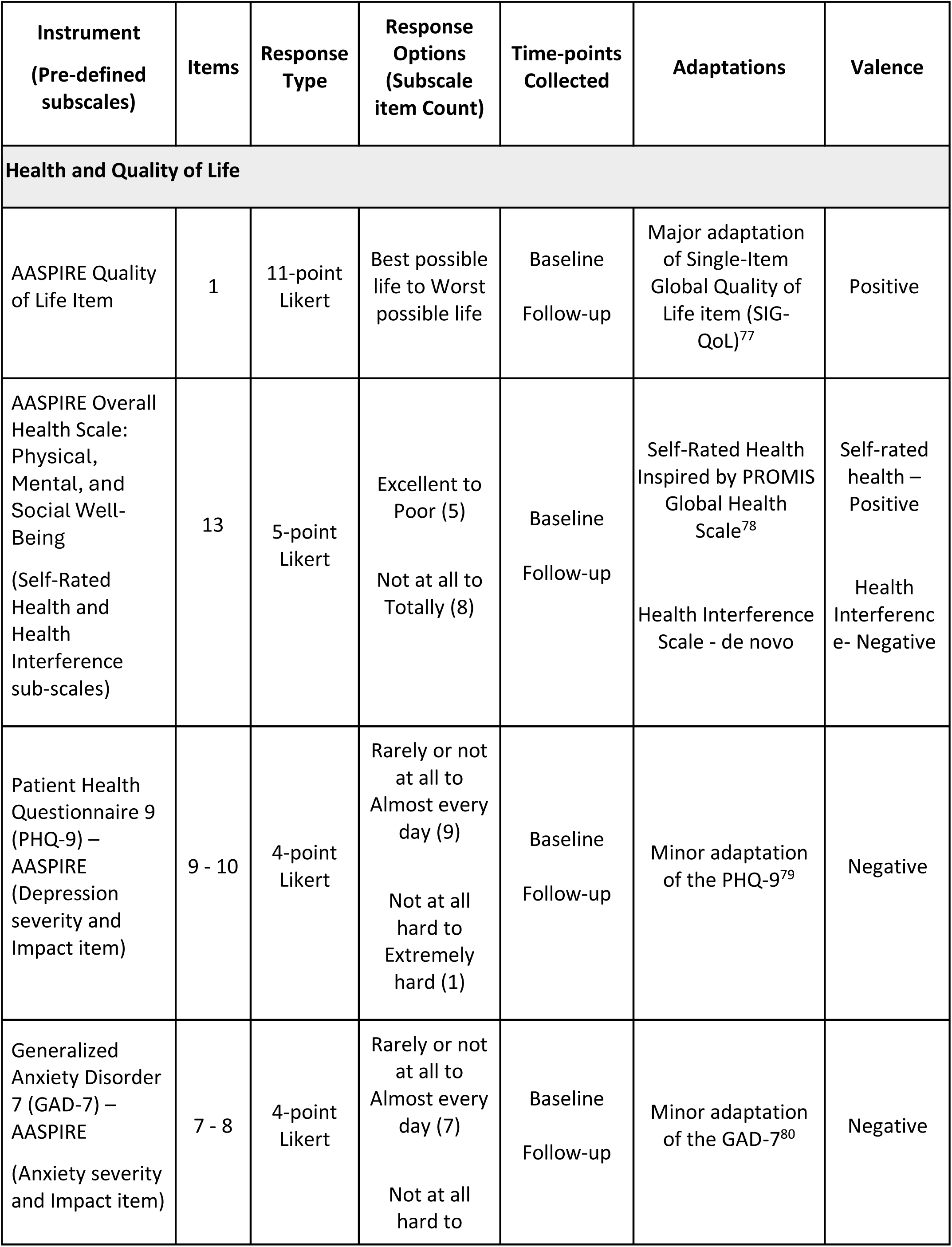
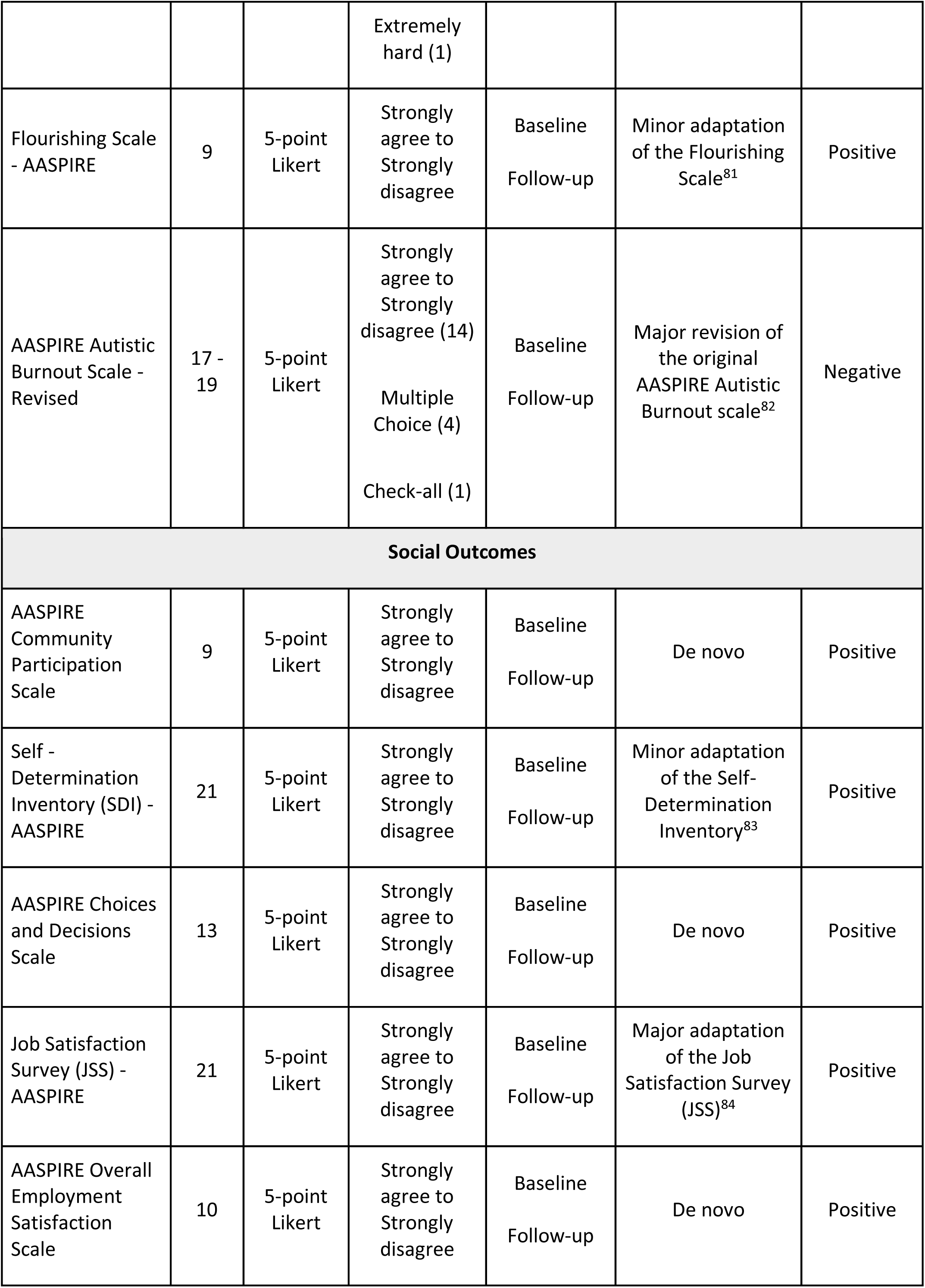
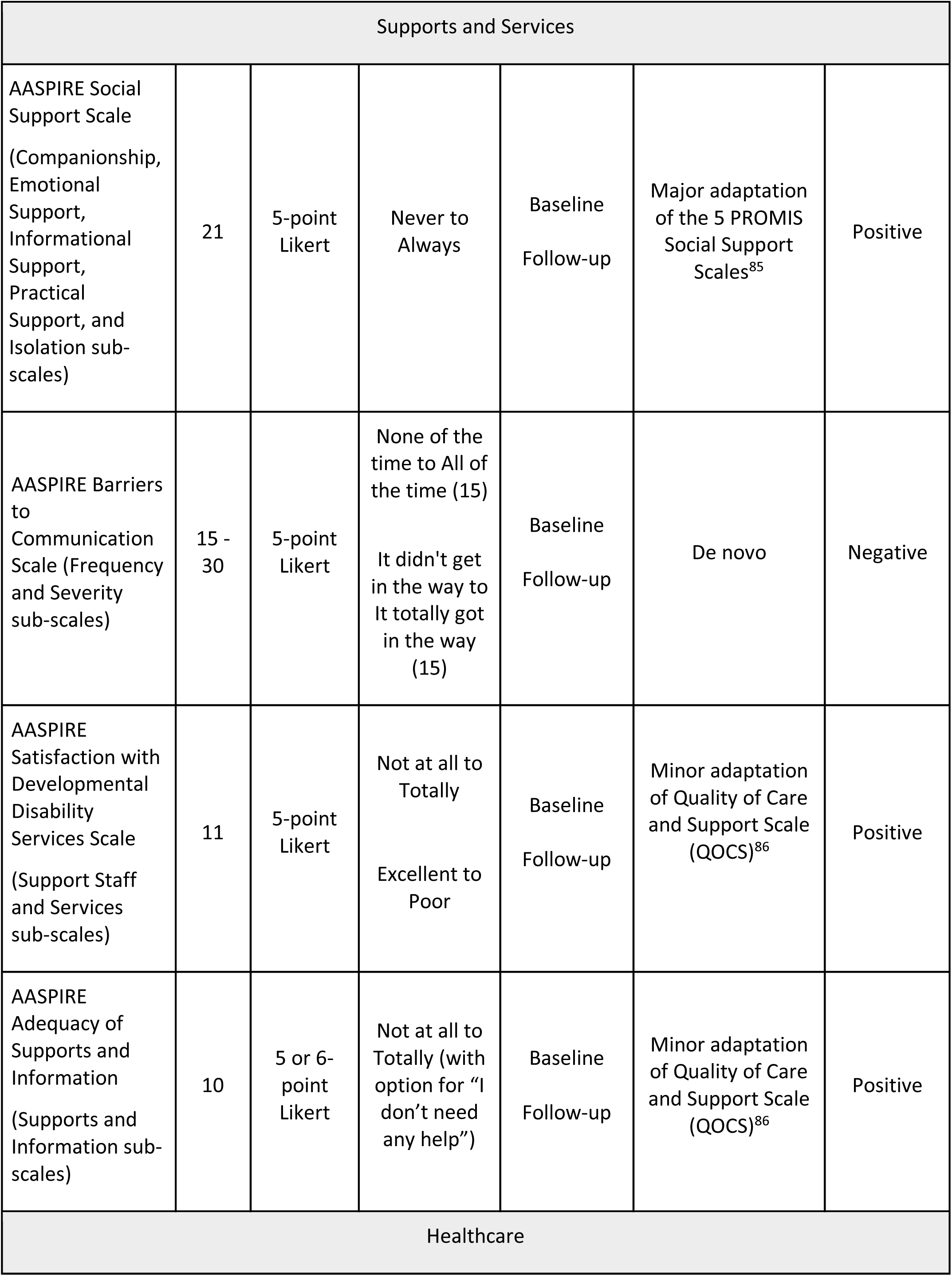
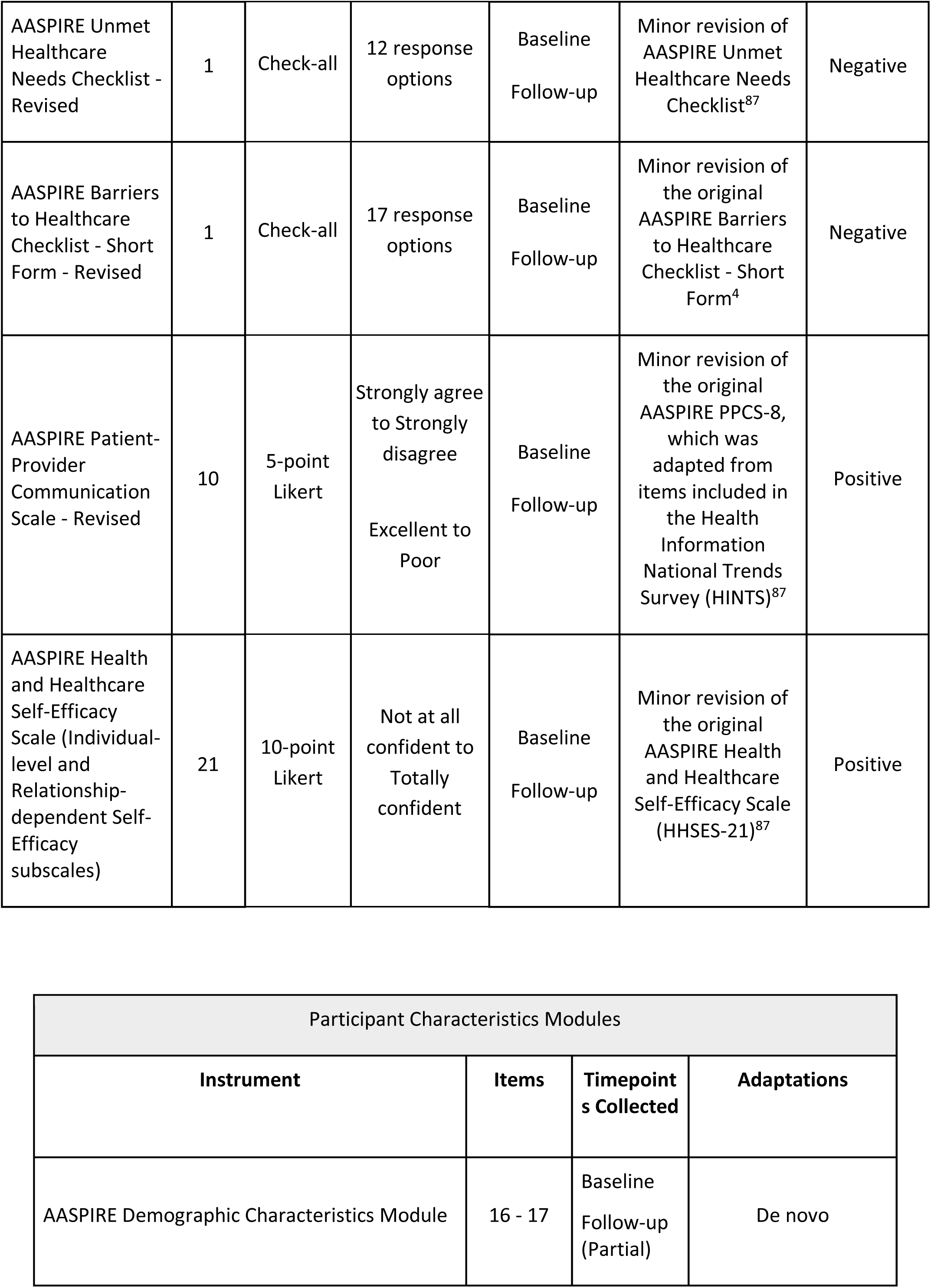
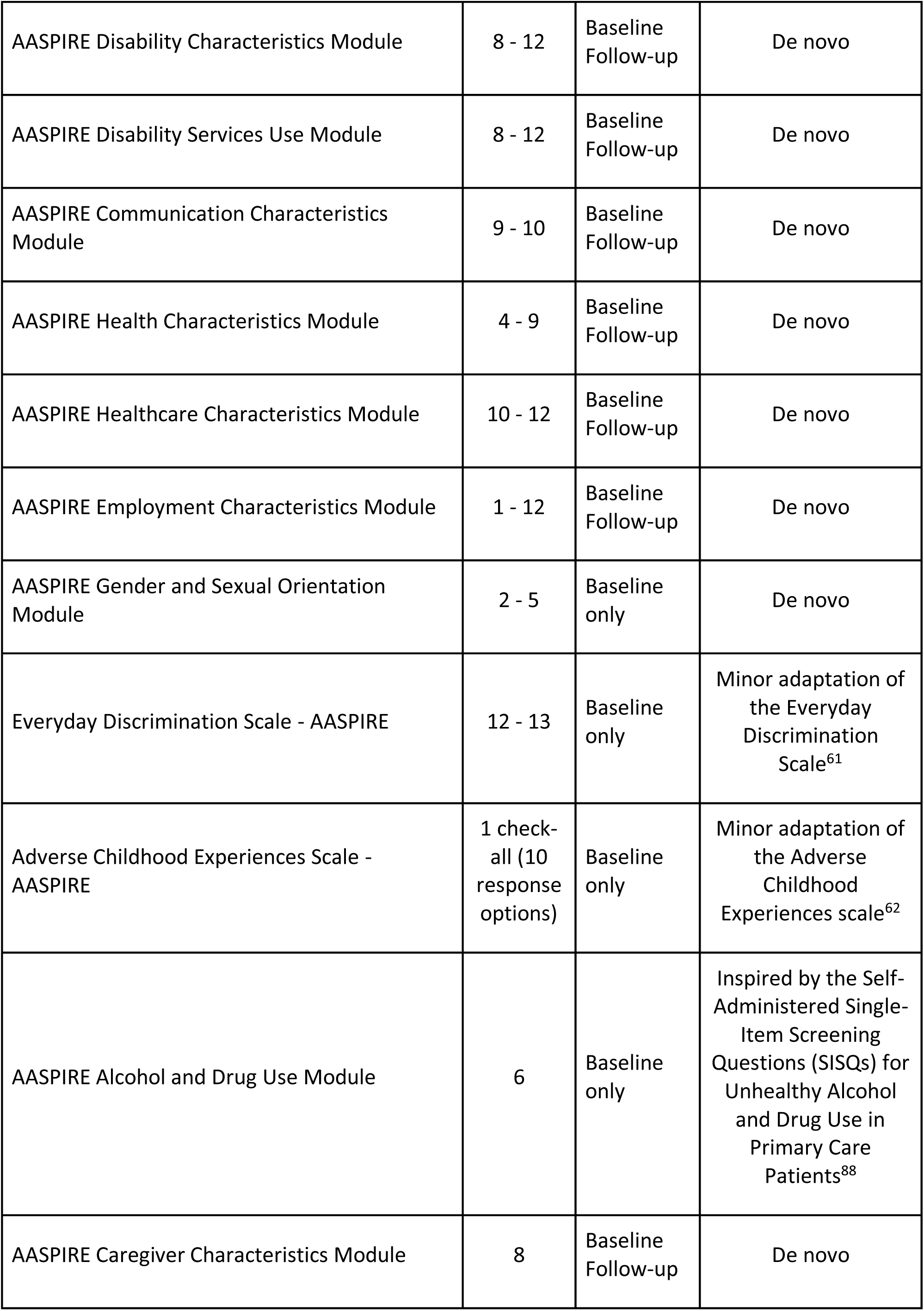
Outcome Measures and Participant Characteristics Modules.

| **Instrument**<br>**(Pre-defined subscales)** | **Items** | **Response Type** | **Response Options (Subscale item Count)** | **Time-points Collected** | **Adaptations** | **Valence** |
|---|---|---|---|---|---|---|
| **Health and Quality of Life** |  |  |  |  |  |  |
| AASPIRE Quality of Life Item | 1 | 11-point Likert | Best possible life to Worst possible life | Baseline<br>Follow-up | Major adaptation of Single-Item Global Quality of Life item (SIG-QoL)<sup>77</sup> | Positive |
| AASPIRE Overall Health Scale: Physical, Mental, and Social Well-Being<br>(Self-Rated Health and Health Interference sub-scales) | 13 | 5-point Likert | Excellent to Poor (5)<br>Not at all to Totally (8) | Baseline<br>Follow-up | Self-Rated Health Inspired by PROMIS Global Health Scale<sup>78</sup><br>Health Interference Scale - de novo | Self-rated health – Positive<br>Health Interference- Negative |
| Patient Health Questionnaire 9 (PHQ-9) – AASPIRE (Depression severity and Impact item) | 9 - 10 | 4-point Likert | Rarely or not at all to Almost every day (9)<br>Not at all hard to Extremely hard (1) | Baseline<br>Follow-up | Minor adaptation of the PHQ-9<sup>79</sup> | Negative |
| Generalized Anxiety Disorder 7 (GAD-7) – AASPIRE<br>(Anxiety severity and Impact item) | 7 - 8 | 4-point Likert | Rarely or not at all to Almost every day (7)<br>Not at all hard to Extremely hard (1) | Baseline<br>Follow-up | Minor adaptation of the GAD-7<sup>80</sup> | Negative |
| Flourishing Scale - AASPIRE | 9 | 5-point Likert | Strongly agree to Strongly disagree | Baseline<br>Follow-up | Minor adaptation of the Flourishing Scale<sup>81</sup> | Positive |
| AASPIRE Autistic Burnout Scale - Revised | 17 - 19 | 5-point Likert | Strongly agree to Strongly disagree (14)<br>Multiple Choice (4)<br>Check-all (1) | Baseline<br>Follow-up | Major revision of the original AASPIRE Autistic Burnout scale<sup>82</sup> | Negative |
| **Social Outcomes** |  |  |  |  |  |  |
| AASPIRE Community Participation Scale | 9 | 5-point Likert | Strongly agree to Strongly disagree | Baseline<br>Follow-up | De novo | Positive |
| Self - Determination Inventory (SDI) - AASPIRE | 21 | 5-point Likert | Strongly agree to Strongly disagree | Baseline<br>Follow-up | Minor adaptation of the Self-Determination Inventory<sup>83</sup> | Positive |
| AASPIRE Choices and Decisions Scale | 13 | 5-point Likert | Strongly agree to Strongly disagree | Baseline<br>Follow-up | De novo | Positive |
| Job Satisfaction Survey (JSS) - AASPIRE | 21 | 5-point Likert | Strongly agree to Strongly disagree | Baseline<br>Follow-up | Major adaptation of the Job Satisfaction Survey (JSS)<sup>84</sup> | Positive |
| AASPIRE Overall Employment Satisfaction Scale | 10 | 5-point Likert | Strongly agree to Strongly disagree | Baseline<br>Follow-up | De novo | Positive |
| Supports and Services |  |  |  |  |  |  |
| AASPIRE Social Support Scale<br>(Companionship, Emotional Support, Informational Support, Practical Support, and Isolation sub-scales) | 21 | 5-point Likert | Never to Always | Baseline<br>Follow-up | Major adaptation of the 5 PROMIS Social Support Scales<sup>85</sup> | Positive |
| AASPIRE Barriers to Communication Scale (Frequency and Severity sub-scales) | 15 - 30 | 5-point Likert | None of the time to All of the time (15)<br>It didn't get in the way to It totally got in the way (15) | Baseline<br>Follow-up | De novo | Negative |
| AASPIRE Satisfaction with Developmental Disability Services Scale<br>(Support Staff and Services sub-scales) | 11 | 5-point Likert | Not at all to Totally<br>Excellent to Poor | Baseline<br>Follow-up | Minor adaptation of Quality of Care and Support Scale (QOCS)<sup>86</sup> | Positive |
| AASPIRE Adequacy of Supports and Information<br>(Supports and Information sub-scales) | 10 | 5 or 6-point Likert | Not at all to Totally (with option for “I don’t need any help”) | Baseline<br>Follow-up | Minor adaptation of Quality of Care and Support Scale (QOCS)<sup>86</sup> | Positive |
| Healthcare |  |  |  |  |  |  |
| AASPIRE Unmet Healthcare Needs Checklist - Revised | 1 | Check-all | 12 response options | Baseline<br>Follow-up | Minor revision of AASPIRE Unmet Healthcare Needs Checklist<sup>87</sup> | Negative |
| AASPIRE Barriers to Healthcare Checklist - Short Form - Revised | 1 | Check-all | 17 response options | Baseline<br>Follow-up | Minor revision of the original AASPIRE Barriers to Healthcare Checklist - Short Form<sup>4</sup> | Negative |
| AASPIRE Patient-Provider Communication Scale - Revised | 10 | 5-point Likert | Strongly agree to Strongly disagree<br>Excellent to Poor | Baseline<br>Follow-up | Minor revision of the original AASPIRE PPCS-8, which was adapted from items included in the Health Information National Trends Survey (HINTS)<sup>87</sup> | Positive |
| AASPIRE Health and Healthcare Self-Efficacy Scale (Individual-level and Relationship-dependent Self-Efficacy subscales) | 21 | 10-point Likert | Not at all confident to Totally confident | Baseline<br>Follow-up | Minor revision of the original AASPIRE Health and Healthcare Self-Efficacy Scale (HHSES-21)<sup>87</sup> | Positive |
| Participant Characteristics Modules |  |  |  |  |  |  |
| **Instrument** | **Items** | **Timepoints Collected** | **Adaptations** |  |  |  |
| AASPIRE Demographic Characteristics Module | 16 - 17 | Baseline<br>Follow-up (Partial) | De novo |  |  |  |
| AASPIRE Disability Characteristics Module | 8 - 12 | Baseline<br>Follow-up | De novo |  |  |  |
| AASPIRE Disability Services Use Module | 8 - 12 | Baseline<br>Follow-up | De novo |  |  |  |
| AASPIRE Communication Characteristics Module | 9 - 10 | Baseline<br>Follow-up | De novo |  |  |  |
| AASPIRE Health Characteristics Module | 4 - 9 | Baseline<br>Follow-up | De novo |  |  |  |
| AASPIRE Healthcare Characteristics Module | 10 - 12 | Baseline<br>Follow-up | De novo |  |  |  |
| AASPIRE Employment Characteristics Module | 1 - 12 | Baseline<br>Follow-up | De novo |  |  |  |
| AASPIRE Gender and Sexual Orientation Module | 2 - 5 | Baseline only | De novo |  |  |  |
| Everyday Discrimination Scale - AASPIRE | 12 - 13 | Baseline only | Minor adaptation of the Everyday Discrimination Scale<sup>61</sup> |  |  |  |
| Adverse Childhood Experiences Scale - AASPIRE | 1 check-all (10 response options) | Baseline only | Minor adaptation of the Adverse Childhood Experiences scale<sup>62</sup> |  |  |  |
| AASPIRE Alcohol and Drug Use Module | 6 | Baseline only | Inspired by the Self-Administered Single-Item Screening Questions (SISQs) for Unhealthy Alcohol and Drug Use in Primary Care Patients<sup>88</sup> |  |  |  |
| AASPIRE Caregiver Characteristics Module | 8 | Baseline<br>Follow-up | De novo |  |  |  |

The survey for each time point included all outcome measures. However, we used branching logic to skip instruments not applicable to a subset of participants (e.g., we only asked participants receiving disability services their satisfaction with those services).

The DR version of each instrument asked if they received support while completing that specific instrument and if so, provided a checklist of the types of support they received. We used responses to classify participants into those who used the DR version independently (DR subgroup) or with support (DR-S subgroup). The CR version of each instrument included an item on how confident they felt about their answers and a checklist with options for how they came to know the information.

To help assess responsiveness to change, each instrument included a single item on perceived change. The item said, “Overall, in the last six months, did the things this survey asked about…” with response options “get worse,” “stay about the same,” and “get better.”

The baseline survey included modules to describe the sample, adjust for potential confounders, assess differences between subpopulations, and explore predictors of change in outcomes over time. Separate modules focused on demographic, disability, communication, health, and employment characteristics, as well as healthcare use, disability service use, and use of alcohol or illicit drugs. We also included minorly adapted versions of the Everyday Discrimination Scale^61^ and the Adverse Childhood Experiences Scale.^62^ A wrap-up section at the end of each half of the baseline survey asked how easy it was to understand the survey and how important the constructs were, and gave an opportunity for participants to make open-ended comments. We also included attention checks throughout the survey to help assess if participants were answering questions carefully. We repeated modules about characteristics likely to change (e.g. employment status) in the follow-up surveys.

The full baseline survey included a total of 319 to 383 items and follow-up surveys included 298 to 358 items, depending on branching patterns.

### Sample Characteristics

The full sample consists of 870 participants, including 319 in the HC subcohort; 266 in the DS subcohort; and 285 in the Community subcohort. There were 605 (70%) participants in the DR subgroup, 194 (22%) in the DR-S subgroup, and 71 (8%) in the CR subgroup. Overall, follow-up rates for the 2^nd^ and 3th time-points were 87% and 81% respectively.

Participants were 18 to 77 years old (mean 31 years; SD 11.6). The sample includes heterogeneity in gender, sex assigned at birth (approximately 1:1 ratio of male to female), and sexual orientation, with almost half of the sample identifying as LGBTQ+ (lesbian, gay, bisexual, trans, queer, plus other). Approximately 30% of participants were Black, Indigenous, or People of Color (BIPOC). A little over half the sample found it hard or very hard to afford basic necessities. Half the sample had at least some paid employment.

There was a wide spread of educational attainment, with 32% having a high school degree or less; 35% having some college or an associate’s degree; and 33% with bachelor’s or graduate degrees. Approximately a quarter of the sample could not read or had trouble reading.

Over 25% required support with basic activities of daily living (ADLs) and another 44% required support with instrumental activities of daily living (IADLs). Approximately equal numbers lived in their own home or student housing (46%) vs their family’s home (45%), with a small proportion (9%) living in other settings (e.g., group homes, facilities, shelters). Almost half (45%) received disability services.

About a quarter reported they had no difficulty speaking, 45% reported they could speak fairly well, and 30% found speech difficult or impossible. Conversely, 39% had no difficulty understanding speech, 49% could understand speech fairly well, and 12% had a difficult time understanding speech under daily conditions.

A majority (61%) of the sample had at least one physical condition, 55% had attention deficit hyperactivity disorder (ADHD), and 75% had at least one other co-occurring mental health condition.

**Table 2** presents information about study participation and participant characteristics. Supplements B1-4 present sample characteristics of various subgroups (see below) and caregiver reporters. A separate paper provides a more detailed description of the study sample.^51^

**Table 2:** Participant Characteristics.

| Characteristic |  | N | % |
|---|---|---|---|
| **Study Participation** |  |  |  |
| Subcohort | Disability Services (DDS) | 266 | 31% |
|  | Healthcare | 319 | 37% |
|  | Community | 285 | 33% |
| Reporter type | Direct Report (DR) | 605 | 70% |
|  | DR – Supported (DR-S) | 194 | 22% |
|  | Caregiver Report (CR) | 71 | 8% |
| **Demographic Characteristics** |  |  |  |
| Age group | 18–29 | 505 | 58% |
|  | 30–39 | 200 | 23% |
|  | 40–49 | 93 | 11% |
|  | 50+ | 71 | 8% |
| BIPOC<sup>1</sup> | Yes | 244 | 30% |
| Race/ethnicity | Asian | 35 | 4% |
|  | Biracial / multi-racial | 30 | 4% |
|  | Black, African American, or of African ancestry | 31 | 4% |
|  | Native American or Native Alaskan | 15 | 2% |
|  | Native Hawaiian or other Pacific Islander | 1 | 0% |
|  | White | 607 | 73% |
|  | Hispanic | 97 | 12% |
|  | Other | 12 | 1% |
| Gender | Female | 346 | 40% |
|  | Male | 393 | 45% |
|  | Non-binary / agender / genderfluid | 267 | 31% |
| Sex assigned at birth | Male | 404 | 49% |
|  | Female | 427 | 51% |
|  | Other | 2 | 0% |
| Gender same as SAB<sup>2</sup> | Yes | 616 | 79% |
| Sexual and gender minority status | LGBTQIA+<sup>3</sup> | 403 | 48% |
| Educational attainment | Less than high school | 59 | 7% |
|  | Modified HS Diploma | 92 | 11% |
|  | Regular HS Diploma | 116 | 14% |
|  | Some college | 201 | 25% |
|  | Associates degree / certificate | 84 | 10% |
|  | Bachelors degree | 148 | 18% |
|  | Graduate degree | 120 | 15% |
| Received Special Education services<sup>4</sup> | Yes | 579 | 70% |
| Living Situation | Own home (including student housing) | 384 | 46% |
|  | Family's home | 379 | 45% |
|  | Group home, facility, unhoused, or other<sup>5</sup> | 72 | 9% |
| Financial Hardship (difficulty affording basic necessities) | Very hard | 170 | 20% |
|  | Somewhat hard | 275 | 33% |
|  | Not hard at all | 387 | 47% |
| Employed for pay | Yes | 420 | 50% |
| **Disability Characteristics** |  |  |  |
| Autism diagnosis<sup>6</sup> | Formal (medical or educational) | 787 | 92% |
|  | Self-diagnosed | 69 | 8% |
| Co-occurring ADHD | Yes (formal or self-diagnosed) | 460 | 55% |
| Support needs<sup>7</sup> | Needs assistance with ADLs | 221 | 26% |
|  | Needs assistance with IADLs only | 371 | 44% |
|  | Does not need assistance | 243 | 29% |
| Receive disability services | Yes | 360 | 45% |
| **Communication Characteristics** |  |  |  |
| Understand speech under daily conditions<sup>8</sup> | No difficulty | 325 | 39% |
|  | Fairly well | 414 | 49% |
|  | Difficult time / or minimal | 101 | 12% |
| Speak under daily conditions<sup>8</sup> | No difficulty | 214 | 25% |
|  | Fairly well | 376 | 45% |
|  | Difficult time / or minimal | 250 | 30% |
| Reading level | Advanced | 627 | 75% |
|  | Basic (e.g., uses "Plain Language" materials) | 143 | 17% |
|  | Minimal or none | 71 | 8% |
| Writing level | Advanced | 563 | 67% |
|  | Basic (e.g. can explain simple ideas) | 192 | 23% |
|  | Minimal or none | 87 | 10% |
| **Health Status and Co-occurring Conditions** |  |  |  |
| Health Status | Poor | 69 | 8% |
|  | Fair | 205 | 25% |
|  | Good | 297 | 36% |
|  | Very Good | 176 | 21% |
|  | Excellent | 85 | 10% |
| Co-occurring ADHD | Yes (formal or self-diagnosed) | 460 | 55% |
| Other Mental Health Conditions<sup>9</sup> | Depression | 374 | 43% |
|  | Anxiety | 525 | 60% |
|  | PTSD | 229 | 26% |
|  | Bipolar | 60 | 7% |
|  | Schizophrenia or psychosis | 21 | 3% |
| Any mental health (other than autism or ADHD) | Yes | 616 | 75% |
| Number of physical conditions | 0 | 321 | 38% |
|  | 1 | 179 | 21% |
|  | 2 or more | 335 | 40% |

<sup>1</sup> Black, Indigenous, and People of Color
<sup>2</sup> Sex Assigned at Birth
<sup>3</sup> Lesbian, Gay, Bisexual, Transgender, Queer, Intersex, Asexual, plus others. Includes participants who chose any of the following categories on a check-all item about sexual orientation: “Gay or lesbian”, “Bi or bisexual”, “Queer”, “Pansexual”, “Asexual”, “Demisexual”, or on a check-all item about gender: “Nonbinary,” “Trans woman,” “Trans man,” “Trans or transgender,” “Genderqueer,” “Genderfluid,” or “Agender or no gender.”
<sup>4</sup> Including an IEP, a 504 plan, or other special education services.
<sup>5</sup> Including foster homes, adult care homes, residential treatment centers, long-term care, emergency or transitional housing, institutional housing, and buildings not meant for housing.
<sup>6</sup> Formal medical or educational diagnosis vs. self-diagnosis of autism, ASD, Asperger's disorder, or PDD-NOS.
<sup>7</sup> Needs assistance with ADLs = requires support for basic self-care activities; Needs assistance with IADLs only = requires support for instrumental activities but not basic self-care; Does not need assistance = rarely or never requires either.
<sup>8</sup> Daily conditions, including when the environment may be disruptive or overstimulating, and when a person may feel tired, hungry, or unwell.
<sup>9</sup> Other than autism or ADHD. Column totals may exceed 100% as multiple conditions were permitted.
Note: Percentages for gender, race/ethnicity, and mental health condition items may sum to >100% as multiple responses were permitted. All other percentages are column percentages within each group. Total N varies across rows due to missing data.

### Data Analysis

We assessed the psychometric properties of our outcome measures in a variety of ways.

First, we ensured **content validity,** as per best practice guidelines,^56,63^ via a combination of strategies, including reviewing candidate measures with Delphi panel participants, conducting iterative stakeholder reviews with our academic and community team members throughout the adaptation and development of the instruments, and conducting cognitive interviews with autistic adults and supporters.

Second, we assessed **accessibility** by summarizing responses to a question about how easy the items on each half of the baseline survey were to understand, by calculating completion rates at each time point, and by reviewing open-ended comments related to accessibility.

Third, we assessed **internal consistency reliability** by calculating Cronbach’s alpha for each measure and any pre-determined subscales.

Fourth, we assessed **test-retest reliability** using a retest survey administered approximately two weeks after the 2^nd^ time point to a random sample of 15% of participants. We calculated single-measure intraclass correlation coefficients (ICCs) from one-way random effects models.^64,65^ As a sensitivity analysis, we also estimated test-retest ICCs using two-way mixed-effects models with absolute agreement, treating time point (time 2 survey vs. retest survey) as a fixed effect.

Fifth, we assessed **convergent and discriminant validity** using pair-wise Pearson’s correlations between each of the outcome measures or predetermined subscales. Consistent with the COSMIN approach to hypothesis testing, we prespecified our expectations for each comparison.^66–68^

For two of the outcomes, the toolkit includes pairs of scales that measure the same broad construct in different ways. Specifically, we included a single-item measure of Quality of Life, and the Self-Rated Health Scale of our Overall Health: Physical, Mental, and Social Wellbeing, which can be conceptualized as another measure of quality of life. Similarly, the toolkit includes two measures of employment satisfaction. Since each pair targets the same construct, we hypothesized very high correlations (above 0.70).^66^ Because we designed the other instruments to capture distinct constructs, we hypothesized that correlations between them would fall below 0.85, a level at which the instruments would be largely redundant. We considered correlations with absolute values below 0.20 to be negligible.^66^

We identified sets of outcomes likely to have moderate to high correlations among them (absolute values of r between 0.3-0.7)^66^. These sets included mental health (anxiety, depression, and autistic burnout); support (social support, adequacy of supports and information, and satisfaction with disability services), and quality of healthcare (unmet healthcare needs, barriers to healthcare, patient-provider communication, individual-level healthcare self-efficacy, and relationship-dependent self-efficacy). Similarly, we expected a moderate correlation between self-determination and the ability or opportunity to make choices and decisions. We hypothesized that all mental health outcomes, community participation, self-determination, and social support would have moderate to high correlations with overall health, quality of life, and flourishing.

We looked for general patterns across the entire correlation matrix, expecting that more closely related constructs would have higher correlations than more distant constructs. We will present more detailed analyses of construct validity, with instrument-specific hypotheses separately.

Sixth, we assessed external **responsiveness to change** for each measure by comparing self-reported perceptions of change with changes in scale scores. In most cases, instruments included one perceived change item for the entire construct, which we used both for the total score and predefined subscales. Two instruments (social support and satisfaction with disability services) included a perceived change item for each subscale, so we assessed responsiveness to change for each subscale separately. We calculated Time 2 change scores by subtracting mean scores at Time 1 from mean scores at Time 2, and we calculated Time 3 change scores by subtracting mean Time 2 scores from mean scores at Time 3. We used linear regression with robust standard errors clustered by participants to compare change scores between participants who reported that for them the outcome had improved, stayed the same, or worsened. We used the ‘stayed the same’ group as the reference category.

We conducted **subgroup analyses** to assess participant characteristics, internal consistency reliability, construct validity, and responsiveness to change in three sets of subgroups defined by subcohort (HC, DS, or Community), support needs for daily living (support for ADLs, IADLs only, or neither), and reporter type (DR, DR-S, CR).

Most instruments are scored by summing Likert scale responses to individual items. To account for missing data, we used mean scores (dividing the sum by the number of items answered) in all analyses that involved scale scores. We scored the AASPIRE Barriers to Communication Scale differently: for each barrier, we multiplied the frequency rating (0 to 4) by the severity rating (1 to 5), assigned a product of 0 to barriers that participants reported never experiencing, and averaged these products across barriers, producing scores that range from 0 to 20. We summed checklist responses to produce count scores. We examined distributional properties (skewness, kurtosis, SD) for all outcome variables. Given the positively skewed count distributions of the Unmet Healthcare Needs and Barriers to Healthcare checklists and the single-item design of the Quality of Life measure, we calculated Spearman’s rho as a non-parametric test-retest reliability coefficient for these three instruments as **sensitivity analyses**. We also replicated the responsiveness analyses for the checklist variables using Wilcoxon signed-rank tests with rank-biserial correlations.

As additional analyses related to **construct validity and structural validity** require instrument-specific methods and detailed results, we will present further in-depth psychometric analyses in separate papers focused on each measure or small sets of measures.

## Results

### Content validity and accessibility

We used data from the CBPR-nested Delphi process^50^ and discussions with the academic and community members of our team to assess how well existing instruments captured each intended construct and how much they would need to be adapted to be accessible to autistic adults with a wide range of characteristics. Our team identified instruments that could adequately capture the constructs of depression, anxiety, flourishing, satisfaction with disability services, and adequacy of supports and services with only minor adaptations. We also felt that we could adequately measure healthcare quality by making minor revisions to four instruments that AASPIRE developed in prior projects. We felt we could measure quality of life, social support, and autistic burnout with major adaptations or revisions of existing measures. In the case of self-determination, employment satisfaction, and overall health, we identified measures that could adequately capture part of the intended construct (with minor or major adaptations), but we felt we would need to create an additional instrument or subscale to measure other important aspects of the construct. While there were many measures assessing communication effectiveness, we could not find any measure to capture the construct of *access* to communication. Similarly, while we identified multiple instruments to assess community participation, we were concerned that it would be hard to use them as outcome measures in intervention studies because their structure may make them less likely to be responsive to change.

Common accessibility concerns with existing instruments included 1) difficult vocabulary, 2) complex sentence structure, 3) imprecise language, 4) double-barreled items, 5) confusing or overlapping response options, 6) potential for responses to vary at different times or in different situations, and 7) lack of precise timeframes or difficult to visualize timeframes. The team addressed concerns through our CBPR process by 1) simplifying vocabulary or sentence structure, 2) including hotlinks to define terms and/or offer examples, 3) finding a more general way to ask about different parts of a double-barreled item and then including those parts as “examples,” 4) adding prefaces to provide additional context, 5) adding links for participants who need additional explanations, and 6) providing links to vignettes with how different participants may answer the items.

We used discussions with our full team and cognitive interviews with 37 participants to reassess the content validity and accessibility of the adapted, revised, or newly created measures. In most cases, cognitive interviews identified the need for only minor revisions. However, we identified significant problems with our original measures of quality of life and freedom to make choices and decisions, requiring us to abandon the draft instruments and go back to the full team to adapt or create new instruments. Cognitive interview participants had more positive reactions to the new versions (in preparation).

Supplement C1 details the types of support participants used to take the DR version of the survey in the longitudinal study. Over 90% of participants who started the survey at each time point completed all applicable sections. Furthermore, participants who started each instrument completed, on average, 99% of items, with fewer than 5% of participants missing more than 10% of items (**Supplement C2**). Follow-up rates at Time 2 and 3 were >80% for the full sample, and >70% in all subgroups divided by cohort or support needs, but they were lower for participants who required support to take the survey directly (Supplement C3).

Over 90% of all participants felt that it was very or extremely easy to understand the questions on the survey. A lower proportion (84%–85%) of participants who needed help to take the survey directly felt it was easy to understand the items than caregivers (94-97%) or those who didn’t need help (96%; p<0.0001) but at least 90% of participants found it easy to understand in all subgroups divided by need for support with daily activities (Supplement C4).

While most people who used the general comment boxes at the end of each survey explained or elaborated on answers, some commented about accessibility. Sporadic comments noted a specific wording choice a participant personally struggled with or a desire for additional responses on particular multiple-choice options, but none were prevalent or significant enough to warrant changing instruments. Other comments praised the survey’s accessibility, using terms such as “thoughtfully-designed,” “easy-to-understand,” “non-ambiguous,” “well-constructed,” and “fabulous” to describe the overall survey.

Many comments praised specific strategies we used to increase accessibility. For example, participants wrote:

- “This survey is so well-done for my autistic mind. The explanations of each section, definitions of terms, and examples to clarify in ambiguous cases were extremely helpful.”
- “Thank you for wording the questions the way you did. It made it very easy to understand & added the right amount of context I think. Also having examples of the abstract thoughts/feelings/ideas asked about was extremely helpful.”
- “…The visual icons like the filled cups were great support too. I have some dyscalculia, and usually I have to go back and reread numbered rating scales over and over, which can be frustrating, but the visual icons for less and more made it way easier to see.”
- “Bravo for including hover-text definitions and elaborations on things! I only sometimes struggle to understand things like that, but when I do (and I did a few times in this survey), accommodations like that make a huge difference.”

### Internal consistency reliability

**Table 3** presents Cronbach’s alphas for all scored outcome measures and any pre-defined subscales. All scales demonstrated good to excellent internal consistency (α > 0.80), with most showing excellent reliability (α ≥ 0.90) in the full sample. Results were similar across subsamples, with alphas >0.7 in all subgroup analyses (Supplements D1-3).

**Table 3:**
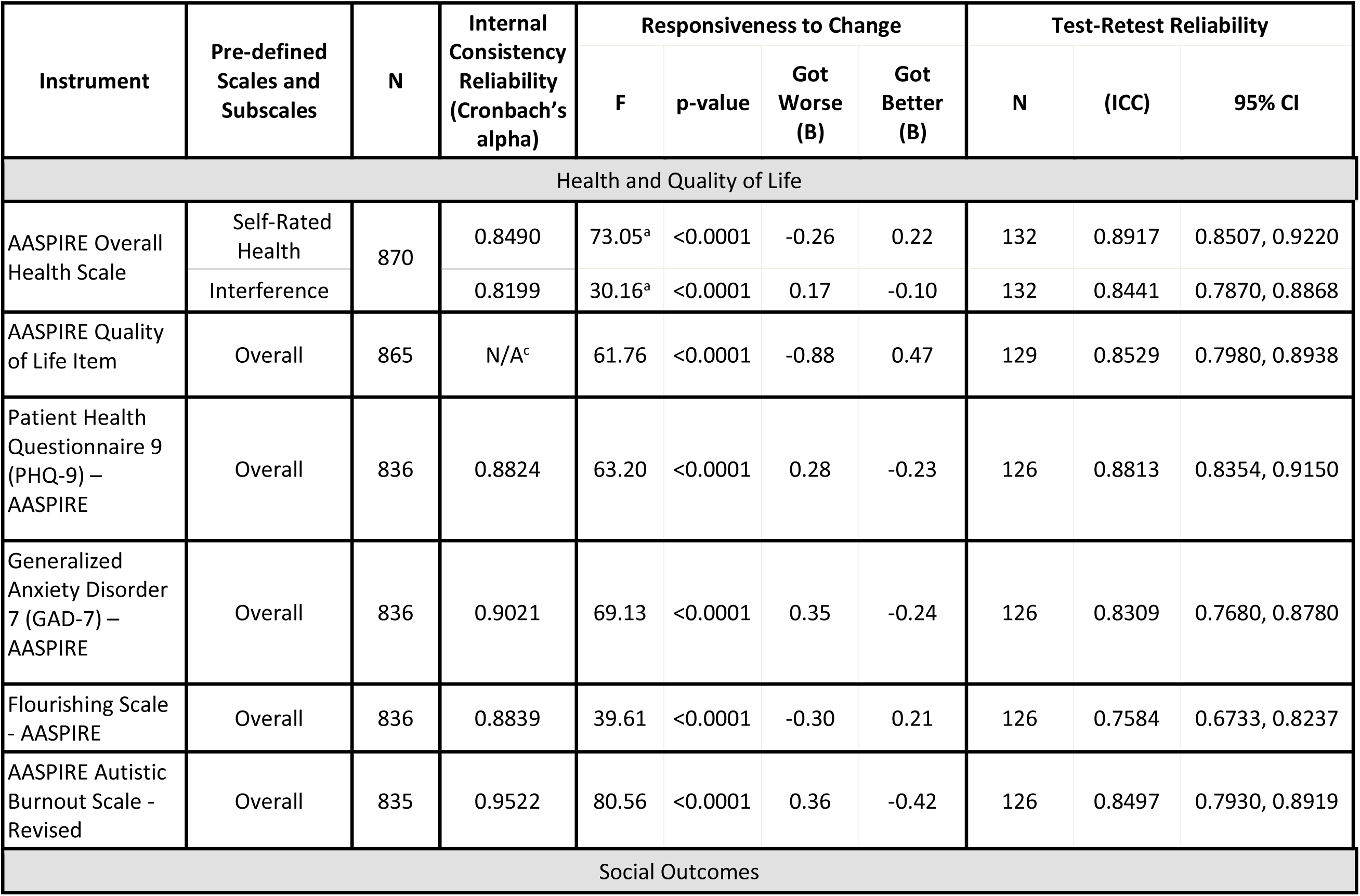
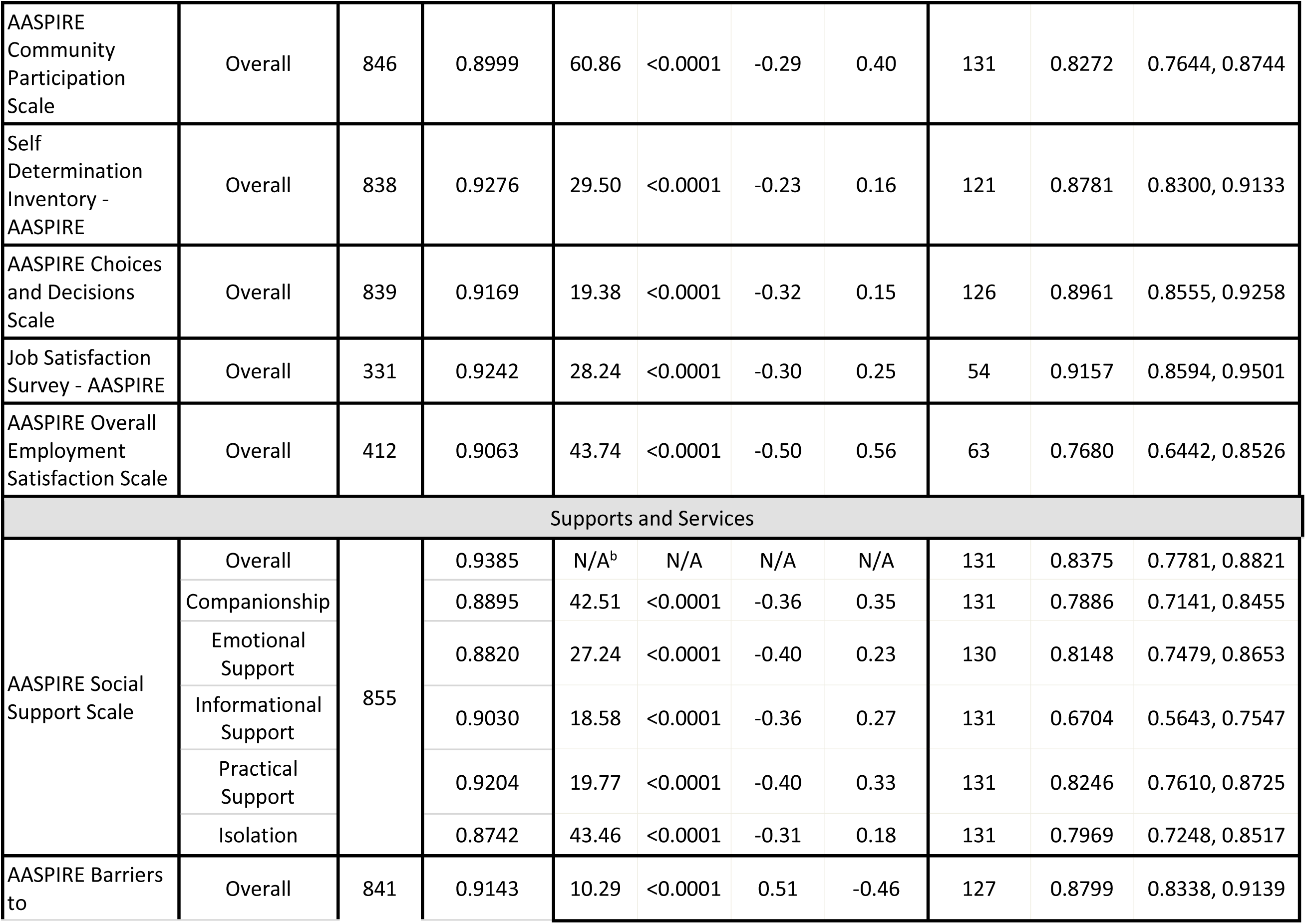
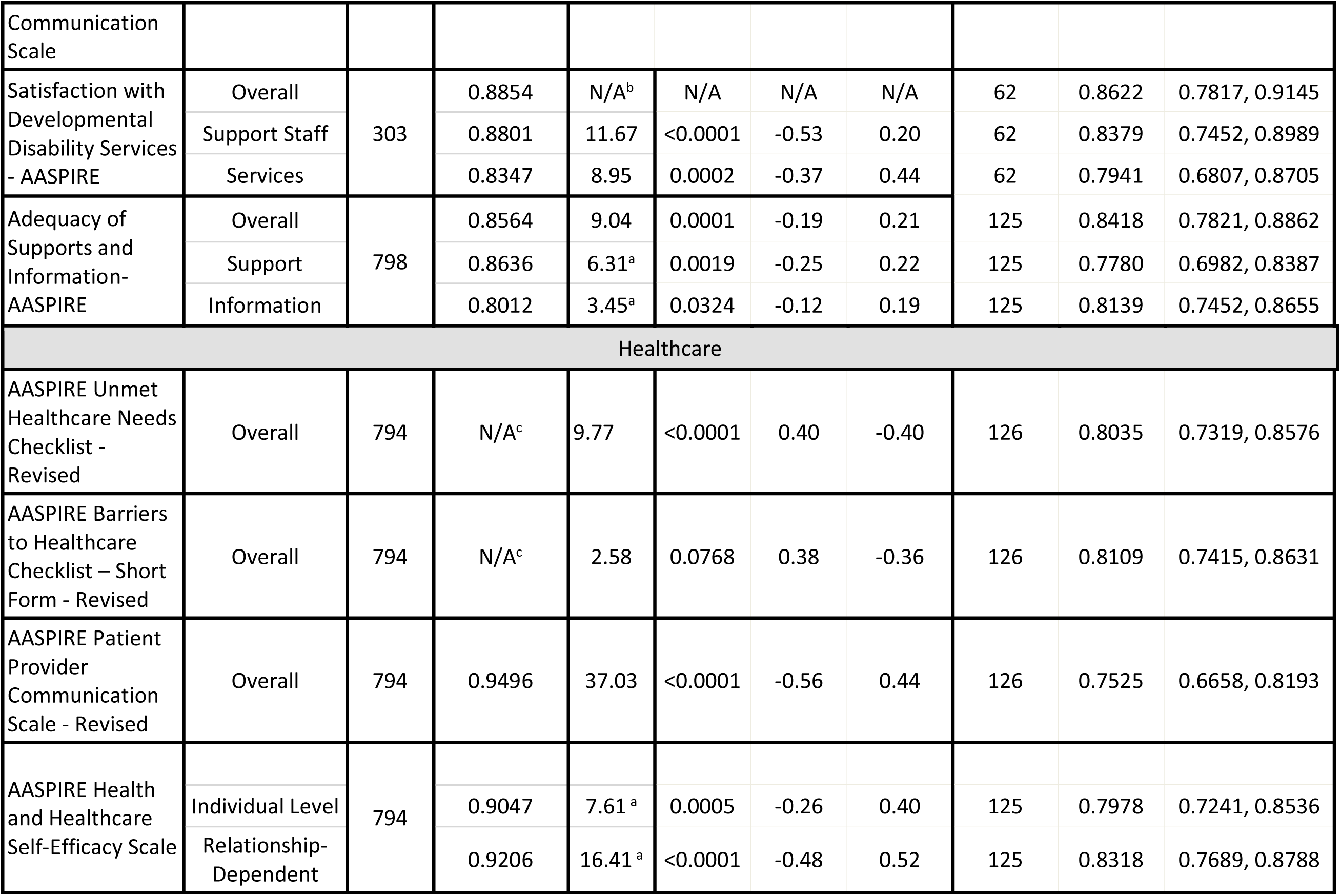
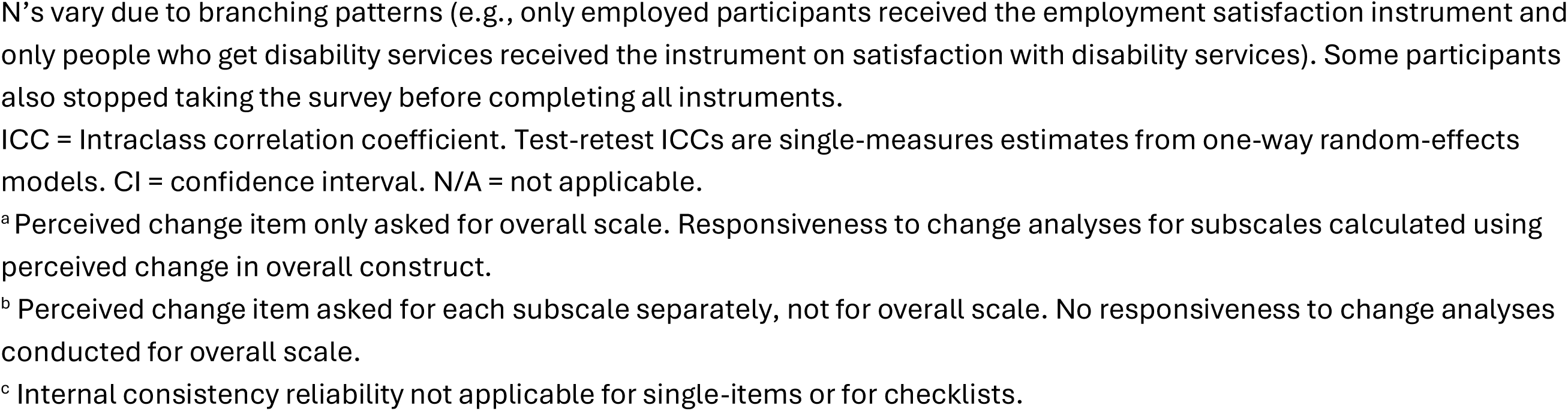
Reliability and Responsiveness to Change.

| Instrument | Pre-defined Scales and Subscales | N | Internal Consistency Reliability (Cronbach’s alpha) | Responsiveness to Change |  |  |  | Test-Retest Reliability |  |  |
|---|---|---|---|---|---|---|---|---|---|---|
|  |  |  |  | F | p-value | Got Worse (B) | Got Better (B) | N | (ICC) | 95% CI |
| Health and Quality of Life |  |  |  |  |  |  |  |  |  |  |
| AASPIRE Overall Health Scale | Self-Rated Health | 870 | 0.8490 | 73.05<sup>a</sup> | <0.0001 | -0.26 | 0.22 | 132 | 0.8917 | 0.8507, 0.9220 |
|  | Interference |  | 0.8199 | 30.16<sup>a</sup> | <0.0001 | 0.17 | -0.10 | 132 | 0.8441 | 0.7870, 0.8868 |
| AASPIRE Quality of Life Item | Overall | 865 | N/A<sup>c</sup> | 61.76 | <0.0001 | -0.88 | 0.47 | 129 | 0.8529 | 0.7980, 0.8938 |
| Patient Health Questionnaire 9 (PHQ-9) – AASPIRE | Overall | 836 | 0.8824 | 63.20 | <0.0001 | 0.28 | -0.23 | 126 | 0.8813 | 0.8354, 0.9150 |
| Generalized Anxiety Disorder 7 (GAD-7) – AASPIRE | Overall | 836 | 0.9021 | 69.13 | <0.0001 | 0.35 | -0.24 | 126 | 0.8309 | 0.7680, 0.8780 |
| Flourishing Scale - AASPIRE | Overall | 836 | 0.8839 | 39.61 | <0.0001 | -0.30 | 0.21 | 126 | 0.7584 | 0.6733, 0.8237 |
| AASPIRE Autistic Burnout Scale - Revised | Overall | 835 | 0.9522 | 80.56 | <0.0001 | 0.36 | -0.42 | 126 | 0.8497 | 0.7930, 0.8919 |
| Social Outcomes |  |  |  |  |  |  |  |  |  |  |
| AASPIRE Community Participation Scale | Overall | 846 | 0.8999 | 60.86 | <0.0001 | -0.29 | 0.40 | 131 | 0.8272 | 0.7644, 0.8744 |
| Self Determination Inventory - AASPIRE | Overall | 838 | 0.9276 | 29.50 | <0.0001 | -0.23 | 0.16 | 121 | 0.8781 | 0.8300, 0.9133 |
| AASPIRE Choices and Decisions Scale | Overall | 839 | 0.9169 | 19.38 | <0.0001 | -0.32 | 0.15 | 126 | 0.8961 | 0.8555, 0.9258 |
| Job Satisfaction Survey - AASPIRE | Overall | 331 | 0.9242 | 28.24 | <0.0001 | -0.30 | 0.25 | 54 | 0.9157 | 0.8594, 0.9501 |
| AASPIRE Overall Employment Satisfaction Scale | Overall | 412 | 0.9063 | 43.74 | <0.0001 | -0.50 | 0.56 | 63 | 0.7680 | 0.6442, 0.8526 |
| Supports and Services |  |  |  |  |  |  |  |  |  |  |
| AASPIRE Social Support Scale | Overall | 855 | 0.9385 | N/A<sup>b</sup> | N/A | N/A | N/A | 131 | 0.8375 | 0.7781, 0.8821 |
|  | Companionship |  | 0.8895 | 42.51 | <0.0001 | -0.36 | 0.35 | 131 | 0.7886 | 0.7141, 0.8455 |
|  | Emotional Support |  | 0.8820 | 27.24 | <0.0001 | -0.40 | 0.23 | 130 | 0.8148 | 0.7479, 0.8653 |
|  | Informational Support |  | 0.9030 | 18.58 | <0.0001 | -0.36 | 0.27 | 131 | 0.6704 | 0.5643, 0.7547 |
|  | Practical Support |  | 0.9204 | 19.77 | <0.0001 | -0.40 | 0.33 | 131 | 0.8246 | 0.7610, 0.8725 |
|  | Isolation |  | 0.8742 | 43.46 | <0.0001 | -0.31 | 0.18 | 131 | 0.7969 | 0.7248, 0.8517 |
| AASPIRE Barriers to Communication Scale | Overall | 841 | 0.9143 | 10.29 | <0.0001 | 0.51 | -0.46 | 127 | 0.8799 | 0.8338, 0.9139 |
| Satisfaction with Developmental Disability Services - AASPIRE | Overall | 303 | 0.8854 | N/A<sup>b</sup> | N/A | N/A | N/A | 62 | 0.8622 | 0.7817, 0.9145 |
|  | Support Staff |  | 0.8801 | 11.67 | <0.0001 | -0.53 | 0.20 | 62 | 0.8379 | 0.7452, 0.8989 |
|  | Services |  | 0.8347 | 8.95 | 0.0002 | -0.37 | 0.44 | 62 | 0.7941 | 0.6807, 0.8705 |
| Adequacy of Supports and Information-AASPIRE | Overall | 798 | 0.8564 | 9.04 | 0.0001 | -0.19 | 0.21 | 125 | 0.8418 | 0.7821, 0.8862 |
|  | Support |  | 0.8636 | 6.31<sup>a</sup> | 0.0019 | -0.25 | 0.22 | 125 | 0.7780 | 0.6982, 0.8387 |
|  | Information |  | 0.8012 | 3.45<sup>a</sup> | 0.0324 | -0.12 | 0.19 | 125 | 0.8139 | 0.7452, 0.8655 |
| Healthcare |  |  |  |  |  |  |  |  |  |  |
| AASPIRE Unmet Healthcare Needs Checklist - Revised | Overall | 794 | N/A<sup>c</sup> | 9.77 | <0.0001 | 0.40 | -0.40 | 126 | 0.8035 | 0.7319, 0.8576 |
| AASPIRE Barriers to Healthcare Checklist – Short Form - Revised | Overall | 794 | N/A<sup>c</sup> | 2.58 | 0.0768 | 0.38 | -0.36 | 126 | 0.8109 | 0.7415, 0.8631 |
| AASPIRE Patient Provider Communication Scale - Revised | Overall | 794 | 0.9496 | 37.03 | <0.0001 | -0.56 | 0.44 | 126 | 0.7525 | 0.6658, 0.8193 |
| AASPIRE Health and Healthcare Self-Efficacy Scale |  | 794 |  |  |  |  |  |  |  |  |
|  | Individual Level |  | 0.9047 | 7.61<sup>a</sup> | 0.0005 | -0.26 | 0.40 | 125 | 0.7978 | 0.7241, 0.8536 |
|  | Relationship-Dependent |  | 0.9206 | 16.41<sup>a</sup> | <0.0001 | -0.48 | 0.52 | 125 | 0.8318 | 0.7689, 0.8788 |

N’s vary due to branching patterns (e.g., only employed participants received the employment satisfaction instrument and only people who get disability services received the instrument on satisfaction with disability services). Some participants also stopped taking the survey before completing all instruments.
ICC = Intraclass correlation coefficient. Test-retest ICCs are single-measures estimates from one-way random-effects models. CI = confidence interval. N/A = not applicable.
<sup>a</sup>Perceived change item only asked for overall scale. Responsiveness to change analyses for subscales calculated using perceived change in overall construct.
<sup>b</sup> Perceived change item asked for each subscale separately, not for overall scale. No responsiveness to change analyses conducted for overall scale.
<sup>c</sup> Internal consistency reliability not applicable for single-items or for checklists.

### Convergent and discriminant validity

All correlations were in the expected directions (e.g., negatively valenced scales had negative correlations with positively valenced scales and positive correlations with other negatively valenced scales).

All comparisons between scales measuring separate constructs had correlation coefficients with absolute values below 0.85, demonstrating adequate discriminant validity.

Both pairs of measures assessing similar constructs had high correlations, with a correlation of 0.75 between the single item QoL measure and the self-rated health scale in the AASPIRE Overall Health: Physical, Mental, Social Wellbeing instrument, and a correlation of 0.86 between the two employment scales.

The three mental health scales — depression, anxiety, and autistic burnout — were highly intercorrelated (r = 0.59–0.73) and all three were strongly associated with Health Interference, QoL, and Overall Health, behaving together as a coherent psychological distress cluster. Within this cluster, depression and anxiety were most closely correlated (r = 0.73), followed by depression and autistic burnout (r = 0.64).

Sets of outcomes we had hypothesized would be moderately correlated all demonstrated correlations between 0.3 and 0.7. The healthcare quality instruments — unmet healthcare needs, barriers to healthcare, patient-provider communication, and both healthcare self-efficacy subscales —intercorrelated within this range. The two healthcare self-efficacy subscales (individual-level and relationship-dependent) were more strongly correlated with each other (r = 0.80) than with the other healthcare quality measures, suggesting they share substantial variance as components of a common self-efficacy construct while also capturing distinct aspects of the healthcare relationship. Adequacy of Supports and Information and Satisfaction with Disability Services were more strongly correlated with each other (r = 0.66) than with other instruments in the toolkit. All Social Support subscales correlated strongly with the Total Social Support scale (r = 0.69–0.83). Self-Determination and Choices and Decisions were moderately correlated (r = 0.54), as expected given their conceptual proximity.

The Choices and Decisions Scale was not statistically correlated with anxiety, total social support, or companionship. Several additional associations were statistically significant but negligible in magnitude (r < 0.04): depression, autistic burnout, unmet healthcare needs, practical support, and lack of isolation. These results suggest that the Choices and Decisions Scale measures a construct independent of mental health, social support, and unmet healthcare needs, and is most closely related to self-determination (r = 0.54).

Consistent with hypotheses, mental health outcomes, community participation, self-determination, and social support all demonstrated moderate to strong correlations with overall health, quality of life, and flourishing. Flourishing showed a broad pattern of associations, correlating strongly not only with QoL and Overall Health but also with Community Participation (r = 0.63) and Self-Determination (r = 0.64), suggesting it captures elements of both health and social engagement.

Within the Social Support subscales, Practical Support had consistently weaker associations with other outcome measures compared to the other Social Support subscales, while Lack of Isolation showed the strongest associations with Community Participation (r = 0.65) and the mental health cluster. Overall, more closely related constructs demonstrated stronger correlations than more distal constructs, consistent with a theoretically coherent structure across the full matrix.

**Table 4** provides the full correlation matrix. The overall pattern of associations was similar across all subgroups, with two exceptions: 1) there was a systematic gradient in the relationship between Choices and Decisions and mental health outcomes across support needs levels, suggesting that external constraints may function differently in groups with differing support needs; and 2) Practical Support showed weaker associations with quality-of-life and engagement outcomes in subgroups with greater reliance on supports and services (DS subcohort, ADL support needs group; see Supplements E1-3).

**Table 4:**
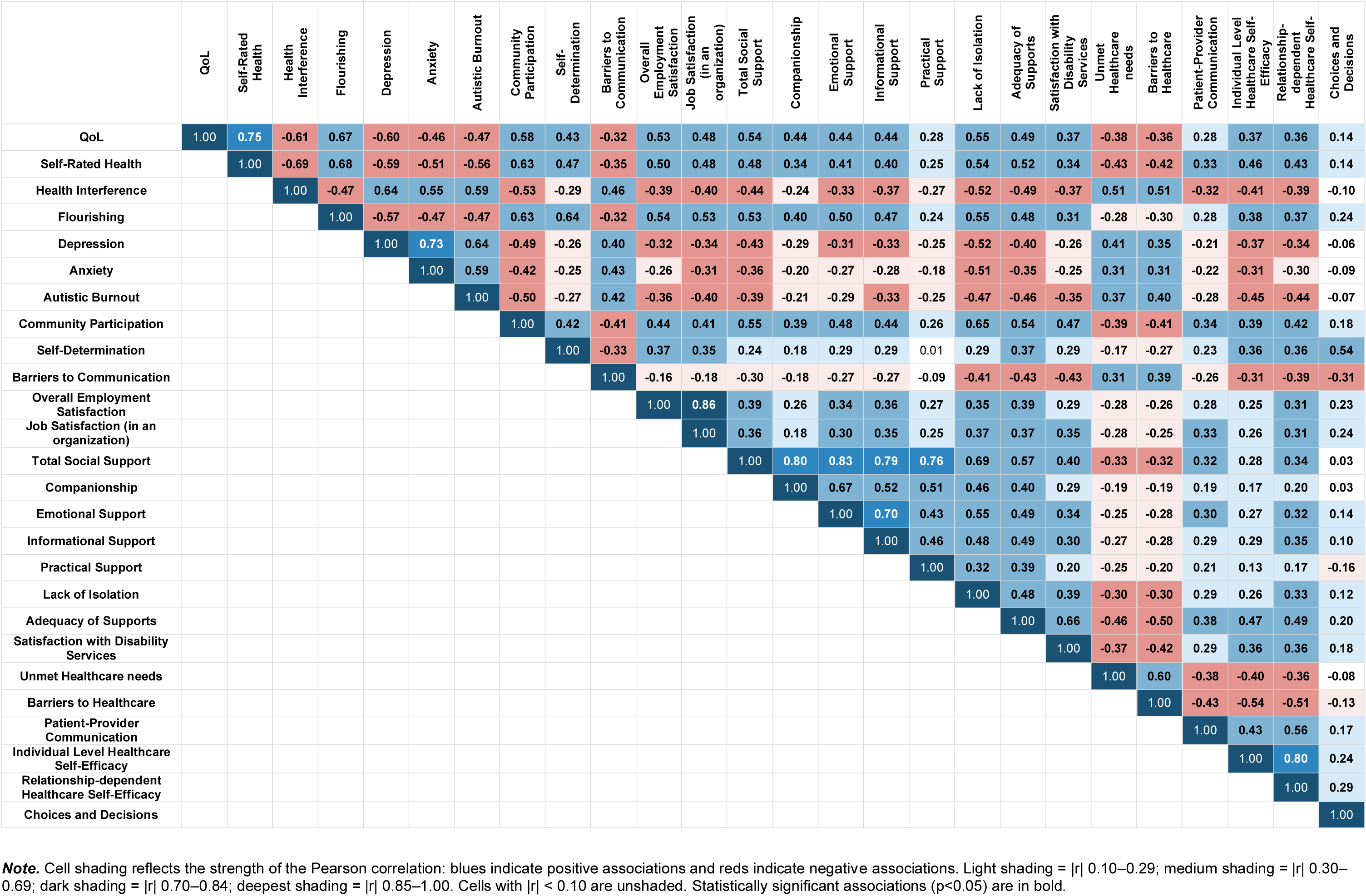
Pairwise correlations between outcome measures.

|  | QoL | Self-Rated Health | Health Interference | Flourishing | Depression | Anxiety | Autistic Burnout | Community Participation | Self-Determination | Barriers to Communication | Overall Employment Satisfaction | Job Satisfaction (in an organization) | Total Social Support | Companionship | Emotional Support | Informational Support | Practical Support | Lack of Isolation | Adequacy of Supports | Satisfaction with Disability Services | Unmet Healthcare needs | Barriers to Healthcare | Patient-Provider Communication | Individual Level Healthcare Self-Efficacy | Relationship-dependent Healthcare Self-Efficacy | Choices and Decisions |
|---|---|---|---|---|---|---|---|---|---|---|---|---|---|---|---|---|---|---|---|---|---|---|---|---|---|---|
| **QoL** | 1.00 | **0.75** | **-0.61** | **0.67** | **-0.60** | **-0.46** | **-0.47** | **0.58** | **0.43** | **-0.32** | **0.53** | **0.48** | **0.54** | **0.44** | **0.44** | **0.44** | **0.28** | **0.55** | **0.49** | **0.37** | **-0.38** | **-0.36** | **0.28** | **0.37** | **0.36** | **0.14** |
| **Self-Rated Health** |  | 1.00 | **-0.69** | **0.68** | **-0.59** | **-0.51** | **-0.56** | **0.63** | **0.47** | **-0.35** | **0.50** | **0.48** | **0.48** | **0.34** | **0.41** | **0.40** | **0.25** | **0.54** | **0.52** | **0.34** | **-0.43** | **-0.42** | **0.33** | **0.46** | **0.43** | **0.14** |
| **Health Interference** |  |  | 1.00 | **-0.47** | **0.64** | **0.55** | **0.59** | **-0.53** | **-0.29** | **0.46** | **-0.39** | **-0.40** | **-0.44** | **-0.24** | **-0.33** | **-0.37** | **-0.27** | **-0.52** | **-0.49** | **-0.37** | **0.51** | **0.51** | **-0.32** | **-0.41** | **-0.39** | **-0.10** |
| **Flourishing** |  |  |  | 1.00 | **-0.57** | **-0.47** | **-0.47** | **0.63** | **0.64** | **-0.32** | **0.54** | **0.53** | **0.53** | **0.40** | **0.50** | **0.47** | **0.24** | **0.55** | **0.48** | **0.31** | **-0.28** | **-0.30** | **0.28** | **0.38** | **0.37** | **0.24** |
| **Depression** |  |  |  |  | 1.00 | **0.73** | **0.64** | **-0.49** | **-0.26** | **0.40** | **-0.32** | **-0.34** | **-0.43** | **-0.29** | **-0.31** | **-0.33** | **-0.25** | **-0.52** | **-0.40** | **-0.26** | **0.41** | **0.35** | **-0.21** | **-0.37** | **-0.34** | **-0.06** |
| **Anxiety** |  |  |  |  |  | 1.00 | **0.59** | **-0.42** | **-0.25** | **0.43** | **-0.26** | **-0.31** | **-0.36** | **-0.20** | **-0.27** | **-0.28** | **-0.18** | **-0.51** | **-0.35** | **-0.25** | **0.31** | **0.31** | **-0.22** | **-0.31** | **-0.30** | **-0.09** |
| **Autistic Burnout** |  |  |  |  |  |  | 1.00 | **-0.50** | **-0.27** | **0.42** | **-0.36** | **-0.40** | **-0.39** | **-0.21** | **-0.29** | **-0.33** | **-0.25** | **-0.47** | **-0.46** | **-0.35** | **0.37** | **0.40** | **-0.28** | **-0.45** | **-0.44** | **-0.07** |
| **Community Participation** |  |  |  |  |  |  |  | 1.00 | **0.42** | **-0.41** | **0.44** | **0.41** | **0.55** | **0.39** | **0.48** | **0.44** | **0.26** | **0.65** | **0.54** | **0.47** | **-0.39** | **-0.41** | **0.34** | **0.39** | **0.42** | **0.18** |
| **Self-Determination** |  |  |  |  |  |  |  |  | 1.00 | **-0.33** | **0.37** | **0.35** | **0.24** | **0.18** | **0.29** | **0.29** | 0.01 | **0.29** | **0.37** | **0.29** | **-0.17** | **-0.27** | **0.23** | **0.36** | **0.36** | **0.54** |
| **Barriers to Communication** |  |  |  |  |  |  |  |  |  | 1.00 | **-0.16** | **-0.18** | **-0.30** | **-0.18** | **-0.27** | **-0.27** | **-0.09** | **-0.41** | **-0.43** | **-0.43** | **0.31** | **0.39** | **-0.26** | **-0.31** | **-0.39** | **-0.31** |
| **Overall Employment Satisfaction** |  |  |  |  |  |  |  |  |  |  | 1.00 | **0.86** | **0.39** | **0.26** | **0.34** | **0.36** | **0.27** | **0.35** | **0.39** | **0.29** | **-0.28** | **-0.26** | **0.28** | **0.25** | **0.31** | **0.23** |
| **Job Satisfaction (in an organization)** |  |  |  |  |  |  |  |  |  |  |  | 1.00 | **0.36** | **0.18** | **0.30** | **0.35** | **0.25** | **0.37** | **0.37** | **0.35** | **-0.28** | **-0.25** | **0.33** | **0.26** | **0.31** | **0.24** |
| **Total Social Support** |  |  |  |  |  |  |  |  |  |  |  |  | 1.00 | **0.80** | **0.83** | **0.79** | **0.76** | **0.69** | **0.57** | **0.40** | **-0.33** | **-0.32** | **0.32** | **0.28** | **0.34** | **0.03** |
| **Companionship** |  |  |  |  |  |  |  |  |  |  |  |  |  | 1.00 | **0.67** | **0.52** | **0.51** | **0.46** | **0.40** | **0.29** | **-0.19** | **-0.19** | **0.19** | **0.17** | **0.20** | **0.03** |
| **Emotional Support** |  |  |  |  |  |  |  |  |  |  |  |  |  |  | 1.00 | **0.70** | **0.43** | **0.55** | **0.49** | **0.34** | **-0.25** | **-0.28** | **0.30** | **0.27** | **0.32** | **0.14** |
| **Informational Support** |  |  |  |  |  |  |  |  |  |  |  |  |  |  |  | 1.00 | **0.46** | **0.48** | **0.49** | **0.30** | **-0.27** | **-0.28** | **0.29** | **0.29** | **0.35** | **0.10** |
| **Practical Support** |  |  |  |  |  |  |  |  |  |  |  |  |  |  |  |  | 1.00 | **0.32** | **0.39** | **0.20** | **-0.25** | **-0.20** | **0.21** | **0.13** | **0.17** | **-0.16** |
| **Lack of Isolation** |  |  |  |  |  |  |  |  |  |  |  |  |  |  |  |  |  | 1.00 | **0.48** | **0.39** | **-0.30** | **-0.30** | **0.29** | **0.26** | **0.33** | **0.12** |
| **Adequacy of Supports** |  |  |  |  |  |  |  |  |  |  |  |  |  |  |  |  |  |  | 1.00 | **0.66** | **-0.46** | **-0.50** | **0.38** | **0.47** | **0.49** | **0.20** |
| **Satisfaction with Disability Services** |  |  |  |  |  |  |  |  |  |  |  |  |  |  |  |  |  |  |  | 1.00 | **-0.37** | **-0.42** | **0.29** | **0.36** | **0.36** | **0.18** |
| **Unmet Healthcare needs** |  |  |  |  |  |  |  |  |  |  |  |  |  |  |  |  |  |  |  |  | 1.00 | **0.60** | **-0.38** | **-0.40** | **-0.36** | **-0.08** |
| **Barriers to Healthcare** |  |  |  |  |  |  |  |  |  |  |  |  |  |  |  |  |  |  |  |  |  | 1.00 | **-0.43** | **-0.54** | **-0.51** | **-0.13** |
| **Patient-Provider Communication** |  |  |  |  |  |  |  |  |  |  |  |  |  |  |  |  |  |  |  |  |  |  | 1.00 | **0.43** | **0.56** | **0.17** |
| **Individual Level Healthcare Self-Efficacy** |  |  |  |  |  |  |  |  |  |  |  |  |  |  |  |  |  |  |  |  |  |  |  | 1.00 | **0.80** | **0.24** |
| **Relationship-dependent Healthcare Self-Efficacy** |  |  |  |  |  |  |  |  |  |  |  |  |  |  |  |  |  |  |  |  |  |  |  |  | 1.00 | **0.29** |
| **Choices and Decisions** |  |  |  |  |  |  |  |  |  |  |  |  |  |  |  |  |  |  |  |  |  |  |  |  |  | 1.00 |

***Note.*** Cell shading reflects the strength of the Pearson correlation: blues indicate positive associations and reds indicate negative associations. Light shading = \|r\| 0.10–0.29; medium shading = \|r\| 0.30–0.69; dark shading = \|r\| 0.70–0.84; deepest shading = \|r\| 0.85–1.00. Cells with \|r\| < 0.10 are unshaded. Statistically significant associations (p<0.05) are in bold.

### Test-retest reliability

**Table 3** shows intraclass coefficients comparing answers between time point 2 and a retest survey administered 2 weeks later in a subset of participants. Test-retest reliability, using one-way models was excellent across all scales (ICC 0.75-0.92), except for Informational Support, that showed good agreement (0.67).^69^ Two-way models were similar, with at a difference in ICCs of at most 0.001. Spearman’s rhos for the single-item and checklist instruments were consistent with primary analyses (ρ = 0.73–0.85, all p < 0.0001).

### Responsiveness to Change

Self-reported change categories were significantly associated with change in outcome scores, in the predicted directions, for almost all scales and sub-scales (**Table 3).** The only exception was the AASPIRE Barriers to Healthcare Checklist (F = 2.58, p = 0.077). Patterns were similar in analyses stratified by each subgroup, though some associations were no longer significant, especially for the small CR group (Supplements F1-4). Wilcoxon signed-rank sensitivity analyses confirmed primary results for the two checklist instruments.

## Discussion

Our community-academic team used a CBPR-nested Delphi process to reach consensus on which outcomes were most important to measure when evaluating the effectiveness of services and interventions for autistic adults.^50^ We then used our CBPR process to collaboratively adapt, revise, or create 19 self-reported survey instruments to measure these outcomes, in addition to 12 modules to measure characteristics that could serve as potential predictors or confounders. This involved a complex 2-year process with multiple meetings, workgroups, and draft iterations. The goal was to ensure the instruments were as relevant and accessible as possible.

The current study suggests that these efforts were successful, as the results provide evidence that the instruments are accessible and have strong initial psychometric characteristics, including internal consistency reliability, test-retest reliability, content validity, convergent and discriminant validity, and responsiveness to change (otherwise known as longitudinal validity), at least for participants taking part in the study directly. We have conducted more in-depth analyses to further assess construct and structural validity for most measures, but due to the detailed instrument-specific nature of the methods and results of such analyses, we will be reporting on them separately.

### Convergent and Discriminant Validity

The correlation matrix provided evidence of convergent and discriminant validity across the 19 outcome measures. All hypothesized patterns were supported, and no unexpected associations emerged that would suggest construct confounding or problematic overlap between instruments.

The strong correlation between the two employment scales (r = 0.86) confirms that the AASPIRE Overall Employment Satisfaction Scale and the adapted Job Satisfaction Survey largely capture the same construct. Researchers may wish to select one based on study context: the Overall Employment Satisfaction Scale may be preferable in settings where employment situations are heterogeneous or where brevity is important, while the Job Satisfaction Survey may be more appropriate for participants employed in formal organizational settings.

The coherence of the mental health cluster — with depression, anxiety, and autistic burnout showing high intercorrelations — is consistent with the well-documented cooccurrence of these conditions in autistic adults.^14,70^ It does not indicate construct redundancy, as each instrument measures a conceptually distinct construct with specific clinical and intervention implications. The strong association of Health Interference with this cluster suggests that health-related functional limitations may be closely intertwined with psychological distress in autistic adults.

The position of Flourishing as a bridge between health and social-functioning outcomes — correlating strongly with both QoL-adjacent measures and with Self-Determination and Community Participation — suggests it may serve as a particularly sensitive and broad indicator of wellbeing in this population. This is consistent with theoretical models that conceptualize flourishing as encompassing social, psychological, and purposive dimensions beyond health alone.^71,72^

The Choices and Decisions Scale’s pattern of negligible associations with mental health, social support, and healthcare needs, despite being statistically detectable in this large sample, suggests it is measuring something distinct: the degree to which an individual has the opportunity and freedom to exercise choice in their daily life. This construct may be more dependent on structural and environmental factors, such as living situation, guardianship status, degree of staff support, and disability characteristics such as communication fluency than on individual psychological states.^73^ Notably, the scale did demonstrate significant responsiveness to change (F = 19.38, p < 0.001), which supports its use as an outcome measure in intervention studies targeting autonomy and self-determination. Additional analyses examining whether Choices and Decisions scores vary by living arrangement, guardianship, support context, or communication fluency support the scale’s construct validity, but are presented in the paper focused specifically on that instrument.^52^

Within the Social Support subscales, the relative weakness of Practical Support associations across the matrix is consistent with the finding in general populations that emotional support is associated with lower depressive symptoms, while instrumental support is not.^74^ The associations are particularly weak in subgroups with greater service use or support needs. We speculate that it may be because practical support is often provided by families, paid staff, or formal systems, and thus may be decoupled from broader social connectedness. The particular strength of Lack of Isolation associations with Community Participation and the mental health cluster is consistent with research linking social isolation to adverse health outcomes in autistic adults,^75^ and suggests this subscale may be especially useful as an outcome target in intervention research.

### Responsiveness to Change

Nearly all outcome measures demonstrated significant external responsiveness to change, with self-perceived change predicting change in scores in the expected directions. The only exception was the Barriers to Healthcare Checklist. Interestingly, when we used a very similar version of that scale in a prior single-arm trial of the AASPIRE Healthcare Toolkit, barriers to healthcare decreased 6-months post intervention, suggesting good internal responsiveness to change in pre-post intervention comparisons.^76^ It is possible that the current lack of responsiveness reflects the smaller magnitude of change in healthcare barriers in the absence of a targeted intervention. The significant responsiveness to change observed for the Choices and Decisions Scale (F = 19.38, p < 0.001) is particularly noteworthy given its constrained correlation profile, as it demonstrates the scale’s sensitivity to meaningful change in autonomy despite its independence from related constructs.

### Role of the Participatory Approach

We believe the good psychometric performance of these instruments reflects the rigor of the participatory process we used to develop them. By centering autistic adults, including non-speaking individuals and people with intellectual disability, throughout all phases of the project, we addressed accessibility concerns typically overlooked in standard instrument development.^40^ The high completion rates and accessibility ratings in the current study, especially in participants taking the survey directly without support, combined with the strong internal consistency, convergent validity, and responsiveness to change, suggest that instruments developed through authentic community partnership can meet or exceed conventional psychometric standards while also being relevant and accessible to the populations they are designed to serve.

### Limitations

While we were pleased to see that efforts to increase accessibility were successful enough for most participants to take part in the study directly, with or without support, that left only a very small number of people using the caregiver report version of the survey. Post-hoc stratified analyses by reporter type showed similar patterns as in the full sample, but many associations were no longer significant in the CR subgroup, presumably due to the small N. We did not include any pairs of direct and caregiver reporters in the longitudinal study to assess concordance of responses. More research is needed to validate the instruments for use by caregivers. Researchers should use caregiver report versions with caution.

We classified participants as DR vs DR-S based on whether they reported that they received support while taking the survey. While overall psychometric properties were relatively similar for the DR and DR-S groups, DR-S participants, not surprisingly, were less likely to say that the survey questions were easy to answer than those who did not need to use support to answer them. Follow-up rates were also lower for the DR-S group. However, results of subgroup analyses stratifying participants by need for support with ADLs and IADLs demonstrated strong accessibility, psychometric properties, and follow-up rates for all three subgroups, suggesting that the instruments may still be used with populations that have higher support needs.

We did not assess criterion validity by comparing results to a reference standard as, in most cases, there is no clear reference standard for the constructs the instruments measure. Future research can compare survey results to assessments made via structured diagnostic interviews for clinical constructs such as depression or anxiety. We did not compare the adapted and original versions of surveys to directly assess whether the adaptations increased accessibility, reliability, or validity.

While we do not expect that the types of adaptation we made would decrease accessibility for general populations, and we do not see a reason why general populations would not be able to use the new measures, we did not test the adapted or de novo instruments with non-autistic participants. We purposefully included autistic people with intellectual disability on our team and in the sample. We expect that these instruments would function similarly, but we have not tested them with non-autistic people with intellectual disability.

*Internal* responsiveness to change is often assessed during intervention trials, where changes in scores over time can be assumed to be attributed to the intervention.^45^ However, using a new instrument to assess the outcome of an intervention would be risky without already knowing that it is responsive to change. As such, we aimed to test *external* responsiveness to change, which is done by comparing changes in scores to a different measure of change^45^ - in our case, participants’ own ratings of change over the past 6 months. We either included a perceived change item for the entire instrument or for each subscale, not both. We had initially intended to collect follow-up data 6 and 12 months after baseline, but to prioritize high follow-up rates, we lengthened the response window. As such, the time-points compared were not always exactly 6 months apart and may not have correlated as well to perceived change over 6 months. Additionally, the checklist format of some instruments may have limited sensitivity to incremental change. However, the significant correlations, in the expected directions, between change scores and self-perceived change in the full sample are promising enough to warrant using them in intervention studies, where researchers can further establish the instruments’ responsiveness to change.

While the heterogeneity of our sample is a strength, the sample skewed younger (slight majority 18-29). As discussed elsewhere,^51^ the HC and DS subcohort had a much greater proportion of younger patients than the community subcohort, presumably due to the age distribution of autistic people formally diagnosed in the healthcare and disability service systems. Given that we only included participants from the US, caution is needed if using these instruments in other countries.

Using a single analytic framework across all instruments prioritized cross-instrument comparability over instrument-specific optimal methods; sensitivity analyses using non-parametric approaches support the robustness of findings for the instruments most affected by this tradeoff.

### Conclusions / Implications

This study provides evidence that the direct report versions of the outcome measures in the AASPIRE Measurement Toolkit have strong initial psychometric properties across a heterogeneous sample. Further research with a larger sample of caregiver reporters is needed. We believe our CBPR approach and close attention to relevance and accessibility greatly contributed to the success of the project. Additional papers focused on more in-depth psychometric analyses for each of the individual measures are in preparation. All instruments are available for others to use, free of charge, at <www.aaspire.org/measurement>. Researchers and service providers can use these instruments to follow outcomes over time and test the effectiveness of services interventions for autistic adults.

## Data Availability

Data will be available in the NIH Data Archive.

https://nda.nih.gov/

## Acknowledgements

We would like to thank Rachel Schuck, Reilly Caldwell, Dominque Soriano, Madeline Proctor, Sullivan Swift, and Janea Jones for their assistance with participant recruitment and management.

## Authorship Confirmation Statement

Conceptualization: C.N., D.M.R., M.B.-E., T.E., S.d.R.R., E.F., W.H.-J., A.J., S.K.K., C.K., J.L.T., J.L., R.K.-L., J.M., K.M., I.M., M.S., Z.S., I.S., A.W., L.-Q.Y.; Methodology: C.N., D.M.R., M.B.-E., S.d.R.R., E.F., W.H.-J., A.J., S.K.K., C.K., J.L.T., J.L., R.K.-L., J.M., K.M., I.M., Z.S., I.S., J.V., A.W., L.-Q.Y.; Investigation: C.N., D.M.R., M.B.-E., W.H.-J., J.L., M.S., J.V., K.J.F.; Data curation: J.V., K.J.F.; Formal analysis: C.N., J.V., N.C.; Funding acquisition: C.N., D.M.R., M.B.-E., W.H.-J., A.J., J.L.T., J.M.; Project administration: C.N., I.M., M.S.; Resources: M.B.-E., W.H.-J., J.L.T., J.L.; Supervision: C.N., M.B.-E., W.H.-J., J.L.T.; Writing – original draft: C.N.; Writing – review & editing: C.N., D.M.R., M.B.-E., T.E., S.d.R.R., E.F., W.H.-J., A.J., S.K.K., C.K., J.L.T., J.L., R.K.-L., J.M., K.M., I.M., M.S., Z.S., I.S., J.V., A.W., L.-Q.Y., K.J.F., N.C. All authors reviewed and approved the final manuscript. We have not published this manuscript elsewhere, and no other journal is currently considering it.

## Availability of data

all data from this study will be available through the National Institutes of Health (NIH) National Data Archive (NDA).

## Ethical considerations

The Institutional Review Board (IRB) at Portland State University (PSU) approved the project and served as the Single IRB (approval # 227674-18); Oregon Health & Science University, Vanderbilt University Medical Center, and San Diego State University ceded review to the PSU IRB. Involvement by the Multnomah County Developmental Disability Services was subsumed under the PSU IRB.

## Consent to participate

Participants or their legally authorized representatives gave written consent online or in-person.

## Disclosure of Artificial Intelligence Use (AI use)

We used AI to assist in the creation and formatting of select tables; to check for errors in coding and verify the accuracy of manually entered tables; and to assist with the compilation of references.

## Supplemental Material

### supplement A: AASPIRE Measurement Toolkit Development Study - Cognitive Interview Guide

As you know, we are putting together a set of survey measures to evaluate services for autistic adults.

We are trying to make sure that our survey measures are easy to understand and that they ask about things that are important.

We’re going to look at [# OF SURVEYS] from the link. We’ll start with the one you see here, [IDENTIFY SURVEY TO ORIENT]

- For each one, think about what it would be like if you took a survey that included these questions.
- Only think about what it would be like for **you** – don’t worry about how other autistic people may answer these questions.

*<Repeat interview questions for each pre-selected instrument for a total of 3-6 instruments, depending on how many instruments the participant can review in approximately 60-120 minutes.>*

I would like to hear your opinions on the instructions for this set of questions. <*Show or read instructions.>*

1. Overall, what do you think about these instructions? <*If comments are negative, probe further. For example,>*

a. Please tell me more about that.
b. What do you feel makes the instructions <*negative phrase/comment used by participant*>?

I would like to hear your opinions on this set of questions. *<Show or read set of questions.>*

1. Overall, what do you think about this set of questions? *<If comments are negative, probe further. For example,>*

a. Please tell me more about that.
b. What do you feel makes this set of questions *<negative phrase/comment used by participant>?*
2. In general, how easy would it be for you to answer these questions? *<If participant has said the questions would be hard to answer, probe further. For example,>*

a. What makes it hard to for you to answer these questions?
b. Are there any particular questions that are hard to answer? If so, which ones?
c. In your own words, what do you think that question is asking?
d. What would make it easier for you to answer these questions?
e. What if we *<suggest idea for possible way to address the problem>*? Would that help?

Now let’s talk a little bit more about some of the **individual** questions. Look at question number [#].

*<Show or read specific item. Repeat for one or more items that were pre-selected for this measure. No need to repeat items that the person already talked about in depth in interview question 2 above.>*

3. Please tell me, in your own words, what you think this question is asking.
4. How would you answer this question? Please give me an example of what you mean by your answer to this question. *<If paraphrase or example does not match intended meaning, probe further.>* Thanks, that’s really helpful! It looks like we may need to do some more work to improve this question. We want to ask participants about *<TRY to rephrase the question to distinguish intended construct from paraphrase>*.

a. How can we change this question to make it easier to understand?
b. What if we asked *<offer possible change>*? Would that help?

Thanks! That’s really helpful! Now please think about the **full set** of questions again.

5. Do you think these questions are asking about something that is important?
6. Are there other important things about *<TOPIC being covered by instrument>* that we should ask about?
7. Do you have any other comments or suggestions?

*<If still have time to review more instruments>*

Thanks so much. That was great. If it’s ok with you, let’s look at another set of questions.

*<Repeat interview guide for next instrument.>*

*<If done>*

Thank you very much for your thoughts and recommendations. We really appreciate it!

**Supplement B1:** Participant Characteristics by Subcohort.

|  |  | Total<br>N = 870 | Disability Services<br>N = 265 | Healthcare<br>N = 319 | Community<br>N = 285 |
|---|---|---|---|---|---|
| **Study Participation** |  |  |  |  |  |
| Reporter Type | Direct Report | 605 (70%) | 134 (50%) | 204 (64%) | 267 (94%) |
|  | Direct Report-Supported | 194 (22%) | 102 (38%) | 76 (24%) | 16 (7%) |
|  | Caregiver Report | 71 (8%) | 30 (11%) | 39 (12%) | 2 (1%) |
| **Demographic Characteristics** |  |  |  |  |  |
| Age group | 18–29 | 505 (58%) | 186 (70%) | 212 (67%) | 107 (38%) |
|  | 30–39 | 200 (23%) | 61 (23%) | 56 (18%) | 83 (29%) |
|  | 40–49 | 93 (11%) | 12 (5%) | 27 (8%) | 54 (19%) |
|  | 50+ | 71 (8%) | 6 (2%) | 24 (8%) | 41 (14%) |
| BIPOC<sup>1</sup> | Yes | 244 (30%) | 97 (38%) | 98 (34%) | 49 (18%) |
| Race/ethnicity | Asian | 35 (4%) | 15 (6%) | 13 (4%) | 7 (3%) |
|  | Biracial / multi-racial | 30 (4%) | 9 (4%) | 14 (5%) | 7 (3%) |
|  | Black, African American, or of African ancestry | 31 (4%) | 11 (4%) | 15 (5%) | 5 (2%) |
|  | Native American or Native Alaskan | 15 (2%) | 3 (1%) | 11 (4%) | 1 (0%) |
|  | Native Hawaiian or other Pacific Islander | 1 (0%) | 1 (0%) | 0 (0%) | 0 (0%) |
|  | White | 607 (73%) | 163 (64%) | 214 (72%) | 230 (84%) |
|  | Hispanic | 97 (12%) | 47 (18%) | 29 (10%) | 21 (8%) |
|  | Other | 12 (1%) | 6 (2%) | 2 (1%) | 4 (1%) |
| Gender | Female | 346 (40%) | 77 (29%) | 121 (38%) | 148 (52%) |
|  | Male | 393 (45%) | 170 (64%) | 154 (48%) | 69 (24%) |
|  | Non-binary / agender / genderfluid | 267 (31%) | 50 (19%) | 80 (25%) | 137 (48%) |
| Sex assigned at birth | Male | 404 (49%) | 180 (70%) | 153 (51%) | 71 (25%) |
|  | Female | 427 (51%) | 75 (29%) | 144 (48%) | 208 (75%) |
|  | Other | 2 (0%) | 1 (0%) | 1 (0%) | 0 (0%) |
| Gender same as SAB<sup>2</sup> | Yes | 616 (74%) | 226 (89%) | 231 (78%) | 159 (57%) |
| LGBTQIA+<sup>3</sup> | Yes | 404 (53%) | 72 (33%) | 135 (50%) | 197 (72%) |
| Educational attainment | Less than high school | 59 (7%) | 27 (11%) | 30 (10%) | 2 (1%) |
|  | Modified HS diploma | 92 (11%) | 44 (18%) | 46 (16%) | 2 (1%) |
|  | Regular HS diploma | 116 (14%) | 51 (20%) | 49 (17%) | 16 (6%) |
|  | Some college | 201 (25%) | 86 (35%) | 75 (25%) | 40 (15%) |
|  | Associates degree / certificate | 84 (10%) | 24 (10%) | 29 (10%) | 31 (11%) |
|  | Bachelors degree | 148 (18%) | 15 (6%) | 45 (15%) | 88 (32%) |
|  | Graduate degree | 120 (15%) | 2 (1%) | 22 (7%) | 96 (35%) |
| Received Special Education services<sup>4</sup> | Yes | 579 (70%) | 231 (90%) | 228 (78%) | 120 (44%) |
| Living situation | Own home (including student housing) | 384 (46%) | 57 (22%) | 122 (41%) | 205 (74%) |
|  | Family's home | 379 (45%) | 158 (61%) | 155 (52%) | 66 (24%) |
|  | Group home, facility, unhoused, or other<sup>5</sup> | 72 (9%) | 43 (17%) | 21 (7%) | 8 (3%) |
| Employed for pay | Yes | 420 (50%) | 90 (36%) | 137 (45%) | 193 (69%) |
| Financial hardship | Very hard | 170 (20%) | 51 (20%) | 63 (21%) | 56 (20%) |
|  | Somewhat hard | 275 (33%) | 71 (28%) | 99 (33%) | 105 (38%) |
|  | Not hard at all | 387 (47%) | 134 (52%) | 136 (46%) | 117 (42%) |
| **Disability Characteristics** |  |  |  |  |  |
| Autism diagnosis<sup>6</sup> | Formal (medical or educational) | 787 (92%) | 266 (100%) | 319 (100%) | 202 (75%) |
|  | Self-diagnosed | 69 (8%) | 0 (0%) | 0 (0%) | 69 (26%) |
| Co-occurring ADHD | Yes (formal or self-diagnosed) | 460 (55%) | 127 (49%) | 167 (56%) | 166 (60%) |
| Support needs<sup>7</sup> | Needs assistance with ADLs | 221 (26%) | 73 (28%) | 88 (30%) | 60 (22%) |
|  | Needs assistance with IADLs only | 371 (44%) | 120 (47%) | 127 (43%) | 124 (44%) |
|  | Does not need assistance | 243 (29%) | 65 (25%) | 83 (28%) | 95 (34%) |
| **Communication Characteristics** |  |  |  |  |  |
| Understand speech under daily conditions<sup>8</sup> | No difficulty | 325 (39%) | 108 (42%) | 112 (37%) | 105 (38%) |
|  | Fairly well | 414 (49%) | 130 (50%) | 141 (47%) | 143 (51%) |
|  | Difficult or minimal | 101 (12%) | 21 (8%) | 49 (16%) | 31 (11%) |
| Speak under daily conditions<sup>8</sup> | No difficulty | 214 (25%) | 81 (31%) | 76 (25%) | 57 (20%) |
|  | Fairly well | 376 (45%) | 111 (43%) | 115 (38%) | 150 (54%) |
|  | Difficult or minimal | 250 (30%) | 66 (26%) | 111 (37%) | 73 (26%) |
| Reading level | Advanced | 627 (75%) | 157 (61%) | 207 (68%) | 263 (94%) |
|  | Basic (e.g., uses "Plain Language" materials) | 143 (17%) | 73 (28%) | 55 (18%) | 15 (5%) |
|  | Minimal or none | 71 (8%) | 29 (11%) | 41 (14%) | 1 (0%) |
| Writing level | Advanced | 563 (67%) | 128 (49%) | 181 (60%) | 254 (91%) |
|  | Basic (e.g., can explain simple ideas) | 192 (23%) | 98 (38%) | 72 (24%) | 22 (8%) |
|  | Minimal or none | 87 (10%) | 35 (13%) | 49 (16%) | 3 (1%) |
| Disability service use | Receives disability services | 270 (43%) | 176 (75%) | 68 (34%) | 26 (14%) |
|  | Needs but does not receive | 56 (9%) | 10 (4%) | 25 (12%) | 21 (11%) |
|  | Neither needs nor receives | 215 (35%) | 18 (8%) | 84 (41%) | 113 (62%) |
|  | Unknown / other | 80 (13%) | 31 (13%) | 26 (13%) | 23 (13%) |
| **Health Status and Co-occurring Conditions** |  |  |  |  |  |
| Health status | Poor | 69 (8%) | 10 (4%) | 31 (10%) | 28 (10%) |
|  | Fair | 205 (25%) | 44 (17%) | 74 (25%) | 87 (31%) |
|  | Good | 297 (36%) | 93 (36%) | 107 (36%) | 97 (35%) |
|  | Very good | 176 (21%) | 61 (24%) | 61 (21%) | 54 (19%) |
|  | Excellent | 85 (10%) | 49 (19%) | 23 (8%) | 13 (5%) |
| Other mental health conditions<sup>9</sup> | Depression | 374 (45%) | 96 (37%) | 139 (47%) | 139 (50%) |
|  | Anxiety | 525 (63%) | 142 (55%) | 187 (63%) | 196 (70%) |
|  | PTSD | 229 (27%) | 39 (15%) | 84 (28%) | 106 (38%) |
|  | Bipolar | 60 (7%) | 18 (7%) | 29 (10%) | 13 (5%) |
|  | Schizophrenia or psychosis | 21 (3%) | 7 (3%) | 12 (4%) | 2 (1%) |
| Any mental health condition | Yes | 616 (75%) | 166 (66%) | 221 (76%) | 229 (83%) |
| Number of physical conditions | 0 | 321 (38%) | 141 (55%) | 108 (36%) | 72 (26%) |
|  | 1 | 179 (21%) | 63 (24%) | 63 (21%) | 53 (19%) |
|  | 2 or more | 335 (40%) | 54 (21%) | 127 (43%) | 154 (55%) |

<sup>1</sup> Black, Indigenous, and People of Color
<sup>2</sup> Sex Assigned at Birth
<sup>3</sup> Lesbian, Gay, Bisexual, Transgender, Queer, Intersex, Asexual, plus others.
<sup>4</sup> Including an IEP, a 504 plan, or other special education services.
<sup>5</sup> Including foster homes, adult care homes, residential treatment centers, long-term care, emergency or transitional housing, institutional housing, and buildings not meant for housing.
<sup>6</sup> Formal medical or educational diagnosis vs. self-diagnosis of autism, ASD, Asperger's disorder, or PDD-NOS.
<sup>7</sup> Needs assistance with ADLs = requires support for basic self-care; Needs assistance with IADLs only = requires support for instrumental activities but not basic self-care; Does not need assistance = rarely or never requires support for either.
<sup>8</sup> Daily conditions, including when the environment may be disruptive or overstimulating, and when a person may feel tired, hungry, or unwell.
<sup>9</sup> Other than autism or ADHD. Column totals may exceed 100% as multiple conditions were permitted.
Note: N and % for follow-up reflect participants who completed at least part of each survey, as a percentage of baseline N for that subcohort. Percentages for gender, race/ethnicity, and mental health condition items may sum to >100% as multiple responses were permitted. All other percentages are column percentages. Total N varies across rows due to missing data.

**Supplemental Table B2:** Participant Characteristics by Support Needs.

|  |  | **Total<br>N=870** | **No Assistance Needed<br>N=243** | **IADLs Only<br>N=371** | **ADLs<br>N=221** |
|---|---|---|---|---|---|
| **Study Participation** |  |  |  |  |  |
| Reporter type | Direct Report (DR) | 605 (70%) | 226 (93%) | 255 (69%) | 104 (47%) |
|  | DR – Supported (DR-S) | 194 (22%) | 17 (7%) | 105 (28%) | 60 (27%) |
|  | Caregiver Report (CR) | 71 (8%) | 0 (0%) | 11 (3%) | 57 (26%) |
| Subcohort | Disability Services (DDS) | 266 (31%) | 65 (27%) | 120 (32%) | 73 (33%) |
|  | Healthcare | 319 (37%) | 83 (34%) | 127 (34%) | 88 (40%) |
|  | Community | 285 (33%) | 95 (39%) | 124 (33%) | 60 (27%) |
| **Demographic Characteristics** |  |  |  |  |  |
| Age group | 18–29 | 505 (58%) | 110 (45%) | 217 (58%) | 153 (69%) |
|  | 30–39 | 200 (23%) | 55 (23%) | 87 (23%) | 52 (23%) |
|  | 40–49 | 93 (11%) | 36 (15%) | 47 (13%) | 8 (4%) |
|  | 50+ | 71 (8%) | 42 (17%) | 19 (5%) | 8 (4%) |
| BIPOC<sup>1</sup> | Yes | 244 (30%) | 64 (27%) | 106 (29%) | 74 (33%) |
| Race/ethnicity | Asian | 35 (4%) | 8 (3%) | 16 (4%) | 11 (5%) |
|  | Biracial / multi-racial | 30 (4%) | 10 (4%) | 9 (2%) | 11 (5%) |
|  | Black, African American, or of African ancestry | 31 (4%) | 7 (3%) | 13 (4%) | 11 (5%) |
|  | Native American or Native Alaskan | 15 (2%) | 3 (1%) | 6 (2%) | 6 (3%) |
|  | Native Hawaiian or other Pacific Islander | 1 (0%) | 1 (0%) | 0 (0%) | 0 (0%) |
|  | White | 607 (73%) | 183 (76%) | 276 (75%) | 148 (67%) |
|  | Hispanic | 97 (12%) | 25 (10%) | 43 (12%) | 29 (13%) |
|  | Other | 12 (1%) | 3 (1%) | 6 (2%) | 3 (1%) |
| Gender | Female | 346 (40%) | 90 (37%) | 176 (47%) | 80 (36%) |
|  | Male | 393 (45%) | 135 (56%) | 151 (41%) | 107 (48%) |
|  | Non-binary / agender / genderfluid | 267 (31%) | 64 (26%) | 124 (33%) | 79 (35%) |
| Sex assigned at birth | Male | 404 (49%) | 140 (58%) | 156 (42%) | 108 (49%) |
|  | Female | 427 (51%) | 102 (42%) | 213 (57%) | 112 (50%) |
|  | Other | 2 (0%) | 1 (0%) | 1 (0%) | 0 (0%) |
| Gender same as SAB<sup>2</sup> | Yes | 616 (74%) | 194 (80%) | 272 (74%) | 150 (68%) |
| LGBTQIA+<sup>3</sup> | Yes | 404 (53%) | 100 (44%) | 200 (58%) | 104 (54%) |
| Educational attainment | Less than high school | 59 (7%) | 5 (2%) | 20 (5%) | 34 (16%) |
|  | Modified HS diploma | 92 (11%) | 5 (2%) | 43 (12%) | 44 (20%) |
|  | Regular HS diploma | 116 (14%) | 39 (16%) | 53 (15%) | 24 (11%) |
|  | Some college | 201 (25%) | 54 (22%) | 94 (26%) | 53 (24%) |
|  | Associates degree / certificate | 84 (10%) | 29 (12%) | 35 (10%) | 20 (9%) |
|  | Bachelors degree | 148 (18%) | 55 (23%) | 73 (20%) | 20 (9%) |
|  | Graduate degree | 120 (15%) | 53 (22%) | 45 (12%) | 22 (10%) |
| Received Special Education services<sup>4</sup> | Yes | 579 (70%) | 143 (60%) | 257 (70%) | 179 (81%) |
| Living situation | Own home (including student housing) | 384 (46%) | 150 (62%) | 158 (42%) | 76 (34%) |
|  | Family's home | 379 (45%) | 80 (33%) | 172 (46%) | 127 (57%) |
|  | Group home, facility, unhoused, or other<sup>5</sup> | 72 (9%) | 13 (5%) | 41 (11%) | 18 (8%) |
| Employed for pay | Yes | 420 (50%) | 155 (65%) | 195 (55%) | 59 (28%) |
| Financial hardship | Very hard | 170 (20%) | 23 (10%) | 82 (22%) | 65 (29%) |
|  | Somewhat hard | 275 (33%) | 69 (29%) | 136 (37%) | 70 (32%) |
|  | Not hard at all | 387 (47%) | 150 (62%) | 152 (41%) | 85 (38%) |
| **Disability Characteristics** |  |  |  |  |  |
| Autism diagnosis<sup>6</sup> | Formal (medical or educational) | 787 (92%) | 218 (90%) | 336 (91%) | 204 (92%) |
|  | Self-diagnosed | 69 (8%) | 23 (10%) | 31 (8%) | 15 (7%) |
| Co-occurring ADHD | Yes (formal or self-diagnosed) | 460 (55%) | 122 (50%) | 217 (58%) | 121 (55%) |
| **Communication Characteristics** |  |  |  |  |  |
| Understand speech under daily conditions<sup>8</sup> | No difficulty | 325 (39%) | 134 (55%) | 143 (39%) | 46 (21%) |
|  | Fairly well | 414 (49%) | 91 (38%) | 190 (51%) | 126 (57%) |
|  | Difficult or minimal | 101 (12%) | 17 (7%) | 36 (10%) | 48 (22%) |
| Speak under daily conditions<sup>8</sup> | No difficulty | 214 (25%) | 93 (38%) | 93 (25%) | 27 (12%) |
|  | Fairly well | 376 (45%) | 118 (49%) | 179 (49%) | 76 (34%) |
|  | Difficult or minimal | 250 (30%) | 32 (13%) | 96 (26%) | 117 (53%) |
| Reading level | Advanced | 627 (75%) | 215 (89%) | 293 (79%) | 116 (52%) |
|  | Basic (e.g., uses "Plain Language" materials) | 143 (17%) | 19 (8%) | 65 (18%) | 54 (24%) |
|  | Minimal or none | 71 (8%) | 7 (3%) | 12 (3%) | 51 (23%) |
| Writing level | Advanced | 563 (67%) | 205 (84%) | 259 (70%) | 97 (44%) |
|  | Basic (e.g., can explain simple ideas) | 192 (23%) | 32 (13%) | 98 (26%) | 59 (27%) |
|  | Minimal or none | 87 (10%) | 6 (2%) | 14 (4%) | 63 (29%) |
| Disability service use | Receives disability services | 270 (43%) | 39 (21%) | 127 (49%) | 104 (60%) |
|  | Needs but does not receive | 56 (9%) | 9 (5%) | 23 (9%) | 24 (14%) |
|  | Neither needs nor receives | 215 (35%) | 120 (63%) | 78 (30%) | 17 (10%) |
|  | Unknown / other | 80 (13%) | 21 (11%) | 32 (12%) | 27 (16%) |
| **Health Status and Co-occurring Conditions** |  |  |  |  |  |
| Health status | Poor | 69 (8%) | 12 (5%) | 26 (7%) | 31 (14%) |
|  | Fair | 205 (25%) | 45 (19%) | 103 (28%) | 57 (26%) |
|  | Good | 297 (36%) | 77 (32%) | 146 (39%) | 74 (33%) |
|  | Very good | 176 (21%) | 77 (32%) | 62 (17%) | 37 (17%) |
|  | Excellent | 85 (10%) | 31 (13%) | 33 (9%) | 21 (9%) |
| Other mental health conditions<sup>9</sup> | Depression | 374 (45%) | 93 (38%) | 186 (50%) | 95 (43%) |
|  | Anxiety | 525 (63%) | 134 (55%) | 255 (69%) | 136 (61%) |
|  | PTSD | 229 (27%) | 44 (18%) | 116 (31%) | 69 (31%) |
|  | Bipolar | 60 (7%) | 15 (6%) | 27 (7%) | 18 (8%) |
|  | Schizophrenia or psychosis | 21 (3%) | 7 (3%) | 10 (3%) | 4 (2%) |
| Any mental health condition | Yes | 616 (75%) | 158 (66%) | 297 (82%) | 161 (74%) |
| Number of physical conditions | 0 | 321 (38%) | 115 (47%) | 138 (37%) | 68 (30%) |
|  | 1 | 179 (21%) | 55 (23%) | 83 (22%) | 41 (18%) |
|  | 2 or more | 335 (40%) | 73 (30%) | 150 (40%) | 112 (50%) |

<sup>1</sup> Black, Indigenous, and People of Color
<sup>2</sup> Sex Assigned at Birth
<sup>3</sup> Lesbian, Gay, Bisexual, Transgender, Queer, Intersex, Asexual, plus others.
<sup>4</sup> Including an IEP, a 504 plan, or other special education services.
<sup>5</sup> Including foster homes, adult care homes, residential treatment centers, long-term care, emergency or transitional housing, institutional housing, and buildings not meant for housing.
<sup>6</sup> Formal medical or educational diagnosis vs. self-diagnosis of autism, ASD, Asperger's disorder, or PDD-NOS.
<sup>7</sup> Needs assistance with ADLs = requires support for basic self-care activities; Needs assistance with IADLs only = requires support for instrumental activities but not basic self-care; Does not need assistance = rarely or never requires either.
<sup>8</sup> Daily conditions, including when the environment may be disruptive or overstimulating, and when a person may feel tired, hungry, or unwell.
<sup>9</sup> Other than autism or ADHD. Column totals may exceed 100% as multiple conditions were permitted.
Note: Percentages for gender, race/ethnicity, and mental health condition items may sum to >100% as multiple responses were permitted. All other percentages are column percentages within each group. Total N varies across rows due to missing data.

**Supplemental Table B3:** Participant Characteristics by Reporter Type.

| Characteristic |  | Total N=870 | Direct Report (DR) N=605 | DR – Supported (DR-S) N=194 | Caregiver Report (CR) N=71 |
|---|---|---|---|---|---|
| **Study Participation** |  |  |  |  |  |
| Subcohort | Disability Services (DDS) | 266 (31%) | 134 (22%) | 102 (53%) | 30 (42%) |
|  | Healthcare | 319 (37%) | 204 (34%) | 76 (39%) | 39 (55%) |
|  | Community | 285 (33%) | 267 (44%) | 16 (8%) | 2 (3%) |
| **Demographic Characteristics** |  |  |  |  |  |
| Age group | 18–29 | 505 (58%) | 293 (48%) | 155 (81%) | 57 (78%) |
|  | 30–39 | 200 (23%) | 165 (27%) | 22 (12%) | 13 (18%) |
|  | 40–49 | 93 (11%) | 83 (14%) | 8 (4%) | 2 (3%) |
|  | 50+ | 71 (8%) | 64 (11%) | 6 (3%) | 1 (1%) |
| BIPOC<sup>1</sup> | Yes | 244 (30%) | 156 (27%) | 62 (35%) | 26 (37%) |
| Race/ethnicity | Asian | 35 (4%) | 20 (3%) | 9 (5%) | 6 (9%) |
|  | Biracial / multi-racial | 30 (4%) | 18 (3%) | 6 (3%) | 6 (9%) |
|  | Black, African American, or of African ancestry | 31 (4%) | 13 (2%) | 11 (6%) | 7 (10%) |
|  | Native American or Native Alaskan | 15 (2%) | 11 (2%) | 4 (2%) | 0 (0%) |
|  | Native Hawaiian or other Pacific Islander | 1 (0%) | 1 (0%) | 0 (0%) | 0 (0%) |
|  | White | 607 (73%) | 440 (76%) | 121 (68%) | 46 (66%) |
|  | Hispanic | 97 (12%) | 65 (11%) | 27 (15%) | 5 (7%) |
|  | Other | 12 (1%) | 11 (2%) | 1 (1%) | 0 (0%) |
| Gender | Female | 346 (40%) | 266 (44%) | 69 (36%) | 11 (15%) |
|  | Male | 393 (45%) | 234 (39%) | 100 (52%) | 59 (81%) |
|  | Non-binary / agender / genderfluid | 267 (31%) | 232 (38%) | 35 (18%) | 0 (0%) |
| Sex assigned at birth | Male | 404 (49%) | 243 (42%) | 102 (57%) | 59 (84%) |
|  | Female | 427 (51%) | 340 (58%) | 76 (42%) | 11 (16%) |
|  | Other | 2 (0%) | 1 (0%) | 1 (1%) | 0 (0%) |
| Gender same as SAB<sup>2</sup> | Yes | 616 (74%) | 389 (67%) | 158 (89%) | 69 (100%) |
| LGBTQIA+<sup>3</sup> | Yes | 404 (53%) | 349 (63%) | 49 (32%) | 6 (12%) |
| Educational attainment | Less than high school | 59 (7%) | 20 (3%) | 15 (9%) | 24 (35%) |
|  | Modified HS diploma | 92 (11%) | 19 (3%) | 39 (22%) | 34 (50%) |
|  | Regular HS diploma | 116 (14%) | 70 (12%) | 41 (23%) | 5 (7%) |
|  | Some college | 201 (25%) | 141 (24%) | 55 (31%) | 5 (7%) |
|  | Associates degree / certificate | 84 (10%) | 72 (13%) | 12 (7%) | 0 (0%) |
|  | Bachelors degree | 148 (18%) | 138 (24%) | 10 (6%) | 0 (0%) |
|  | Graduate degree | 120 (15%) | 116 (20%) | 4 (2%) | 0 (0%) |
| Received Special Education services<sup>4</sup> | Yes | 579 (70%) | 348 (60%) | 161 (90%) | 70 (100%) |
| Living situation | Own home (including student housing) | 384 (46%) | 360 (62%) | 24 (13%) | 0 (0%) |
|  | Family's home | 379 (45%) | 185 (32%) | 133 (74%) | 61 (87%) |
|  | Group home, facility, unhoused, or other<sup>5</sup> | 72 (9%) | 40 (7%) | 23 (13%) | 9 (13%) |
| Employed for pay | Yes | 420 (50%) | 354 (60%) | 58 (33%) | 8 (12%) |
| Financial hardship | Very hard | 170 (20%) | 121 (21%) | 34 (19%) | 15 (21%) |
|  | Somewhat hard | 275 (33%) | 208 (36%) | 56 (31%) | 11 (16%) |
|  | Not hard at all | 387 (47%) | 254 (44%) | 89 (50%) | 44 (63%) |
| **Disability Characteristics** |  |  |  |  |  |
| Autism diagnosis<sup>6</sup> | Formal (medical or educational) | 787 (92%) | 528 (89%) | 187 (98%) | 72 (100%) |
|  | Self-diagnosed | 69 (8%) | 65 (11%) | 4 (2%) | 0 (0%) |
| Co-occurring ADHD | Yes (formal or self-diagnosed) | 460 (55%) | 344 (59%) | 94 (53%) | 22 (31%) |
| Support needs<sup>7</sup> | Needs assistance with ADLs | 221 (26%) | 104 (18%) | 60 (33%) | 57 (84%) |
|  | Needs assistance with IADLs only | 371 (44%) | 255 (44%) | 105 (58%) | 11 (16%) |
|  | Does not need assistance | 243 (29%) | 226 (39%) | 17 (9%) | 0 (0%) |
| **Communication Characteristics** |  |  |  |  |  |
| Understand speech under daily conditions<sup>8</sup> | No difficulty | 325 (39%) | 241 (41%) | 76 (41%) | 8 (11%) |
|  | Fairly well | 414 (49%) | 295 (50%) | 81 (44%) | 38 (54%) |
|  | Difficult or minimal | 101 (12%) | 50 (9%) | 27 (15%) | 24 (34%) |
| Speak under daily conditions<sup>8</sup> | No difficulty | 214 (25%) | 156 (27%) | 56 (30%) | 2 (3%) |
|  | Fairly well | 376 (45%) | 296 (51%) | 65 (35%) | 15 (21%) |
|  | Difficult or minimal | 250 (30%) | 134 (23%) | 63 (34%) | 53 (76%) |
| Reading level | Advanced | 627 (75%) | 525 (89%) | 95 (52%) | 7 (10%) |
|  | Basic (e.g., uses "Plain Language" materials) | 143 (17%) | 55 (9%) | 64 (35%) | 24 (34%) |
|  | Minimal or none | 71 (8%) | 7 (1%) | 24 (13%) | 40 (56%) |
| Writing level | Advanced | 563 (67%) | 486 (83%) | 72 (39%) | 5 (7%) |
|  | Basic (e.g., can explain simple ideas) | 192 (23%) | 94 (16%) | 81 (44%) | 17 (24%) |
|  | Minimal or none | 87 (10%) | 6 (1%) | 32 (17%) | 49 (69%) |
| Disability service use | Receives disability services | 270 (43%) | 120 (29%) | 95 (67%) | 55 (85%) |
|  | Needs but does not receive | 56 (9%) | 39 (9%) | 13 (9%) | 4 (6%) |
|  | Neither needs nor receives | 215 (35%) | 199 (48%) | 16 (11%) | 0 (0%) |
|  | Unknown / other | 80 (13%) | 56 (14%) | 18 (13%) | 6 (9%) |
| **Health Status and Co-occurring Conditions** |  |  |  |  |  |
| Health status | Poor | 69 (8%) | 59 (10%) | 7 (4%) | 3 (4%) |
|  | Fair | 205 (25%) | 162 (28%) | 36 (20%) | 7 (10%) |
|  | Good | 297 (36%) | 205 (35%) | 64 (36%) | 28 (40%) |
|  | Very good | 176 (21%) | 114 (20%) | 46 (26%) | 16 (23%) |
|  | Excellent | 85 (10%) | 43 (7%) | 26 (15%) | 16 (23%) |
| Other mental health conditions<sup>9</sup> | Depression | 374 (45%) | 307 (52%) | 62 (34%) | 5 (7%) |
|  | Anxiety | 525 (63%) | 391 (67%) | 108 (60%) | 26 (37%) |
|  | PTSD | 229 (27%) | 196 (34%) | 30 (17%) | 3 (4%) |
|  | Bipolar | 60 (7%) | 49 (8%) | 10 (6%) | 1 (1%) |
|  | Schizophrenia or psychosis | 21 (3%) | 13 (2%) | 7 (4%) | 1 (1%) |
| Any mental health condition | Yes | 616 (75%) | 461 (80%) | 122 (69%) | 33 (50%) |
| Number of physical conditions | 0 | 321 (38%) | 203 (35%) | 86 (48%) | 32 (46%) |
|  | 1 | 179 (21%) | 117 (20%) | 40 (22%) | 22 (31%) |
|  | 2 or more | 335 (40%) | 265 (45%) | 54 (30%) | 16 (23%) |

<sup>1</sup> Black, Indigenous, and People of Color
<sup>2</sup> Sex Assigned at Birth
<sup>3</sup> Lesbian, Gay, Bisexual, Transgender, Queer, Intersex, Asexual, plus others.
<sup>4</sup> Including an IEP, a 504 plan, or other special education services.
<sup>5</sup> Including foster homes, adult care homes, residential treatment centers, long-term care, emergency or transitional housing, institutional housing, and buildings not meant for housing.
<sup>6</sup> Formal medical or educational diagnosis vs. self-diagnosis of autism, ASD, Asperger's disorder, or PDD-NOS.
<sup>7</sup> Needs assistance with ADLs = requires support for basic self-care activities; Needs assistance with IADLs only = requires support for instrumental activities but not basic self-care; Does not need assistance = rarely or never requires either.
<sup>8</sup> Daily conditions, including when the environment may be disruptive or overstimulating, and when a person may feel tired, hungry, or unwell.
<sup>9</sup> Other than autism or ADHD. Column totals may exceed 100% as multiple conditions were permitted.
Note: Percentages for gender, race/ethnicity, and mental health condition items may sum to >100% as multiple responses were permitted. All other percentages are column percentages within each group. Total N varies across rows due to missing data.

**Supplemental Table B4:** Demographic Characteristics of Caregiver Reporters (N = 62)

| **Characteristic** | **N (%)** |
|---|---|
| **Age** |  |
| Mean (SD) | 55.8 (8.1) |
| Range | 39–75 |
| **Gender<sup>1</sup>** |  |
| Woman | 55 (89%) |
| Man | 7 (11%) |
| Non-binary or other gender identity | 0 (0%) |
| **Race/ethnicity<sup>1</sup>** |  |
| Asian | 3 (5%) |
| Black, African, or African American | 7 (11%) |
| Latino/a/x or Hispanic | 3 (5%) |
| Native American or Native Alaskan | 1 (2%) |
| White | 49 (79%) |
| **Educational attainment** |  |
| Less than high school | 0 (0%) |
| High school diploma or equivalent | 9 (15%) |
| College or training certificate | 7 (11%) |
| Associate degree | 2 (3%) |
| Bachelor’s degree | 21 (34%) |
| Master’s degree or graduate certificate | 20 (32%) |
| Doctoral degree | 2 (3%) |
| Other | 1 (2%) |
| **Relationship to participant<sup>1</sup>** |  |
| Parent | 60 (97%) |
| Direct support provider | 9 (15%) |
| Friend | 2 (3%) |
| Guardian or conservator | 6 (10%) |
| Foster parent | 1 (2%) |
| Sibling | 0 (0%) |
| Spouse or romantic partner | 0 (0%) |
| Other | 1 (2%) |
| **How long have you known them?** |  |
| Less than 6 months | 0 (0%) |
| 6 months to a year | 0 (0%) |
| 1 year to 5 years | 0 (0%) |
| More than 5 years (but not all their life) | 2 (3%) |
| Almost or all their life | 60 (97%) |
| **Time spent with participant per week** |  |
| Less than 1 hour | 2 (3%) |
| 1 to 4 hours | 0 (0%) |
| 5 to 10 hours | 9 (15%) |
| 11 or more hours | 51 (82%) |

<sup>1</sup> Check-all-that-apply item; percentages may sum to more than 100%.
Note: Age missing for 3 participants; all other variables N = 62. Caregiver reporters completed the survey on behalf of an autistic adult who could not participate directly, even with accommodations and supports.

**Supplemental Table C1:** Survey Completion Assistance by Instrument.

| **Instrument** | **N Answered** | **Got Help? N (%)** | **Type of Help Received<sup>1</sup> (N)** |  |  |  |  |  |
|---|---|---|---|---|---|---|---|---|
|  |  |  | **Device help** | **Read survey** | **Helped Understand questions** | **Helped decide between answers** | **Other type of help** | **Answered for me** |
| **Outcome Measures** |  |  |  |  |  |  |  |  |
| ***Health and Quality of Life*** |  |  |  |  |  |  |  |  |
| AASPIRE Quality of Life Item | 787 | 81 (10%) | 38 | 50 | 55 | 19 | 2 | 4 |
| AASPIRE Overall Health Scale | 793 | 110 (14%) | 45 | 60 | 85 | 22 | 11 | 3 |
| Patient Health Questionnaire 9 (PHQ-9) – AASPIRE | 754 | 68 (9%) | 30 | 48 | 45 | 23 | 2 | 6 |
| Generalized Anxiety Scale 7 (GAD-7) – AASPIRE | 756 | 73 (10%) | 42 | 54 | 41 | 20 | 2 | 3 |
| Flourishing Scale – AASPIRE | 754 | 75 (10%) | 41 | 53 | 46 | 19 | 1 | 3 |
| AASPIRE Autistic Burnout Scale – Revised | 754 | 80 (11%) | 35 | 52 | 56 | 20 | 2 | 5 |
| ***Social Participation and Functioning*** |  |  |  |  |  |  |  |  |
| AASPIRE Community Participation Scale | 764 | 79 (10%) | 41 | 54 | 56 | 14 | 5 | 3 |
| Self Determination Inventory – AASPIRE | 757 | 82 (11%) | 40 | 51 | 58 | 24 | 4 | 6 |
| AASPIRE Choices and Decisions Scale | 763 | 78 (10%) | 41 | 47 | 54 | 19 | 1 | 6 |
| AASPIRE Barriers to Communication Scale | 754 | 81 (11%) | 41 | 51 | 61 | 26 | 3 | 4 |
| Job Satisfaction Survey – AASPIRE | 325 | 17 (5%) | 7 | 14 | 13 | 4 | 0 | 0 |
| AASPIRE Overall Employment Satisfaction Scale | 398 | 16 (4%) | 8 | 13 | 10 | 4 | 0 | 0 |
| ***Supports and Services*** |  |  |  |  |  |  |  |  |
| AASPIRE Social Support Scale | 779 | 81 (10%) | 40 | 55 | 57 | 18 | 5 | 2 |
| Satisfaction with Developmental Disability Services – AASPIRE | 246 | 60 (24%) | 25 | 34 | 35 | 19 | 4 | 4 |
| Adequacy of Supports and Information – AASPIRE | 725 | 79 (11%) | 30 | 43 | 50 | 24 | 5 | 6 |
| ***Healthcare Quality*** |  |  |  |  |  |  |  |  |
| AASPIRE Unmet Healthcare Needs Checklist – Revised | 716 | 67 (9%) | 29 | 39 | 36 | 19 | 5 | 6 |
| AASPIRE Barriers to Healthcare Checklist – Short Form – Revised | 718 | 65 (9%) | 28 | 39 | 36 | 18 | 3 | 7 |
| AASPIRE Patient Provider Communication Scale – Revised | 718 | 67 (9%) | 32 | 42 | 41 | 18 | 3 | 2 |
| AASPIRE Health and Healthcare Self-Efficacy Scale | 719 | 72 (10%) | 31 | 40 | 45 | 23 | 5 | 5 |
| **Participant Characteristics Modules** |  |  |  |  |  |  |  |  |
| AASPIRE Demographic Characteristics Module | 759 | 97 (13%) | 37 | 53 | 63 | 27 | 9 | 4 |
| AASPIRE Disability Characteristics Module | 757 | 108 (14%) | 35 | 57 | 71 | 30 | 6 | 7 |
| AASPIRE Disability Services Use Module | 730 | 90 (12%) | 29 | 42 | 56 | 21 | 8 | 9 |
| AASPIRE Communication Characteristics Module | 769 | 91 (12%) | 44 | 54 | 64 | 22 | 6 | 3 |
| AASPIRE Health Characteristics Module | 759 | 94 (12%) | 37 | 53 | 57 | 25 | 6 | 7 |
| AASPIRE Healthcare Characteristics Module | 723 | 82 (11%) | 30 | 41 | 47 | 30 | 3 | 9 |
| AASPIRE Employment Characteristics Module | 775 | 92 (12%) | 8 | 15 | 19 | 12 | 0 | 0 |
| Everyday Discrimination Scale – AASPIRE | 721 | 73 (10%) | 33 | 47 | 50 | 28 | 5 | 2 |
| Adverse Childhood Experiences Scale – AASPIRE | 725 | 65 (9%) | 31 | 43 | 37 | 15 | 4 | 3 |
| AASPIRE Alcohol and Drug Use Module | 725 | 65 (9%) | 31 | 39 | 31 | 16 | 5 | 6 |

Only includes participants using the Direct Report version of the survey. <sup>1</sup> Help types are not mutually exclusive; participants could select multiple types. N for each help type reflects number of participants who selected that option among those who reported receiving any help.
Options included:
- Someone helped me use a computer, smartphone, or other device (for example, they clicked on the answers I chose)
- Someone read the survey to me.
- Someone helped me understand what the questions or answers options meant.
- Someone helped me decide how to answer the questions on the survey.
- I got some other type of help to answer the questions.
- Someone answered the questions for me, without my input.

**Supplement C2:** Item Completion Rates by Instrument in the AASPIRE Measurement Toolkit.

| Instrument / subscale | N starters | Mean % answered | Min % | Max % | N missing >10% | % missing >10% |
|---|---|---|---|---|---|---|
| **Overall health** |  |  |  |  |  |  |
| Overall Health Self-Rated Health | 870 | 99.6 | 50.0 | 100 | 18 | 2.1% |
| Overall Health Interference | 869 | 99.7 | 62.5 | 100 | 20 | 2.3% |
| **Social support** |  |  |  |  |  |  |
| Companionship | 857 | 99.8 | 75.0 | 100 | 8 | 0.9% |
| Emotional support | 856 | 99.6 | 50.0 | 100 | 11 | 1.3% |
| Practical support | 859 | 99.8 | 60.0 | 100 | 7 | 0.8% |
| Isolation | 857 | 99.6 | 25.0 | 100 | 9 | 1.1% |
| **Employment** |  |  |  |  |  |  |
| Job satisfaction in an organization | 331 | 99.7 | 72.7 | 100 | 2 | 0.6% |
| Overall employment satisfaction | 413 | 99.7 | 60.0 | 100 | 2 | 0.5% |
| **Community participation** |  |  |  |  |  |  |
| Community Participation | 845 | 99.6 | 22.2 | 100 | 20 | 2.4% |
| **Autonomy** |  |  |  |  |  |  |
| Choices & Decisions | 839 | 99.7 | 30.8 | 100 | 6 | 0.7% |
| Self-Determination | 830 | 99.8 | 81.0 | 100 | 2 | 0.2% |
| **Mental health** |  |  |  |  |  |  |
| Flourishing | 836 | 99.8 | 75.0 | 100 | 11 | 1.3% |
| Anxiety (GAD) | 836 | 99.8 | 85.7 | 100 | 10 | 1.2% |
| Depression (PHQ-9) | 836 | 99.8 | 77.8 | 100 | 13 | 1.6% |
| Autistic Burnout | 835 | 99.7 | 78.6 | 100 | 2 | 0.2% |
| **Disability services** |  |  |  |  |  |  |
| Satisfaction with Disability Services | 302 | 99.2 | 50.0 | 100 | 3 | 1.0% |
| Adequacy of Supports and Information | 798 | 99.6 | 55.6 | 100 | 22 | 2.8% |
| **Healthcare** |  |  |  |  |  |  |
| Patient-Provider Communication | 793 | 99.8 | 75.0 | 100 | 13 | 1.6% |
| Health Self-Efficacy – Individual | 792 | 99.7 | 70.0 | 100 | 2 | 0.3% |
| Health Self-Efficacy – Relationship Dependent | 792 | 99.4 | 54.5 | 100 | 3 | 0.4% |

Among participants who started each instrument. Mean %, min %, and max % reflect the percentage of items answered. The final two columns show the number and percentage of starters who missed more than 10% of items.

**Supplement C3:** Follow-Up Survey Completion by Subgroup.

| **Time Point** | **Total N=870** | **Subcohort** |  |  | **Support Needs** |  |  | **Reporter Type** |  |  |
|---|---|---|---|---|---|---|---|---|---|---|
|  |  | **Disability Services N=266** | **Healthcare N=319** | **Community N=285** | **No Assistance N=243** | **IADLs Only N=371** | **ADLs N=221** | **Direct Report (DR) N=605** | **DR – Supported (DR-S) N=194** | **Caregiver Report (CR) N=71** |
| Time 2 (n, % of baseline) | 751 (86%) | 224 (85%) | 270 (85%) | 257 (90%) | 228 (94%) | 308 (83%) | 184 (83%) | 576 (95%) | 113 (58%) | 62 (87%) |
| Time 3 (n, % of baseline) | 705 (81%) | 208 (78%) | 245 (77%) | 252 (88%) | 237 (98%) | 276 (74%) | 178 (81%) | 566 (94%) | 88 (45%) | 51 (72%) |

No Assistance = does not need assistance with ADLs or IADLs; IADLs Only = needs assistance with instrumental activities only; ADLs = needs assistance with activities of daily living (may also need assistance with IADLs).
Percentages reflect participants who completed the Time 2 and Time 3 surveys as a proportion of the N who started the baseline survey in that subgroup.

**Supplemental Table C4:** Perceived Survey Ease by Subgroup.

| **Survey Portion** | **Total N<sup>1</sup>** | **Subcohort** |  |  | **Support Needs** |  |  | **Reporter Type** |  |  |
|---|---|---|---|---|---|---|---|---|---|---|
|  |  | **Disability Services (DDS)** | **Healthcare** | **Community** | **No Assistance** | **IADLs Only** | **ADLs** | **Direct Report (DR)** | **DR – Supported (DR-S)** | **Caregiver Report (CR)** |
| Found first half of survey easy or very easy | 783 (94%) | 230 (89%) | 282 (95%) | 271 (97%) | 237 (98%) | 349 (94%) | 197 (90%) | 565 (97%) | 154 (85%) | 64 (96%) |
| Found second half of survey easy or very easy | 740 (94%) | 222 (91%) | 260 (94%) | 258 (97%) | 224 (97%) | 329 (94%) | 187 (90%) | 537 (96%) | 141 (84%) | 62 (98%) |

N = number who responded to this item (excludes missing). Easy or very easy = combined "Very Easy" and "Easy" responses, compared to "Neutral," "Hard," and "Very Hard." Percentages are column percentages within each subgroup. <sup>1</sup> Total N varies between 1<sup>st</sup> half (N=833) and 2<sup>nd</sup> half (N=788) due to attrition and missing data.

**Supplemental Table D1:**
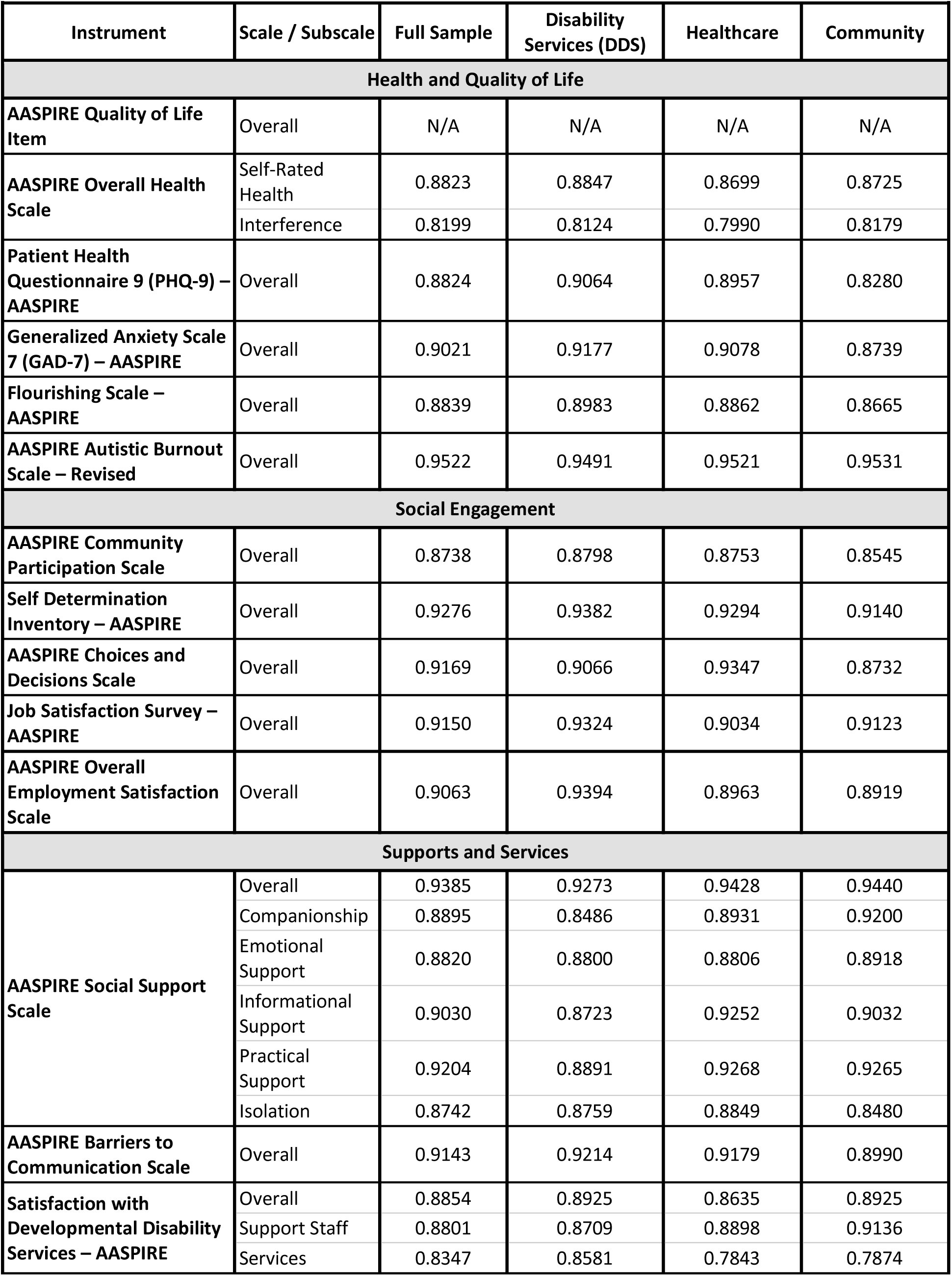
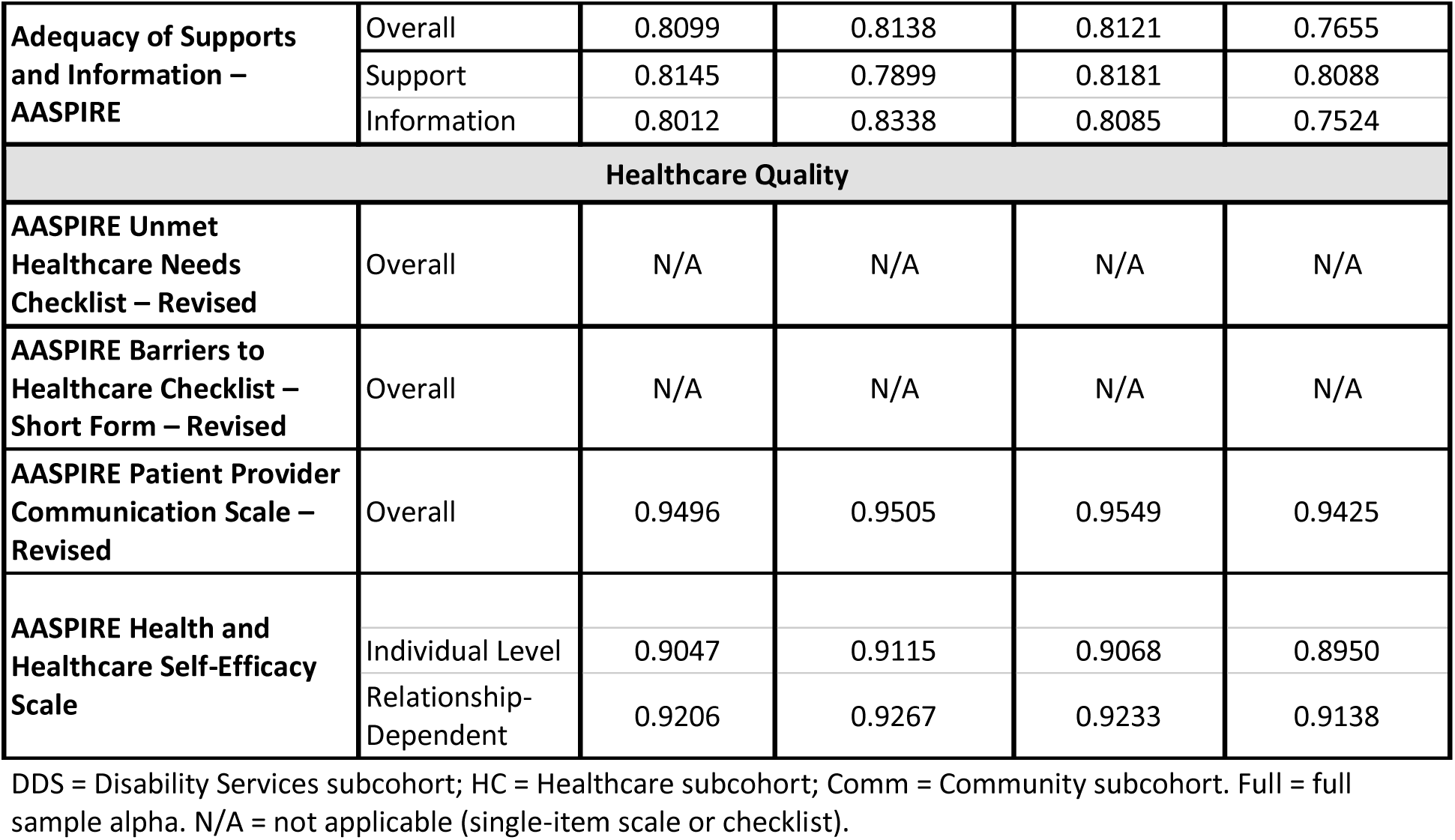
Internal Consistency (Cronbach’s α) by Subcohort.

**Supplemental Table D2:** Internal Consistency (Cronbach’s α) by Support Needs.

| **Instrument** | **Scale / Subscale** | **Full Sample** | **No Assistance Needed for ADL’s or IADLs** | **Assistance with IADLs Only** | **Assistance with Basic ADLs** |
|---|---|---|---|---|---|
| **Health and Quality of Life** |  |  |  |  |  |
| **AASPIRE Quality of Life Item** | Overall | N/A | N/A | N/A | N/A |
| **AASPIRE Overall Health Scale** | Self-Rated Health | 0.8823 | 0.8824 | 0.8468 | 0.9121 |
|  | Interference | 0.8199 | 0.8050 | 0.7778 | 0.8559 |
| **Patient Health Questionnaire 9 (PHQ-9) – AASPIRE** | Overall | 0.8824 | 0.8713 | 0.8675 | 0.9052 |
| **Generalized Anxiety Scale 7 (GAD-7) – AASPIRE** | Overall | 0.9021 | 0.9122 | 0.8888 | 0.9061 |
| **Flourishing Scale – AASPIRE** | Overall | 0.8839 | 0.8716 | 0.8809 | 0.8959 |
| **AASPIRE Autistic Burnout Scale – Revised** | Overall | 0.9522 | 0.9457 | 0.9434 | 0.9637 |
| **Social Engagement** |  |  |  |  |  |
| **AASPIRE Community Participation Scale** | Overall | 0.8738 | 0.8773 | 0.8541 | 0.8924 |
| **Self Determination Inventory – AASPIRE** | Overall | 0.9276 | 0.9184 | 0.9116 | 0.9318 |
| **AASPIRE Choices and Decisions Scale** | Overall | 0.9169 | 0.9250 | 0.8745 | 0.9241 |
| **AASPIRE Barriers to Communication Scale** | Overall | 0.9143 | 0.9082 | 0.8967 | 0.9138 |
| **Job Satisfaction Survey – AASPIRE** | Overall | 0.9150 | 0.9253 | 0.9057 | 0.9191 |
| **AASPIRE Overall Employment Satisfaction Scale** | Overall | 0.9063 | 0.9185 | 0.8992 | 0.8973 |
| **Supports and Services** |  |  |  |  |  |
| **AASPIRE Social Support Scale** | Overall | 0.9385 | 0.9436 | 0.9351 | 0.9397 |
|  | Companionship | 0.8895 | 0.9066 | 0.8760 | 0.8938 |
|  | Emotional Support | 0.8820 | 0.8829 | 0.8857 | 0.8782 |
|  | Informational Support | 0.9030 | 0.9153 | 0.8929 | 0.9088 |
|  | Practical Support | 0.9204 | 0.9204 | 0.8990 | 0.9318 |
|  | Isolation | 0.8742 | 0.8777 | 0.8517 | 0.9016 |
| **Satisfaction with Developmental Disability Services – AASPIRE** | Overall | 0.8854 | 0.8513 | 0.8838 | 0.9010 |
|  | Support Staff | 0.8801 | 0.8083 | 0.8675 | 0.9136 |
|  | Services | 0.8347 | 0.8534 | 0.8060 | 0.8579 |
| **Adequacy of Supports and Information – AASPIRE** | Overall | 0.8099 | 0.7700 | 0.7911 | 0.8900 |
|  | Support | 0.8145 | 0.7998 | 0.7487 | 0.8815 |
|  | Information | 0.8012 | 0.8039 | 0.7874 | 0.8354 |
| **Healthcare Quality** |  |  |  |  |  |
| **AASPIRE Unmet Healthcare Needs Checklist – Revised** | Overall | N/A | N/A | N/A | N/A |
| **AASPIRE Barriers to Healthcare Checklist – Short Form – Revised** | Overall | N/A | N/A | N/A | N/A |
| **AASPIRE Patient Provider Communication Scale – Revised** | Overall | 0.9496 | 0.9515 | 0.9457 | 0.9531 |
| **AASPIRE Health and Healthcare Self-Efficacy Scale** | Individual Level | 0.9047 | 0.8852 | 0.8890 | 0.9101 |
|  | Relationship-Dependent | 0.9206 | 0.9091 | 0.9078 | 0.9306 |

**Supplemental Table D3:** Internal Consistency (Cronbach’s α) by Reporter Type.

| **Instrument** | **Scale / Subscale** | **Direct Report (DR)** |  | **DR – Supported (DR-S)** |  | **Caregiver Report (CR)** |  |
|---|---|---|---|---|---|---|---|
|  |  | **N** | **α** | **N** | **α** | **N** | **α** |
| **Health and Quality of Life** |  |  |  |  |  |  |  |
| **AASPIRE Quality of Life Item** | Overall | 604 | N/A | 191 | N/A | 70 | N/A |
| **AASPIRE Overall Health Scale** | Self-Rated Health | 605 | 0.8810 | 194 | 0.8664 | 71 | 0.8785 |
|  | Interference |  | 0.8319 |  | 0.7704 |  | 0.7012 |
| **Patient Health Questionnaire 9 (PHQ-9) – AASPIRE** | Overall | 586 | 0.8731 | 182 | 0.8802 | 68 | 0.8963 |
| **Generalized Anxiety Scale 7 (GAD-7) – AASPIRE** | Overall | 586 | 0.8959 | 182 | 0.9041 | 68 | 0.9272 |
| **Flourishing Scale – AASPIRE** | Overall | 586 | 0.8797 | 182 | 0.8921 | 68 | 0.8655 |
| **AASPIRE Autistic Burnout Scale – Revised** | Overall | 585 | 0.9502 | 182 | 0.9434 | 68 | 0.9710 |
| **Social Engagement** |  |  |  |  |  |  |  |
| **AASPIRE Community Participation Scale** | Overall | 590 | 0.8737 | 187 | 0.8570 | 69 | 0.8898 |
| **Self Determination Inventory – AASPIRE** | Overall | 586 | 0.9214 | 183 | 0.9221 | 69 | 0.9151 |
| **AASPIRE Choices and Decisions Scale** | Overall | 587 | 0.8983 | 183 | 0.8710 | 69 | 0.8872 |
| **Job Satisfaction Survey – AASPIRE** | Overall | 278 | 0.9178 | 50 | 0.9507 | 3 | 1.0000<sup>a</sup> |
| **AASPIRE Overall Employment Satisfaction Scale** | Overall | 346 | 0.9013 | 59 | 0.9225 | 7 | 0.9070<sup>a</sup> |
| **Supports and Services** |  |  |  |  |  |  |  |
| **AASPIRE Social Support Scale** | Overall | 598 | 0.9178 | 187 | 0.9507 | 70 | 0.9324 |
|  | Companionship |  | 0.9019 |  | 0.8210 |  | 0.8780 |
|  | Emotional Support |  | 0.8893 |  | 0.8642 |  | 0.8296 |
|  | Informational Support |  | 0.9069 |  | 0.8574 |  | 0.9434 |
|  | Practical Support |  | 0.9120 |  | 0.8670 |  | 0.9325 |
|  | Isolation |  | 0.8617 |  | 0.8638 |  | 0.9400 |
| **AASPIRE Barriers to Communication Scale** | Overall | 588 | 0.9150 | 183 | 0.9302 | 70 | 0.8728 |
| **Satisfaction with Developmental Disability Services – AASPIRE** | Overall | 141 | 0.8784 | 108 | 0.9047 | 54 | 0.8782 |
|  | Support Staff |  | 0.8809 |  | 0.8836 |  | 0.8873 |
|  | Services |  | 0.8259 |  | 0.8573 |  | 0.8309 |
| **Adequacy of Supports and Information – AASPIRE** | Overall | 563 | 0.8547 | 170 | 0.8685 | 65 | 0.8396 |
|  | Support |  | 0.8627 |  | 0.8639 |  | 0.8548 |
|  | Information |  | 0.8060 |  | 0.8315 |  | 0.8478 |
| **Healthcare Quality** |  |  |  |  |  |  |  |
| **AASPIRE Unmet Healthcare Needs Checklist – Revised** | Overall | 560 | N/A | 169 | N/A | 65 | N/A |
| **AASPIRE Barriers to Healthcare Checklist – Short Form – Revised** | Overall | 560 | N/A | 169 | N/A | 65 | N/A |
| **AASPIRE Patient Provider Communication Scale – Revised** | Overall | 560 | 0.9499 | 169 | 0.9459 | 65 | 0.9542 |
| **AASPIRE Health and Healthcare Self-Efficacy Scale** |  | 560 |  | 169 |  | 65 |  |
|  | Individual Level |  | 0.9106 |  | 0.9084 |  | 0.8646 |
|  | Relationship-Dependent |  | 0.9238 |  | 0.9185 |  | 0.9192 |

DR = Direct Report; DR-S = Direct Report with Support; CR = Caregiver Report. N = number of participants with complete data. N/A = not applicable (single-item scales or checklists). <sup>a</sup> alphas for Job Satisfaction Survey and Overall Employment Satisfaction in the CR sample were based on N's of 3 and 7 respectively so interpret with caution.

**Supplement E1:**
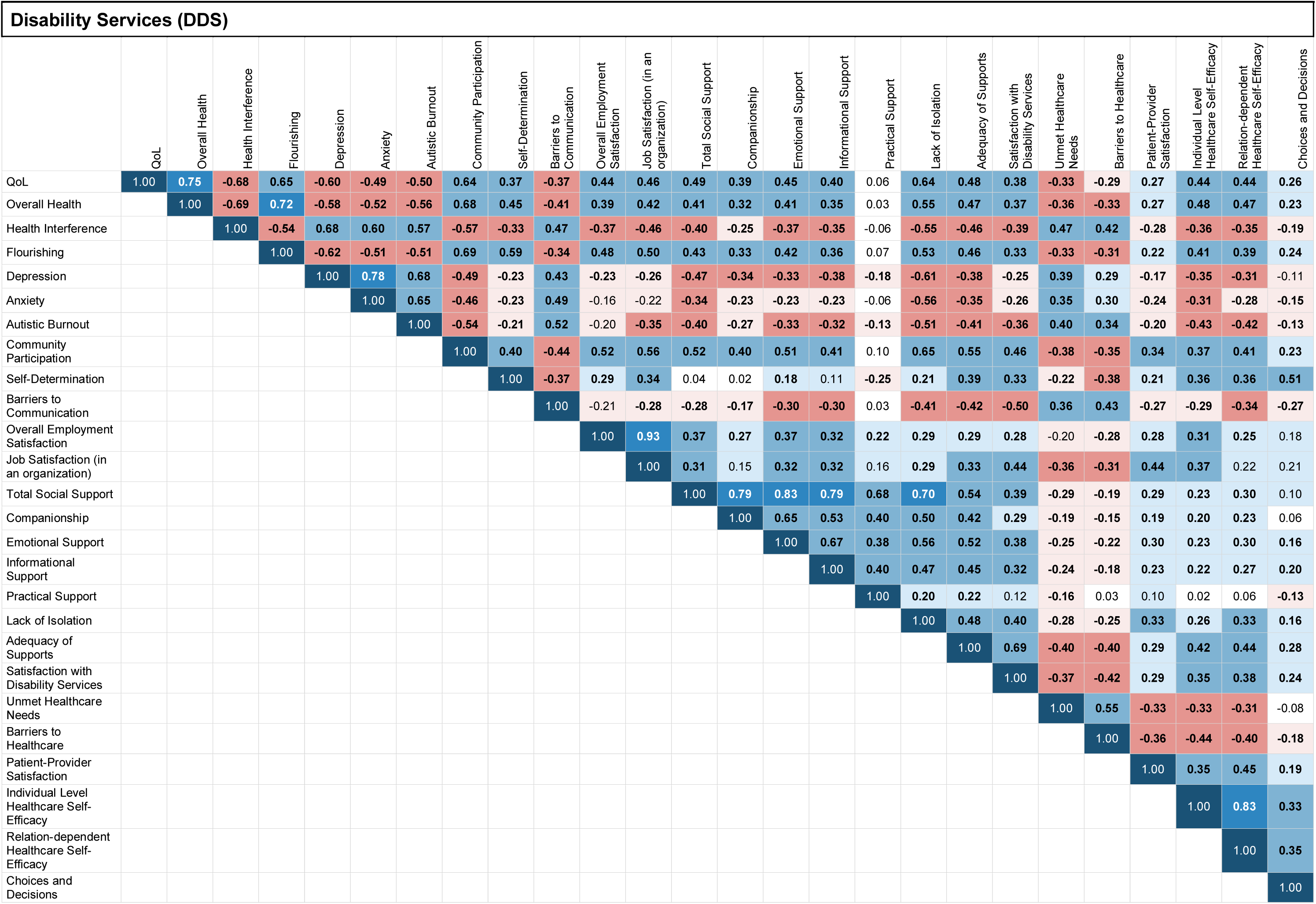
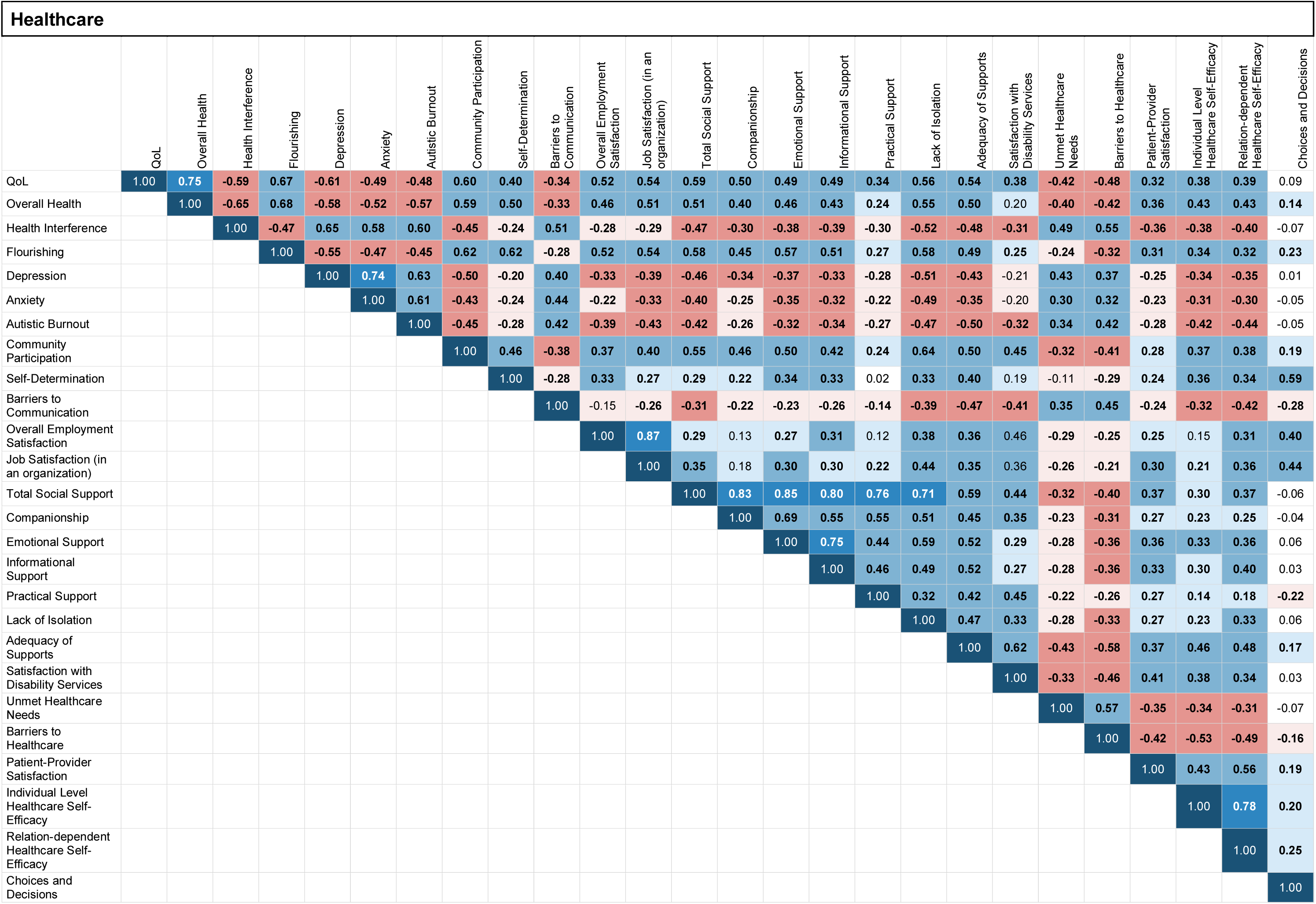
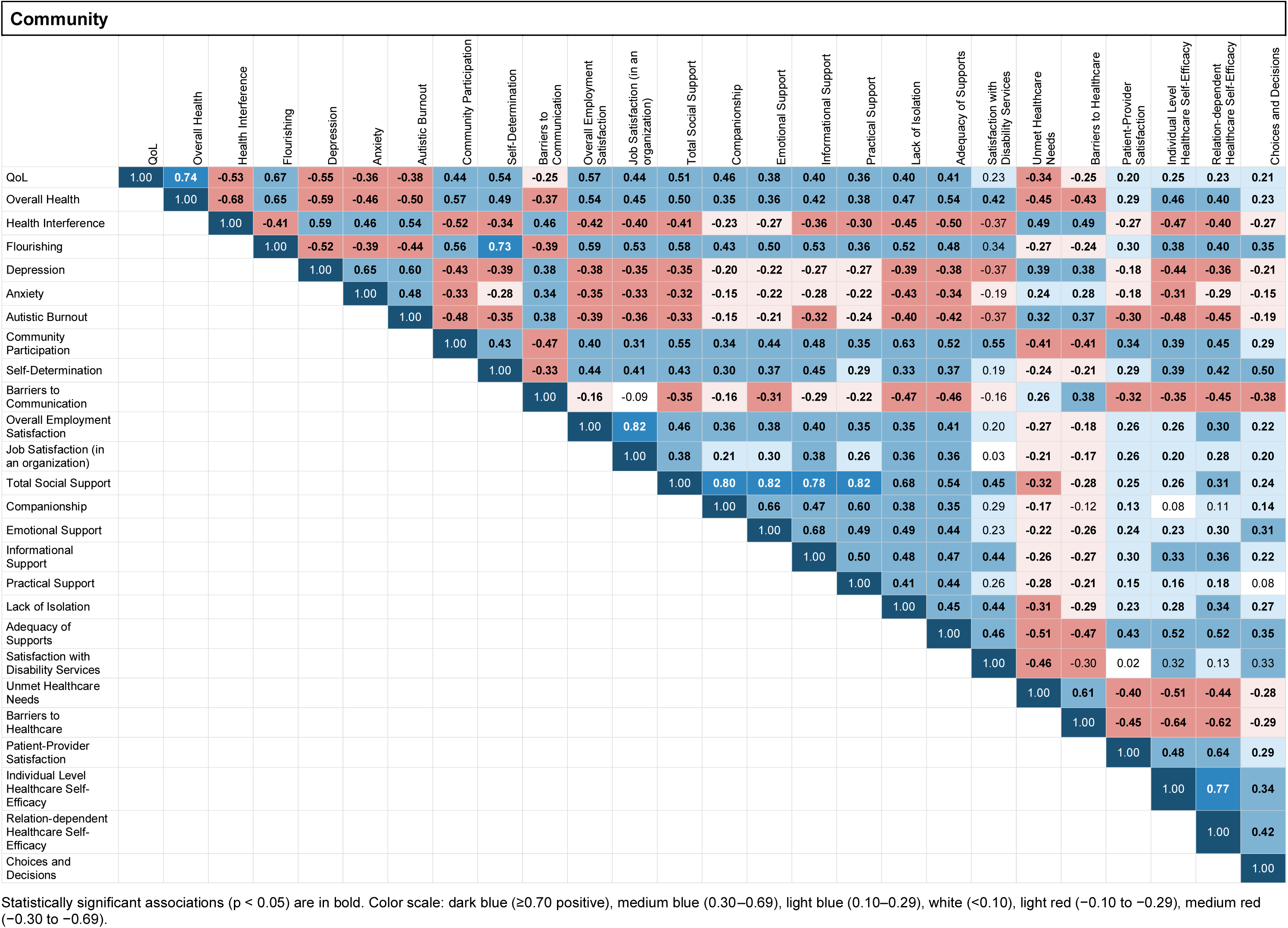
Correlations Between Outcomes, Divided by Subcohorts.

| **Disability Services (DDS)** |  |  |  |  |  |  |  |  |  |  |  |  |  |  |  |  |  |  |  |  |  |  |  |  |  |  |
|  | QoL | Overall Health | Health Interference | Flourishing | Depression | Anxiety | Autistic Burnout | Community Participation | Self-Determination | Barriers to Communication | Overall Employment Satisfaction | Job Satisfaction (in an organization) | Total Social Support | Companionship | Emotional Support | Informational Support | Practical Support | Lack of Isolation | Adequacy of Supports | Satisfaction with Disability Services | Unmet Healthcare Needs | Barriers to Healthcare | Patient-Provider Satisfaction | Individual Level Healthcare Self-Efficacy | Relation-dependent Healthcare Self-Efficacy | Choices and Decisions |
|---|---|---|---|---|---|---|---|---|---|---|---|---|---|---|---|---|---|---|---|---|---|---|---|---|---|---|
| QoL | 1.00 | **0.75** | **-0.68** | **0.65** | **-0.60** | **-0.49** | **-0.50** | **0.64** | **0.37** | **-0.37** | **0.44** | **0.46** | **0.49** | **0.39** | **0.45** | **0.40** | 0.06 | **0.64** | **0.48** | **0.38** | **-0.33** | **-0.29** | **0.27** | **0.44** | **0.44** | **0.26** |
| Overall Health |  | 1.00 | **-0.69** | **0.72** | **-0.58** | **-0.52** | **-0.56** | **0.68** | **0.45** | **-0.41** | **0.39** | **0.42** | **0.41** | **0.32** | **0.41** | **0.35** | 0.03 | **0.55** | **0.47** | **0.37** | **-0.36** | **-0.33** | **0.27** | **0.48** | **0.47** | **0.23** |
| Health Interference |  |  | 1.00 | **-0.54** | **0.68** | **0.60** | **0.57** | **-0.57** | **-0.33** | **0.47** | **-0.37** | **-0.46** | **-0.40** | **-0.25** | **-0.37** | **-0.35** | -0.06 | **-0.55** | **-0.46** | **-0.39** | **0.47** | **0.42** | **-0.28** | **-0.36** | **-0.35** | **-0.19** |
| Flourishing |  |  |  | 1.00 | **-0.62** | **-0.51** | **-0.51** | **0.69** | **0.59** | **-0.34** | **0.48** | **0.50** | **0.43** | **0.33** | **0.42** | **0.36** | 0.07 | **0.53** | **0.46** | **0.33** | **-0.33** | **-0.31** | **0.22** | **0.41** | **0.39** | **0.24** |
| Depression |  |  |  |  | 1.00 | **0.78** | **0.68** | **-0.49** | **-0.23** | **0.43** | **-0.23** | **-0.26** | **-0.47** | **-0.34** | **-0.33** | **-0.38** | **-0.18** | **-0.61** | **-0.38** | **-0.25** | **0.39** | **0.29** | **-0.17** | **-0.35** | **-0.31** | -0.11 |
| Anxiety |  |  |  |  |  | 1.00 | **0.65** | **-0.46** | **-0.23** | **0.49** | -0.16 | -0.22 | **-0.34** | **-0.23** | **-0.23** | **-0.23** | -0.06 | **-0.56** | **-0.35** | **-0.26** | **0.35** | **0.30** | **-0.24** | **-0.31** | **-0.28** | **-0.15** |
| Autistic Burnout |  |  |  |  |  |  | 1.00 | **-0.54** | **-0.21** | **0.52** | -0.20 | **-0.35** | **-0.40** | **-0.27** | **-0.33** | **-0.32** | **-0.13** | **-0.51** | **-0.41** | **-0.36** | **0.40** | **0.34** | **-0.20** | **-0.43** | **-0.42** | **-0.13** |
| Community Participation |  |  |  |  |  |  |  | 1.00 | **0.40** | **-0.44** | **0.52** | **0.56** | **0.52** | **0.40** | **0.51** | **0.41** | 0.10 | **0.65** | **0.55** | **0.46** | **-0.38** | **-0.35** | **0.34** | **0.37** | **0.41** | **0.23** |
| Self-Determination |  |  |  |  |  |  |  |  | 1.00 | **-0.37** | **0.29** | **0.34** | 0.04 | 0.02 | **0.18** | 0.11 | **-0.25** | **0.21** | **0.39** | **0.33** | **-0.22** | **-0.38** | **0.21** | **0.36** | **0.36** | **0.51** |
| Barriers to Communication |  |  |  |  |  |  |  |  |  | 1.00 | -0.21 | **-0.28** | **-0.28** | **-0.17** | **-0.30** | **-0.30** | 0.03 | **-0.41** | **-0.42** | **-0.50** | **0.36** | **0.43** | **-0.27** | **-0.29** | **-0.34** | **-0.27** |
| Overall Employment Satisfaction |  |  |  |  |  |  |  |  |  |  | 1.00 | **0.93** | **0.37** | **0.27** | **0.37** | **0.32** | **0.22** | **0.29** | **0.29** | **0.28** | -0.20 | **-0.28** | **0.28** | **0.31** | **0.25** | 0.18 |
| Job Satisfaction (in an organization) |  |  |  |  |  |  |  |  |  |  |  | 1.00 | **0.31** | 0.15 | **0.32** | **0.32** | 0.16 | **0.29** | **0.33** | **0.44** | **-0.36** | **-0.31** | **0.44** | **0.37** | 0.22 | 0.21 |
| Total Social Support |  |  |  |  |  |  |  |  |  |  |  |  | 1.00 | **0.79** | **0.83** | **0.79** | **0.68** | **0.70** | **0.54** | **0.39** | **-0.29** | **-0.19** | **0.29** | **0.23** | **0.30** | 0.10 |
| Companionship |  |  |  |  |  |  |  |  |  |  |  |  |  | 1.00 | **0.65** | **0.53** | **0.40** | **0.50** | **0.42** | **0.29** | **-0.19** | **-0.15** | **0.19** | **0.20** | **0.23** | 0.06 |
| Emotional Support |  |  |  |  |  |  |  |  |  |  |  |  |  |  | 1.00 | **0.67** | **0.38** | **0.56** | **0.52** | **0.38** | **-0.25** | **-0.22** | **0.30** | **0.23** | **0.30** | **0.16** |
| Informational Support |  |  |  |  |  |  |  |  |  |  |  |  |  |  |  | 1.00 | **0.40** | **0.47** | **0.45** | **0.32** | **-0.24** | **-0.18** | **0.23** | **0.22** | **0.27** | **0.20** |
| Practical Support |  |  |  |  |  |  |  |  |  |  |  |  |  |  |  |  | 1.00 | **0.20** | **0.22** | 0.12 | **-0.16** | 0.03 | 0.10 | 0.02 | 0.06 | **-0.13** |
| Lack of Isolation |  |  |  |  |  |  |  |  |  |  |  |  |  |  |  |  |  | 1.00 | **0.48** | **0.40** | **-0.28** | **-0.25** | **0.33** | **0.26** | **0.33** | **0.16** |
| Adequacy of Supports |  |  |  |  |  |  |  |  |  |  |  |  |  |  |  |  |  |  | 1.00 | **0.69** | **-0.40** | **-0.40** | **0.29** | **0.42** | **0.44** | **0.28** |
| Satisfaction with Disability Services |  |  |  |  |  |  |  |  |  |  |  |  |  |  |  |  |  |  |  | 1.00 | **-0.37** | **-0.42** | **0.29** | **0.35** | **0.38** | **0.24** |
| Unmet Healthcare Needs |  |  |  |  |  |  |  |  |  |  |  |  |  |  |  |  |  |  |  |  | 1.00 | **0.55** | **-0.33** | **-0.33** | **-0.31** | -0.08 |
| Barriers to Healthcare |  |  |  |  |  |  |  |  |  |  |  |  |  |  |  |  |  |  |  |  |  | 1.00 | **-0.36** | **-0.44** | **-0.40** | **-0.18** |
| Patient-Provider Satisfaction |  |  |  |  |  |  |  |  |  |  |  |  |  |  |  |  |  |  |  |  |  |  | 1.00 | **0.35** | **0.45** | **0.19** |
| Individual Level Healthcare Self-Efficacy |  |  |  |  |  |  |  |  |  |  |  |  |  |  |  |  |  |  |  |  |  |  |  | 1.00 | **0.83** | **0.33** |
| Relation-dependent Healthcare Self-Efficacy |  |  |  |  |  |  |  |  |  |  |  |  |  |  |  |  |  |  |  |  |  |  |  |  | 1.00 | **0.35** |
| Choices and Decisions |  |  |  |  |  |  |  |  |  |  |  |  |  |  |  |  |  |  |  |  |  |  |  |  |  | 1.00 |
| **Healthcare** |  |  |  |  |  |  |  |  |  |  |  |  |  |  |  |  |  |  |  |  |  |  |  |  |  |  |
|  | QoL | Overall Health | Health Interference | Flourishing | Depression | Anxiety | Autistic Burnout | Community Participation | Self-Determination | Barriers to Communication | Overall Employment Satisfaction | Job Satisfaction (in an organization) | Total Social Support | Companionship | Emotional Support | Informational Support | Practical Support | Lack of Isolation | Adequacy of Supports | Satisfaction with Disability Services | Unmet Healthcare Needs | Barriers to Healthcare | Patient-Provider Satisfaction | Individual Level Healthcare Self-Efficacy | Relation-dependent Healthcare Self-Efficacy | Choices and Decisions |
| QoL | 1.00 | **0.75** | **-0.59** | **0.67** | **-0.61** | **-0.49** | **-0.48** | **0.60** | **0.40** | **-0.34** | **0.52** | **0.54** | **0.59** | **0.50** | **0.49** | **0.49** | **0.34** | **0.56** | **0.54** | **0.38** | **-0.42** | **-0.48** | **0.32** | **0.38** | **0.39** | 0.09 |
| Overall Health |  | 1.00 | **-0.65** | **0.68** | **-0.58** | **-0.52** | **-0.57** | **0.59** | **0.50** | **-0.33** | **0.46** | **0.51** | **0.51** | **0.40** | **0.46** | **0.43** | **0.24** | **0.55** | **0.50** | 0.20 | **-0.40** | **-0.42** | **0.36** | **0.43** | **0.43** | **0.14** |
| Health Interference |  |  | 1.00 | **-0.47** | **0.65** | **0.58** | **0.60** | **-0.45** | **-0.24** | **0.51** | **-0.28** | **-0.29** | **-0.47** | **-0.30** | **-0.38** | **-0.39** | **-0.30** | **-0.52** | **-0.48** | **-0.31** | **0.49** | **0.55** | **-0.36** | **-0.38** | **-0.40** | -0.07 |
| Flourishing |  |  |  | 1.00 | **-0.55** | **-0.47** | **-0.45** | **0.62** | **0.62** | **-0.28** | **0.52** | **0.54** | **0.58** | **0.45** | **0.57** | **0.51** | **0.27** | **0.58** | **0.49** | **0.25** | **-0.24** | **-0.32** | **0.31** | **0.34** | **0.32** | **0.23** |
| Depression |  |  |  |  | 1.00 | **0.74** | **0.63** | **-0.50** | **-0.20** | **0.40** | **-0.33** | **-0.39** | **-0.46** | **-0.34** | **-0.37** | **-0.33** | **-0.28** | **-0.51** | **-0.43** | -0.21 | **0.43** | **0.37** | **-0.25** | **-0.34** | **-0.35** | 0.01 |
| Anxiety |  |  |  |  |  | 1.00 | **0.61** | **-0.43** | **-0.24** | **0.44** | **-0.22** | **-0.33** | **-0.40** | **-0.25** | **-0.35** | **-0.32** | **-0.22** | **-0.49** | **-0.35** | -0.20 | **0.30** | **0.32** | **-0.23** | **-0.31** | **-0.30** | -0.05 |
| Autistic Burnout |  |  |  |  |  |  | 1.00 | **-0.45** | **-0.28** | **0.42** | **-0.39** | **-0.43** | **-0.42** | **-0.26** | **-0.32** | **-0.34** | **-0.27** | **-0.47** | **-0.50** | **-0.32** | **0.34** | **0.42** | **-0.28** | **-0.42** | **-0.44** | -0.05 |
| Community Participation |  |  |  |  |  |  |  | 1.00 | **0.46** | **-0.38** | **0.37** | **0.40** | **0.55** | **0.46** | **0.50** | **0.42** | **0.24** | **0.64** | **0.50** | **0.45** | **-0.32** | **-0.41** | **0.28** | **0.37** | **0.38** | **0.19** |
| Self-Determination |  |  |  |  |  |  |  |  | 1.00 | **-0.28** | **0.33** | **0.27** | **0.29** | **0.22** | **0.34** | **0.33** | 0.02 | **0.33** | **0.40** | 0.19 | -0.11 | **-0.29** | **0.24** | **0.36** | **0.34** | **0.59** |
| Barriers to Communication |  |  |  |  |  |  |  |  |  | 1.00 | -0.15 | **-0.26** | **-0.31** | **-0.22** | **-0.23** | **-0.26** | **-0.14** | **-0.39** | **-0.47** | **-0.41** | **0.35** | **0.45** | **-0.24** | **-0.32** | **-0.42** | **-0.28** |
| Overall Employment Satisfaction |  |  |  |  |  |  |  |  |  |  | 1.00 | **0.87** | **0.29** | 0.13 | **0.27** | **0.31** | 0.12 | **0.38** | **0.36** | 0.46 | **-0.29** | **-0.25** | **0.25** | 0.15 | **0.31** | **0.40** |
| Job Satisfaction (in an organization) |  |  |  |  |  |  |  |  |  |  |  | 1.00 | **0.35** | 0.18 | **0.30** | **0.30** | **0.22** | **0.44** | **0.35** | 0.36 | **-0.26** | **-0.21** | **0.30** | **0.21** | **0.36** | **0.44** |
| Total Social Support |  |  |  |  |  |  |  |  |  |  |  |  | 1.00 | **0.83** | **0.85** | **0.80** | **0.76** | **0.71** | **0.59** | **0.44** | **-0.32** | **-0.40** | **0.37** | **0.30** | **0.37** | -0.06 |
| Companionship |  |  |  |  |  |  |  |  |  |  |  |  |  | 1.00 | **0.69** | **0.55** | **0.55** | **0.51** | **0.45** | **0.35** | **-0.23** | **-0.31** | **0.27** | **0.23** | **0.25** | -0.04 |
| Emotional Support |  |  |  |  |  |  |  |  |  |  |  |  |  |  | 1.00 | **0.75** | **0.44** | **0.59** | **0.52** | **0.29** | **-0.28** | **-0.36** | **0.36** | **0.33** | **0.36** | 0.06 |
| Informational Support |  |  |  |  |  |  |  |  |  |  |  |  |  |  |  | 1.00 | **0.46** | **0.49** | **0.52** | **0.27** | **-0.28** | **-0.36** | **0.33** | **0.30** | **0.40** | 0.03 |
| Practical Support |  |  |  |  |  |  |  |  |  |  |  |  |  |  |  |  | 1.00 | **0.32** | **0.42** | **0.45** | **-0.22** | **-0.26** | **0.27** | **0.14** | **0.18** | **-0.22** |
| Lack of Isolation |  |  |  |  |  |  |  |  |  |  |  |  |  |  |  |  |  | 1.00 | **0.47** | **0.33** | **-0.28** | **-0.33** | **0.27** | **0.23** | **0.33** | 0.06 |
| Adequacy of Supports |  |  |  |  |  |  |  |  |  |  |  |  |  |  |  |  |  |  | 1.00 | **0.62** | **-0.43** | **-0.58** | **0.37** | **0.46** | **0.48** | **0.17** |
| Satisfaction with Disability Services |  |  |  |  |  |  |  |  |  |  |  |  |  |  |  |  |  |  |  | 1.00 | **-0.33** | **-0.46** | **0.41** | **0.38** | **0.34** | 0.03 |
| Unmet Healthcare Needs |  |  |  |  |  |  |  |  |  |  |  |  |  |  |  |  |  |  |  |  | 1.00 | **0.57** | **-0.35** | **-0.34** | **-0.31** | -0.07 |
| Barriers to Healthcare |  |  |  |  |  |  |  |  |  |  |  |  |  |  |  |  |  |  |  |  |  | 1.00 | **-0.42** | **-0.53** | **-0.49** | **-0.16** |
| Patient-Provider Satisfaction |  |  |  |  |  |  |  |  |  |  |  |  |  |  |  |  |  |  |  |  |  |  | 1.00 | **0.43** | **0.56** | **0.19** |
| Individual Level Healthcare Self-Efficacy |  |  |  |  |  |  |  |  |  |  |  |  |  |  |  |  |  |  |  |  |  |  |  | 1.00 | **0.78** | **0.20** |
| Relation-dependent Healthcare Self-Efficacy |  |  |  |  |  |  |  |  |  |  |  |  |  |  |  |  |  |  |  |  |  |  |  |  | 1.00 | **0.25** |
| Choices and Decisions |  |  |  |  |  |  |  |  |  |  |  |  |  |  |  |  |  |  |  |  |  |  |  |  |  | 1.00 |
| **Community** |  |  |  |  |  |  |  |  |  |  |  |  |  |  |  |  |  |  |  |  |  |  |  |  |  |  |
|  | QoL | Overall Health | Health Interference | Flourishing | Depression | Anxiety | Autistic Burnout | Community Participation | Self-Determination | Barriers to Communication | Overall Employment Satisfaction | Job Satisfaction (in an organization) | Total Social Support | Companionship | Emotional Support | Informational Support | Practical Support | Lack of Isolation | Adequacy of Supports | Satisfaction with Disability Services | Unmet Healthcare Needs | Barriers to Healthcare | Patient-Provider Satisfaction | Individual Level Healthcare Self-Efficacy | Relation-dependent Healthcare Self-Efficacy | Choices and Decisions |
| QoL | 1.00 | **0.74** | **-0.53** | **0.67** | **-0.55** | **-0.36** | **-0.38** | **0.44** | **0.54** | **-0.25** | **0.57** | **0.44** | **0.51** | **0.46** | **0.38** | **0.40** | **0.36** | **0.40** | **0.41** | 0.23 | **-0.34** | **-0.25** | **0.20** | **0.25** | **0.23** | **0.21** |
| Overall Health |  | 1.00 | **-0.68** | **0.65** | **-0.59** | **-0.46** | **-0.50** | **0.57** | **0.49** | **-0.37** | **0.54** | **0.45** | **0.50** | **0.35** | **0.36** | **0.42** | **0.38** | **0.47** | **0.54** | **0.42** | **-0.45** | **-0.43** | **0.29** | **0.46** | **0.40** | **0.23** |
| Health Interference |  |  | 1.00 | **-0.41** | **0.59** | **0.46** | **0.54** | **-0.52** | **-0.34** | **0.46** | **-0.42** | **-0.40** | **-0.41** | **-0.23** | **-0.27** | **-0.36** | **-0.30** | **-0.45** | **-0.50** | -0.37 | **0.49** | **0.49** | **-0.27** | **-0.47** | **-0.40** | **-0.27** |
| Flourishing |  |  |  | 1.00 | **-0.52** | **-0.39** | **-0.44** | **0.56** | **0.73** | **-0.39** | **0.59** | **0.53** | **0.58** | **0.43** | **0.50** | **0.53** | **0.36** | **0.52** | **0.48** | 0.34 | **-0.27** | **-0.24** | **0.30** | **0.38** | **0.40** | **0.35** |
| Depression |  |  |  |  | 1.00 | **0.65** | **0.60** | **-0.43** | **-0.39** | **0.38** | **-0.38** | **-0.35** | **-0.35** | **-0.20** | **-0.22** | **-0.27** | **-0.27** | **-0.39** | **-0.38** | -0.37 | **0.39** | **0.38** | **-0.18** | **-0.44** | **-0.36** | **-0.21** |
| Anxiety |  |  |  |  |  | 1.00 | **0.48** | **-0.33** | **-0.28** | **0.34** | **-0.35** | **-0.33** | **-0.32** | **-0.15** | **-0.22** | **-0.28** | **-0.22** | **-0.43** | **-0.34** | -0.19 | **0.24** | **0.28** | **-0.18** | **-0.31** | **-0.29** | **-0.15** |
| Autistic Burnout |  |  |  |  |  |  | 1.00 | **-0.48** | **-0.35** | **0.38** | **-0.39** | **-0.36** | **-0.33** | **-0.15** | **-0.21** | **-0.32** | **-0.24** | **-0.40** | **-0.42** | -0.37 | **0.32** | **0.37** | **-0.30** | **-0.48** | **-0.45** | **-0.19** |
| Community Participation |  |  |  |  |  |  |  | 1.00 | **0.43** | **-0.47** | **0.40** | **0.31** | **0.55** | **0.34** | **0.44** | **0.48** | **0.35** | **0.63** | **0.52** | **0.55** | **-0.41** | **-0.41** | **0.34** | **0.39** | **0.45** | **0.29** |
| Self-Determination |  |  |  |  |  |  |  |  | 1.00 | **-0.33** | **0.44** | **0.41** | **0.43** | **0.30** | **0.37** | **0.45** | **0.29** | **0.33** | **0.37** | 0.19 | **-0.24** | **-0.21** | **0.29** | **0.39** | **0.42** | **0.50** |
| Barriers to Communication |  |  |  |  |  |  |  |  |  | 1.00 | **-0.16** | -0.09 | **-0.35** | **-0.16** | **-0.31** | **-0.29** | **-0.22** | **-0.47** | **-0.46** | -0.16 | **0.26** | **0.38** | **-0.32** | **-0.35** | **-0.45** | **-0.38** |
| Overall Employment Satisfaction |  |  |  |  |  |  |  |  |  |  | 1.00 | **0.82** | **0.46** | **0.36** | **0.38** | **0.40** | **0.35** | **0.35** | **0.41** | 0.20 | **-0.27** | **-0.18** | **0.26** | **0.26** | **0.30** | **0.22** |
| Job Satisfaction (in an organization) |  |  |  |  |  |  |  |  |  |  |  | 1.00 | **0.38** | **0.21** | **0.30** | **0.38** | **0.26** | **0.36** | **0.36** | 0.03 | **-0.21** | **-0.17** | **0.26** | **0.20** | **0.28** | **0.20** |
| Total Social Support |  |  |  |  |  |  |  |  |  |  |  |  | 1.00 | **0.80** | **0.82** | **0.78** | **0.82** | **0.68** | **0.54** | **0.45** | **-0.32** | **-0.28** | **0.25** | **0.26** | **0.31** | **0.24** |
| Companionship |  |  |  |  |  |  |  |  |  |  |  |  |  | 1.00 | **0.66** | **0.47** | **0.60** | **0.38** | **0.35** | 0.29 | **-0.17** | -0.12 | **0.13** | 0.08 | 0.11 | **0.14** |
| Emotional Support |  |  |  |  |  |  |  |  |  |  |  |  |  |  | 1.00 | **0.68** | **0.49** | **0.49** | **0.44** | 0.23 | **-0.22** | **-0.26** | **0.24** | **0.23** | **0.30** | **0.31** |
| Informational Support |  |  |  |  |  |  |  |  |  |  |  |  |  |  |  | 1.00 | **0.50** | **0.48** | **0.47** | **0.44** | **-0.26** | **-0.27** | **0.30** | **0.33** | **0.36** | **0.22** |
| Practical Support |  |  |  |  |  |  |  |  |  |  |  |  |  |  |  |  | 1.00 | **0.41** | **0.44** | 0.26 | **-0.28** | **-0.21** | **0.15** | **0.16** | **0.18** | 0.08 |
| Lack of Isolation |  |  |  |  |  |  |  |  |  |  |  |  |  |  |  |  |  | 1.00 | **0.45** | **0.44** | **-0.31** | **-0.29** | **0.23** | **0.28** | **0.34** | **0.27** |
| Adequacy of Supports |  |  |  |  |  |  |  |  |  |  |  |  |  |  |  |  |  |  | 1.00 | **0.46** | **-0.51** | **-0.47** | **0.43** | **0.52** | **0.52** | **0.35** |
| Satisfaction with Disability Services |  |  |  |  |  |  |  |  |  |  |  |  |  |  |  |  |  |  |  | 1.00 | **-0.46** | -0.30 | 0.02 | 0.32 | 0.13 | 0.33 |
| Unmet Healthcare Needs |  |  |  |  |  |  |  |  |  |  |  |  |  |  |  |  |  |  |  |  | 1.00 | **0.61** | **-0.40** | **-0.51** | **-0.44** | **-0.28** |
| Barriers to Healthcare |  |  |  |  |  |  |  |  |  |  |  |  |  |  |  |  |  |  |  |  |  | 1.00 | **-0.45** | **-0.64** | **-0.62** | **-0.29** |
| Patient-Provider Satisfaction |  |  |  |  |  |  |  |  |  |  |  |  |  |  |  |  |  |  |  |  |  |  | 1.00 | **0.48** | **0.64** | **0.29** |
| Individual Level Healthcare Self-Efficacy |  |  |  |  |  |  |  |  |  |  |  |  |  |  |  |  |  |  |  |  |  |  |  | 1.00 | **0.77** | **0.34** |
| Relation-dependent Healthcare Self-Efficacy |  |  |  |  |  |  |  |  |  |  |  |  |  |  |  |  |  |  |  |  |  |  |  |  | 1.00 | **0.42** |
| Choices and Decisions |  |  |  |  |  |  |  |  |  |  |  |  |  |  |  |  |  |  |  |  |  |  |  |  |  | 1.00 |

Statistically significant associations (p < 0.05) are in bold. Color scale: dark blue (≥0.70 positive), medium blue (0.30–0.69), light blue (0.10–0.29), white (<0.10), light red (−0.10 to −0.29), medium red (−0.30 to −0.69).

**Supplement E2:**
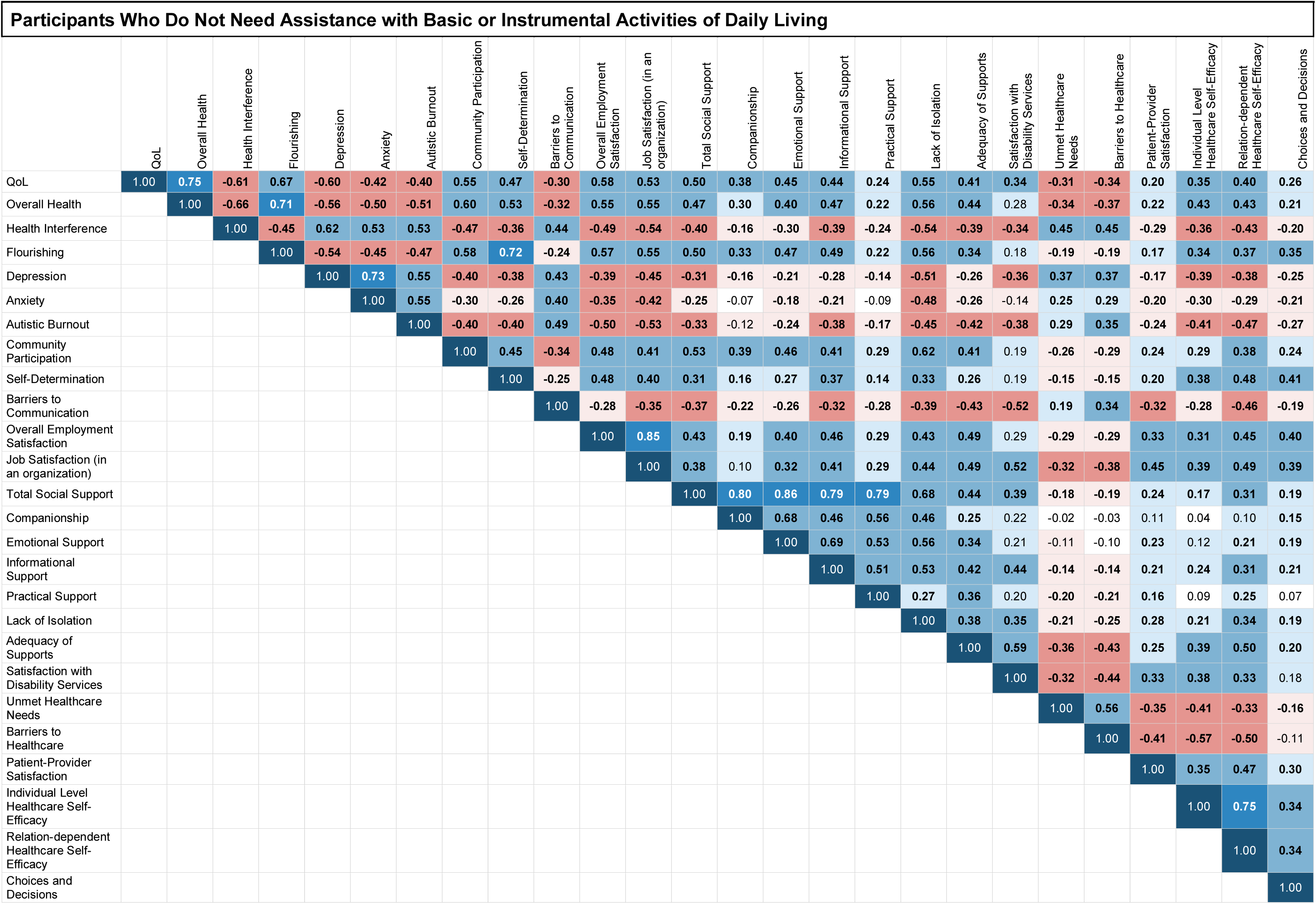
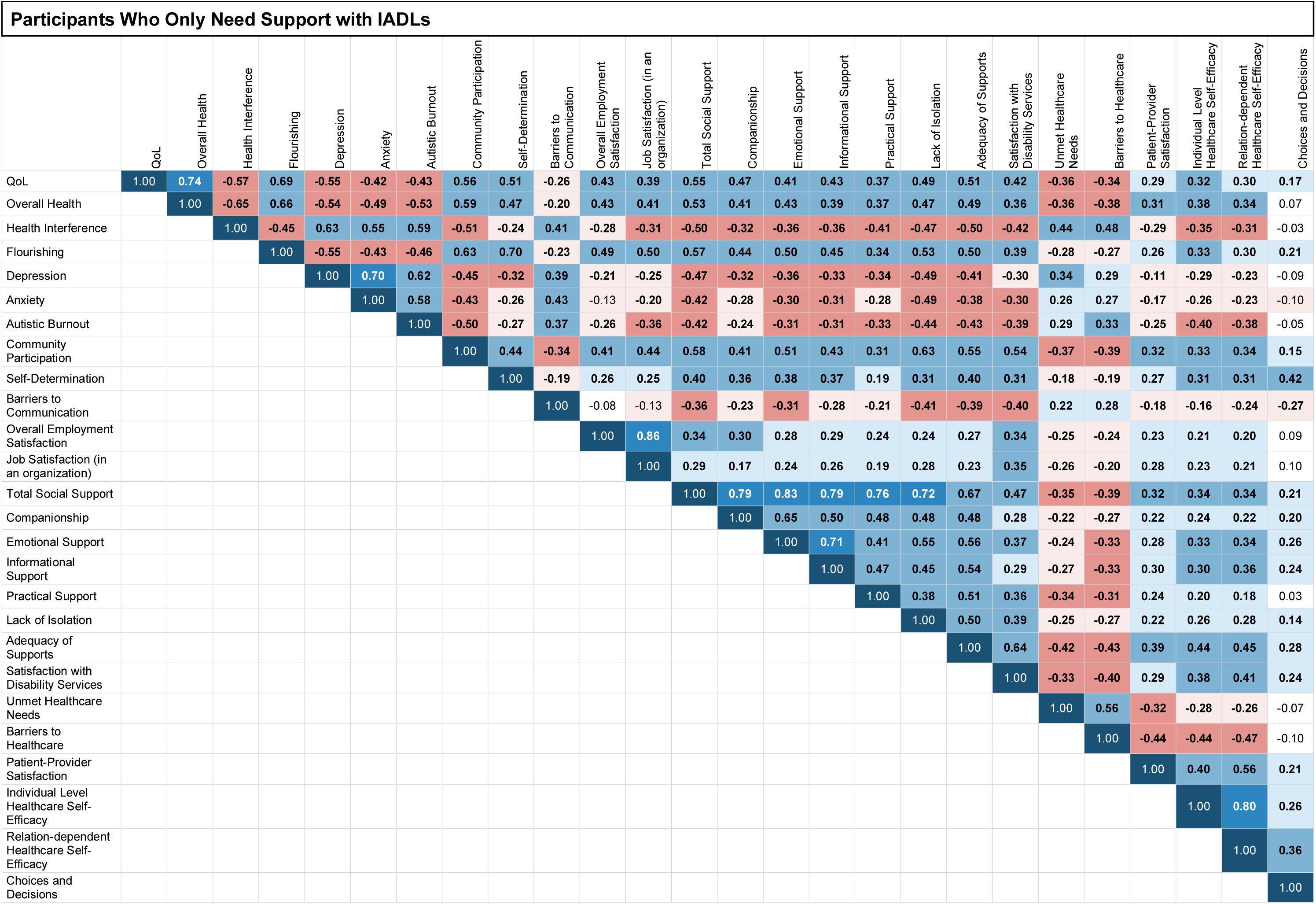
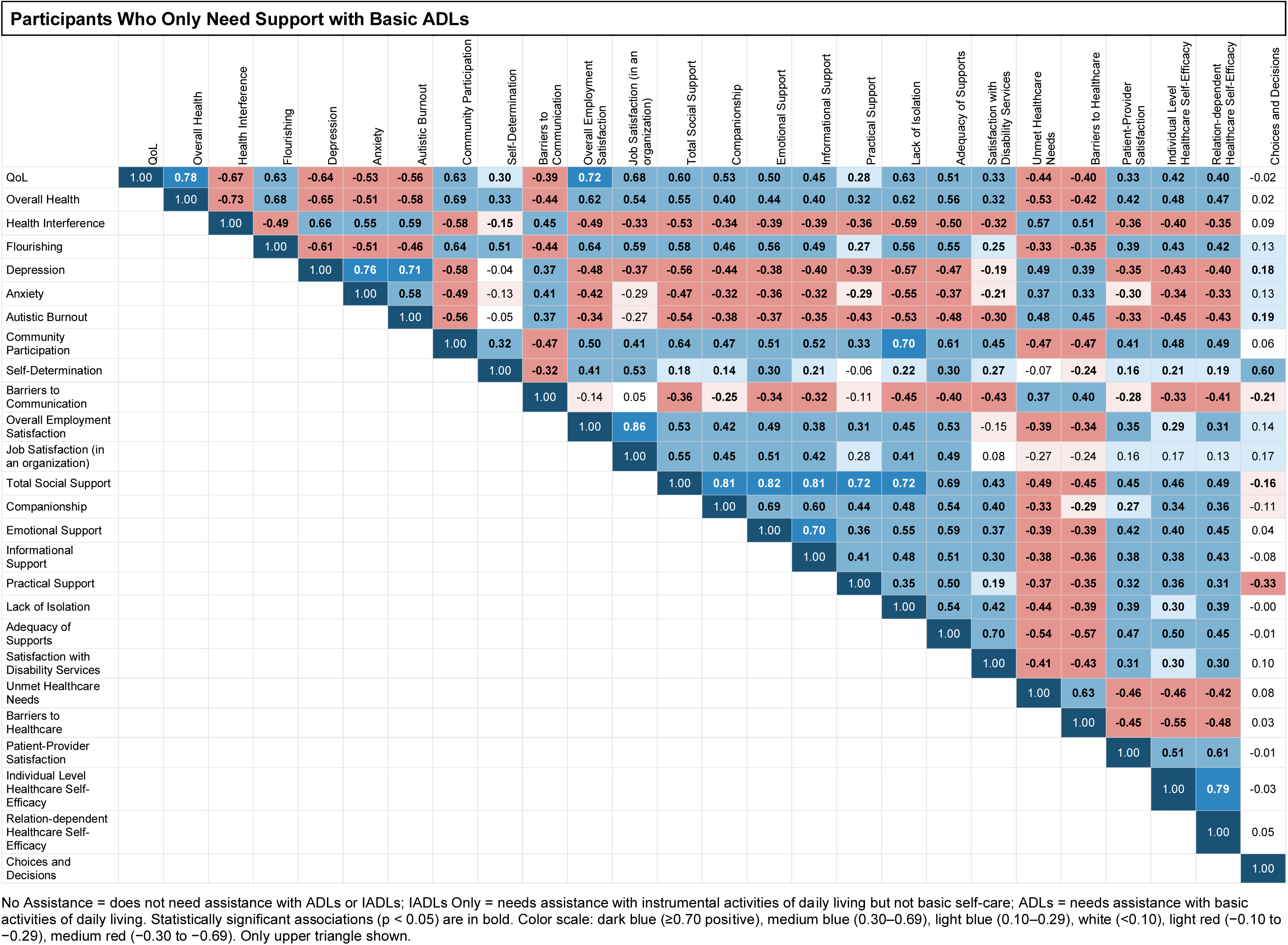
Correlations Between Outcomes, Divided by Need For Support for Basic and Instrumental Activities of Daily Living.

| Participants Who Do Not Need Assistance with Basic or Instrumental Activities of Daily Living |  |  |  |  |  |  |  |  |  |  |  |  |  |  |  |  |  |  |  |  |  |  |  |  |  |  |
|  | QoL | Overall Health | Health Interference | Flourishing | Depression | Anxiety | Autistic Burnout | Community Participation | Self-Determination | Barriers to Communication | Overall Employment Satisfaction | Job Satisfaction (in an organization) | Total Social Support | Companionship | Emotional Support | Informational Support | Practical Support | Lack of Isolation | Adequacy of Supports | Satisfaction with Disability Services | Unmet Healthcare Needs | Barriers to Healthcare | Patient-Provider Satisfaction | Individual Level Healthcare Self-Efficacy | Relation-dependent Healthcare Self-Efficacy | Choices and Decisions |
|---|---|---|---|---|---|---|---|---|---|---|---|---|---|---|---|---|---|---|---|---|---|---|---|---|---|---|
| QoL | 1.00 | **0.75** | **-0.61** | **0.67** | **-0.60** | **-0.42** | **-0.40** | **0.55** | **0.47** | **-0.30** | **0.58** | **0.53** | **0.50** | **0.38** | **0.45** | **0.44** | **0.24** | **0.55** | **0.41** | **0.34** | **-0.31** | **-0.34** | **0.20** | **0.35** | **0.40** | **0.26** |
| Overall Health |  | 1.00 | **-0.66** | **0.71** | **-0.56** | **-0.50** | **-0.51** | **0.60** | **0.53** | **-0.32** | **0.55** | **0.55** | **0.47** | **0.30** | **0.40** | **0.47** | **0.22** | **0.56** | **0.44** | 0.28 | **-0.34** | **-0.37** | **0.22** | **0.43** | **0.43** | **0.21** |
| Health Interference |  |  | 1.00 | **-0.45** | **0.62** | **0.53** | **0.53** | **-0.47** | **-0.36** | **0.44** | **-0.49** | **-0.54** | **-0.40** | **-0.16** | **-0.30** | **-0.39** | **-0.24** | **-0.54** | **-0.39** | **-0.34** | **0.45** | **0.45** | **-0.29** | **-0.36** | **-0.43** | **-0.20** |
| Flourishing |  |  |  | 1.00 | **-0.54** | **-0.45** | **-0.47** | **0.58** | **0.72** | **-0.24** | **0.57** | **0.55** | **0.50** | **0.33** | **0.47** | **0.49** | **0.22** | **0.56** | **0.34** | 0.18 | **-0.19** | **-0.19** | **0.17** | **0.34** | **0.37** | **0.35** |
| Depression |  |  |  |  | 1.00 | **0.73** | **0.55** | **-0.40** | **-0.38** | **0.43** | **-0.39** | **-0.45** | **-0.31** | **-0.16** | **-0.21** | **-0.28** | **-0.14** | **-0.51** | **-0.26** | **-0.36** | **0.37** | **0.37** | **-0.17** | **-0.39** | **-0.38** | **-0.25** |
| Anxiety |  |  |  |  |  | 1.00 | **0.55** | **-0.30** | **-0.26** | **0.40** | **-0.35** | **-0.42** | **-0.25** | -0.07 | **-0.18** | **-0.21** | -0.09 | **-0.48** | **-0.26** | -0.14 | **0.25** | **0.29** | **-0.20** | **-0.30** | **-0.29** | **-0.21** |
| Autistic Burnout |  |  |  |  |  |  | 1.00 | **-0.40** | **-0.40** | **0.49** | **-0.50** | **-0.53** | **-0.33** | -0.12 | **-0.24** | **-0.38** | **-0.17** | **-0.45** | **-0.42** | **-0.38** | **0.29** | **0.35** | **-0.24** | **-0.41** | **-0.47** | **-0.27** |
| Community Participation |  |  |  |  |  |  |  | 1.00 | **0.45** | **-0.34** | **0.48** | **0.41** | **0.53** | **0.39** | **0.46** | **0.41** | **0.29** | **0.62** | **0.41** | 0.19 | **-0.26** | **-0.29** | **0.24** | **0.29** | **0.38** | **0.24** |
| Self-Determination |  |  |  |  |  |  |  |  | 1.00 | **-0.25** | **0.48** | **0.40** | **0.31** | **0.16** | **0.27** | **0.37** | **0.14** | **0.33** | **0.26** | 0.19 | **-0.15** | **-0.15** | **0.20** | **0.38** | **0.48** | **0.41** |
| Barriers to Communication |  |  |  |  |  |  |  |  |  | 1.00 | **-0.28** | **-0.35** | **-0.37** | **-0.22** | **-0.26** | **-0.32** | **-0.28** | **-0.39** | **-0.43** | **-0.52** | **0.19** | **0.34** | **-0.32** | **-0.28** | **-0.46** | **-0.19** |
| Overall Employment Satisfaction |  |  |  |  |  |  |  |  |  |  | 1.00 | **0.85** | **0.43** | **0.19** | **0.40** | **0.46** | **0.29** | **0.43** | **0.49** | 0.29 | **-0.29** | **-0.29** | **0.33** | **0.31** | **0.45** | **0.40** |
| Job Satisfaction (in an organization) |  |  |  |  |  |  |  |  |  |  |  | 1.00 | **0.38** | 0.10 | **0.32** | **0.41** | **0.29** | **0.44** | **0.49** | **0.52** | **-0.32** | **-0.38** | **0.45** | **0.39** | **0.49** | **0.39** |
| Total Social Support |  |  |  |  |  |  |  |  |  |  |  |  | 1.00 | **0.80** | **0.86** | **0.79** | **0.79** | **0.68** | **0.44** | **0.39** | **-0.18** | **-0.19** | **0.24** | **0.17** | **0.31** | **0.19** |
| Companionship |  |  |  |  |  |  |  |  |  |  |  |  |  | 1.00 | **0.68** | **0.46** | **0.56** | **0.46** | **0.25** | 0.22 | -0.02 | -0.03 | 0.11 | 0.04 | 0.10 | **0.15** |
| Emotional Support |  |  |  |  |  |  |  |  |  |  |  |  |  |  | 1.00 | **0.69** | **0.53** | **0.56** | **0.34** | 0.21 | -0.11 | -0.10 | **0.23** | 0.12 | **0.21** | **0.19** |
| Informational Support |  |  |  |  |  |  |  |  |  |  |  |  |  |  |  | 1.00 | **0.51** | **0.53** | **0.42** | **0.44** | **-0.14** | **-0.14** | **0.21** | **0.24** | **0.31** | **0.21** |
| Practical Support |  |  |  |  |  |  |  |  |  |  |  |  |  |  |  |  | 1.00 | **0.27** | **0.36** | 0.20 | **-0.20** | **-0.21** | **0.16** | 0.09 | **0.25** | 0.07 |
| Lack of Isolation |  |  |  |  |  |  |  |  |  |  |  |  |  |  |  |  |  | 1.00 | **0.38** | **0.35** | **-0.21** | **-0.25** | **0.28** | **0.21** | **0.34** | **0.19** |
| Adequacy of Supports |  |  |  |  |  |  |  |  |  |  |  |  |  |  |  |  |  |  | 1.00 | **0.59** | **-0.36** | **-0.43** | **0.25** | **0.39** | **0.50** | **0.20** |
| Satisfaction with Disability Services |  |  |  |  |  |  |  |  |  |  |  |  |  |  |  |  |  |  |  | 1.00 | **-0.32** | **-0.44** | **0.33** | **0.38** | **0.33** | 0.18 |
| Unmet Healthcare Needs |  |  |  |  |  |  |  |  |  |  |  |  |  |  |  |  |  |  |  |  | 1.00 | **0.56** | **-0.35** | **-0.41** | **-0.33** | **-0.16** |
| Barriers to Healthcare |  |  |  |  |  |  |  |  |  |  |  |  |  |  |  |  |  |  |  |  |  | 1.00 | **-0.41** | **-0.57** | **-0.50** | -0.11 |
| Patient-Provider Satisfaction |  |  |  |  |  |  |  |  |  |  |  |  |  |  |  |  |  |  |  |  |  |  | 1.00 | **0.35** | **0.47** | **0.30** |
| Individual Level Healthcare Self-Efficacy |  |  |  |  |  |  |  |  |  |  |  |  |  |  |  |  |  |  |  |  |  |  |  | 1.00 | **0.75** | **0.34** |
| Relation-dependent Healthcare Self-Efficacy |  |  |  |  |  |  |  |  |  |  |  |  |  |  |  |  |  |  |  |  |  |  |  |  | 1.00 | **0.34** |
| Choices and Decisions |  |  |  |  |  |  |  |  |  |  |  |  |  |  |  |  |  |  |  |  |  |  |  |  |  | 1.00 |
| Participants Who Only Need Support with IADLs |  |  |  |  |  |  |  |  |  |  |  |  |  |  |  |  |  |  |  |  |  |  |  |  |  |  |
|  | QoL | Overall Health | Health Interference | Flourishing | Depression | Anxiety | Autistic Burnout | Community Participation | Self-Determination | Barriers to Communication | Overall Employment Satisfaction | Job Satisfaction (in an organization) | Total Social Support | Companionship | Emotional Support | Informational Support | Practical Support | Lack of Isolation | Adequacy of Supports | Satisfaction with Disability Services | Unmet Healthcare Needs | Barriers to Healthcare | Patient-Provider Satisfaction | Individual Level Healthcare Self-Efficacy | Relation-dependent Healthcare Self-Efficacy | Choices and Decisions |
| QoL | 1.00 | **0.74** | **-0.57** | **0.69** | **-0.55** | **-0.42** | **-0.43** | **0.56** | **0.51** | **-0.26** | **0.43** | **0.39** | **0.55** | **0.47** | **0.41** | **0.43** | **0.37** | **0.49** | **0.51** | **0.42** | **-0.36** | **-0.34** | **0.29** | **0.32** | **0.30** | **0.17** |
| Overall Health |  | 1.00 | **-0.65** | **0.66** | **-0.54** | **-0.49** | **-0.53** | **0.59** | **0.47** | **-0.20** | **0.43** | **0.41** | **0.53** | **0.41** | **0.43** | **0.39** | **0.37** | **0.47** | **0.49** | **0.36** | **-0.36** | **-0.38** | **0.31** | **0.38** | **0.34** | 0.07 |
| Health Interference |  |  | 1.00 | **-0.45** | **0.63** | **0.55** | **0.59** | **-0.51** | **-0.24** | **0.41** | **-0.28** | **-0.31** | **-0.50** | **-0.32** | **-0.36** | **-0.36** | **-0.41** | **-0.47** | **-0.50** | **-0.42** | **0.44** | **0.48** | **-0.29** | **-0.35** | **-0.31** | -0.03 |
| Flourishing |  |  |  | 1.00 | **-0.55** | **-0.43** | **-0.46** | **0.63** | **0.70** | **-0.23** | **0.49** | **0.50** | **0.57** | **0.44** | **0.50** | **0.45** | **0.34** | **0.53** | **0.50** | **0.39** | **-0.28** | **-0.27** | **0.26** | **0.33** | **0.30** | **0.21** |
| Depression |  |  |  |  | 1.00 | **0.70** | **0.62** | **-0.45** | **-0.32** | **0.39** | **-0.21** | **-0.25** | **-0.47** | **-0.32** | **-0.36** | **-0.33** | **-0.34** | **-0.49** | **-0.41** | **-0.30** | **0.34** | **0.29** | **-0.11** | **-0.29** | **-0.23** | -0.09 |
| Anxiety |  |  |  |  |  | 1.00 | **0.58** | **-0.43** | **-0.26** | **0.43** | -0.13 | **-0.20** | **-0.42** | **-0.28** | **-0.30** | **-0.31** | **-0.28** | **-0.49** | **-0.38** | **-0.30** | **0.26** | **0.27** | **-0.17** | **-0.26** | **-0.23** | -0.10 |
| Autistic Burnout |  |  |  |  |  |  | 1.00 | **-0.50** | **-0.27** | **0.37** | **-0.26** | **-0.36** | **-0.42** | **-0.24** | **-0.31** | **-0.31** | **-0.33** | **-0.44** | **-0.43** | **-0.39** | **0.29** | **0.33** | **-0.25** | **-0.40** | **-0.38** | -0.05 |
| Community Participation |  |  |  |  |  |  |  | 1.00 | **0.44** | **-0.34** | **0.41** | **0.44** | **0.58** | **0.41** | **0.51** | **0.43** | **0.31** | **0.63** | **0.55** | **0.54** | **-0.37** | **-0.39** | **0.32** | **0.33** | **0.34** | **0.15** |
| Self-Determination |  |  |  |  |  |  |  |  | 1.00 | **-0.19** | **0.26** | **0.25** | **0.40** | **0.36** | **0.38** | **0.37** | **0.19** | **0.31** | **0.40** | **0.31** | **-0.18** | **-0.19** | **0.27** | **0.31** | **0.31** | **0.42** |
| Barriers to Communication |  |  |  |  |  |  |  |  |  | 1.00 | -0.08 | -0.13 | **-0.36** | **-0.23** | **-0.31** | **-0.28** | **-0.21** | **-0.41** | **-0.39** | **-0.40** | **0.22** | **0.28** | **-0.18** | **-0.16** | **-0.24** | **-0.27** |
| Overall Employment Satisfaction |  |  |  |  |  |  |  |  |  |  | 1.00 | **0.86** | **0.34** | **0.30** | **0.28** | **0.29** | **0.24** | **0.24** | **0.27** | **0.34** | **-0.25** | **-0.24** | **0.23** | **0.21** | **0.20** | 0.09 |
| Job Satisfaction (in an organization) |  |  |  |  |  |  |  |  |  |  |  | 1.00 | **0.29** | **0.17** | **0.24** | **0.26** | **0.19** | **0.28** | **0.23** | **0.35** | **-0.26** | **-0.20** | **0.28** | **0.23** | **0.21** | 0.10 |
| Total Social Support |  |  |  |  |  |  |  |  |  |  |  |  | 1.00 | **0.79** | **0.83** | **0.79** | **0.76** | **0.72** | **0.67** | **0.47** | **-0.35** | **-0.39** | **0.32** | **0.34** | **0.34** | **0.21** |
| Companionship |  |  |  |  |  |  |  |  |  |  |  |  |  | 1.00 | **0.65** | **0.50** | **0.48** | **0.48** | **0.48** | **0.28** | **-0.22** | **-0.27** | **0.22** | **0.24** | **0.22** | **0.20** |
| Emotional Support |  |  |  |  |  |  |  |  |  |  |  |  |  |  | 1.00 | **0.71** | **0.41** | **0.55** | **0.56** | **0.37** | **-0.24** | **-0.33** | **0.28** | **0.33** | **0.34** | **0.26** |
| Informational Support |  |  |  |  |  |  |  |  |  |  |  |  |  |  |  | 1.00 | **0.47** | **0.45** | **0.54** | **0.29** | **-0.27** | **-0.33** | **0.30** | **0.30** | **0.36** | **0.24** |
| Practical Support |  |  |  |  |  |  |  |  |  |  |  |  |  |  |  |  | 1.00 | **0.38** | **0.51** | **0.36** | **-0.34** | **-0.31** | **0.24** | **0.20** | **0.18** | 0.03 |
| Lack of Isolation |  |  |  |  |  |  |  |  |  |  |  |  |  |  |  |  |  | 1.00 | **0.50** | **0.39** | **-0.25** | **-0.27** | **0.22** | **0.26** | **0.28** | **0.14** |
| Adequacy of Supports |  |  |  |  |  |  |  |  |  |  |  |  |  |  |  |  |  |  | 1.00 | **0.64** | **-0.42** | **-0.43** | **0.39** | **0.44** | **0.45** | **0.28** |
| Satisfaction with Disability Services |  |  |  |  |  |  |  |  |  |  |  |  |  |  |  |  |  |  |  | 1.00 | **-0.33** | **-0.40** | **0.29** | **0.38** | **0.41** | **0.24** |
| Unmet Healthcare Needs |  |  |  |  |  |  |  |  |  |  |  |  |  |  |  |  |  |  |  |  | 1.00 | **0.56** | **-0.32** | **-0.28** | **-0.26** | -0.07 |
| Barriers to Healthcare |  |  |  |  |  |  |  |  |  |  |  |  |  |  |  |  |  |  |  |  |  | 1.00 | **-0.44** | **-0.44** | **-0.47** | -0.10 |
| Patient-Provider Satisfaction |  |  |  |  |  |  |  |  |  |  |  |  |  |  |  |  |  |  |  |  |  |  | 1.00 | **0.40** | **0.56** | **0.21** |
| Individual Level Healthcare Self-Efficacy |  |  |  |  |  |  |  |  |  |  |  |  |  |  |  |  |  |  |  |  |  |  |  | 1.00 | **0.80** | **0.26** |
| Relation-dependent Healthcare Self-Efficacy |  |  |  |  |  |  |  |  |  |  |  |  |  |  |  |  |  |  |  |  |  |  |  |  | 1.00 | **0.36** |
| Choices and Decisions |  |  |  |  |  |  |  |  |  |  |  |  |  |  |  |  |  |  |  |  |  |  |  |  |  | 1.00 |
| Participants Who Only Need Support with Basic ADLs |  |  |  |  |  |  |  |  |  |  |  |  |  |  |  |  |  |  |  |  |  |  |  |  |  |  |
|  | QoL | Overall Health | Health Interference | Flourishing | Depression | Anxiety | Autistic Burnout | Community Participation | Self-Determination | Barriers to Communication | Overall Employment Satisfaction | Job Satisfaction (in an organization) | Total Social Support | Companionship | Emotional Support | Informational Support | Practical Support | Lack of Isolation | Adequacy of Supports | Satisfaction with Disability Services | Unmet Healthcare Needs | Barriers to Healthcare | Patient-Provider Satisfaction | Individual Level Healthcare Self-Efficacy | Relation-dependent Healthcare Self-Efficacy | Choices and Decisions |
| QoL | 1.00 | **0.78** | **-0.67** | **0.63** | **-0.64** | **-0.53** | **-0.56** | **0.63** | **0.30** | **-0.39** | **0.72** | **0.68** | **0.60** | **0.53** | **0.50** | **0.45** | **0.28** | **0.63** | **0.51** | **0.33** | **-0.44** | **-0.40** | **0.33** | **0.42** | **0.40** | -0.02 |
| Overall Health |  | 1.00 | **-0.73** | **0.68** | **-0.65** | **-0.51** | **-0.58** | **0.69** | **0.33** | **-0.44** | **0.62** | **0.54** | **0.55** | **0.40** | **0.44** | **0.40** | **0.32** | **0.62** | **0.56** | **0.32** | **-0.53** | **-0.42** | **0.42** | **0.48** | **0.47** | 0.02 |
| Health Interference |  |  | 1.00 | **-0.49** | **0.66** | **0.55** | **0.59** | **-0.58** | **-0.15** | **0.45** | **-0.49** | **-0.33** | **-0.53** | **-0.34** | **-0.39** | **-0.39** | **-0.36** | **-0.59** | **-0.50** | **-0.32** | **0.57** | **0.51** | **-0.36** | **-0.40** | **-0.35** | 0.09 |
| Flourishing |  |  |  | 1.00 | **-0.61** | **-0.51** | **-0.46** | **0.64** | **0.51** | **-0.44** | **0.64** | **0.59** | **0.58** | **0.46** | **0.56** | **0.49** | **0.27** | **0.56** | **0.55** | **0.25** | **-0.33** | **-0.35** | **0.39** | **0.43** | **0.42** | 0.13 |
| Depression |  |  |  |  | 1.00 | **0.76** | **0.71** | **-0.58** | -0.04 | **0.37** | **-0.48** | **-0.37** | **-0.56** | **-0.44** | **-0.38** | **-0.40** | **-0.39** | **-0.57** | **-0.47** | **-0.19** | **0.49** | **0.39** | **-0.35** | **-0.43** | **-0.40** | **0.18** |
| Anxiety |  |  |  |  |  | 1.00 | **0.58** | **-0.49** | -0.13 | **0.41** | **-0.42** | -0.29 | **-0.47** | **-0.32** | **-0.36** | **-0.32** | **-0.29** | **-0.55** | **-0.37** | **-0.21** | **0.37** | **0.33** | **-0.30** | **-0.34** | **-0.33** | 0.13 |
| Autistic Burnout |  |  |  |  |  |  | 1.00 | **-0.56** | -0.05 | **0.37** | **-0.34** | -0.27 | **-0.54** | **-0.38** | **-0.37** | **-0.35** | **-0.43** | **-0.53** | **-0.48** | **-0.30** | **0.48** | **0.45** | **-0.33** | **-0.45** | **-0.43** | **0.19** |
| Community Participation |  |  |  |  |  |  |  | 1.00 | **0.32** | **-0.47** | **0.50** | **0.41** | **0.64** | **0.47** | **0.51** | **0.52** | **0.33** | **0.70** | **0.61** | **0.45** | **-0.47** | **-0.47** | **0.41** | **0.48** | **0.49** | 0.06 |
| Self-Determination |  |  |  |  |  |  |  |  | 1.00 | **-0.32** | **0.41** | **0.53** | **0.18** | **0.14** | **0.30** | **0.21** | -0.06 | **0.22** | **0.30** | **0.27** | -0.07 | **-0.24** | **0.16** | **0.21** | **0.19** | **0.60** |
| Barriers to Communication |  |  |  |  |  |  |  |  |  | 1.00 | -0.14 | 0.05 | **-0.36** | **-0.25** | **-0.34** | **-0.32** | -0.11 | **-0.45** | **-0.40** | **-0.43** | **0.37** | **0.40** | **-0.28** | **-0.33** | **-0.41** | **-0.21** |
| Overall Employment Satisfaction |  |  |  |  |  |  |  |  |  |  | 1.00 | **0.86** | **0.53** | **0.42** | **0.49** | **0.38** | **0.31** | **0.45** | **0.53** | -0.15 | **-0.39** | **-0.34** | **0.35** | **0.29** | **0.31** | 0.14 |
| Job Satisfaction (in an organization) |  |  |  |  |  |  |  |  |  |  |  | 1.00 | **0.55** | **0.45** | **0.51** | **0.42** | 0.28 | **0.41** | **0.49** | 0.08 | -0.27 | -0.24 | 0.16 | 0.17 | 0.13 | 0.17 |
| Total Social Support |  |  |  |  |  |  |  |  |  |  |  |  | 1.00 | **0.81** | **0.82** | **0.81** | **0.72** | **0.72** | **0.69** | **0.43** | **-0.49** | **-0.45** | **0.45** | **0.46** | **0.49** | **-0.16** |
| Companionship |  |  |  |  |  |  |  |  |  |  |  |  |  | 1.00 | **0.69** | **0.60** | **0.44** | **0.48** | **0.54** | **0.40** | **-0.33** | **-0.29** | **0.27** | **0.34** | **0.36** | -0.11 |
| Emotional Support |  |  |  |  |  |  |  |  |  |  |  |  |  |  | 1.00 | **0.70** | **0.36** | **0.55** | **0.59** | **0.37** | **-0.39** | **-0.39** | **0.42** | **0.40** | **0.45** | 0.04 |
| Informational Support |  |  |  |  |  |  |  |  |  |  |  |  |  |  |  | 1.00 | **0.41** | **0.48** | **0.51** | **0.30** | **-0.38** | **-0.36** | **0.38** | **0.38** | **0.43** | -0.08 |
| Practical Support |  |  |  |  |  |  |  |  |  |  |  |  |  |  |  |  | 1.00 | **0.35** | **0.50** | **0.19** | **-0.37** | **-0.35** | **0.32** | **0.36** | **0.31** | **-0.33** |
| Lack of Isolation |  |  |  |  |  |  |  |  |  |  |  |  |  |  |  |  |  | 1.00 | **0.54** | **0.42** | **-0.44** | **-0.39** | **0.39** | **0.30** | **0.39** | -0.00 |
| Adequacy of Supports |  |  |  |  |  |  |  |  |  |  |  |  |  |  |  |  |  |  | 1.00 | **0.70** | **-0.54** | **-0.57** | **0.47** | **0.50** | **0.45** | -0.01 |
| Satisfaction with Disability Services |  |  |  |  |  |  |  |  |  |  |  |  |  |  |  |  |  |  |  | 1.00 | **-0.41** | **-0.43** | **0.31** | **0.30** | **0.30** | 0.10 |
| Unmet Healthcare Needs |  |  |  |  |  |  |  |  |  |  |  |  |  |  |  |  |  |  |  |  | 1.00 | **0.63** | **-0.46** | **-0.46** | **-0.42** | 0.08 |
| Barriers to Healthcare |  |  |  |  |  |  |  |  |  |  |  |  |  |  |  |  |  |  |  |  |  | 1.00 | **-0.45** | **-0.55** | **-0.48** | 0.03 |
| Patient-Provider Satisfaction |  |  |  |  |  |  |  |  |  |  |  |  |  |  |  |  |  |  |  |  |  |  | 1.00 | **0.51** | **0.61** | -0.01 |
| Individual Level Healthcare Self-Efficacy |  |  |  |  |  |  |  |  |  |  |  |  |  |  |  |  |  |  |  |  |  |  |  | 1.00 | **0.79** | -0.03 |
| Relation-dependent Healthcare Self-Efficacy |  |  |  |  |  |  |  |  |  |  |  |  |  |  |  |  |  |  |  |  |  |  |  |  | 1.00 | 0.05 |
| Choices and Decisions |  |  |  |  |  |  |  |  |  |  |  |  |  |  |  |  |  |  |  |  |  |  |  |  |  | 1.00 |

No Assistance = does not need assistance with ADLs or IADLs; IADLs Only = needs assistance with instrumental activities of daily living but not basic self-care; ADLs = needs assistance with basic activities of daily living. Statistically significant associations (p < 0.05) are in bold. Color scale: dark blue (≥0.70 positive), medium blue (0.30–0.69), light blue (0.10–0.29), white (<0.10), light red (−0.10 to −0.29), medium red (−0.30 to −0.69). Only upper triangle shown.

**Supplement E3:**
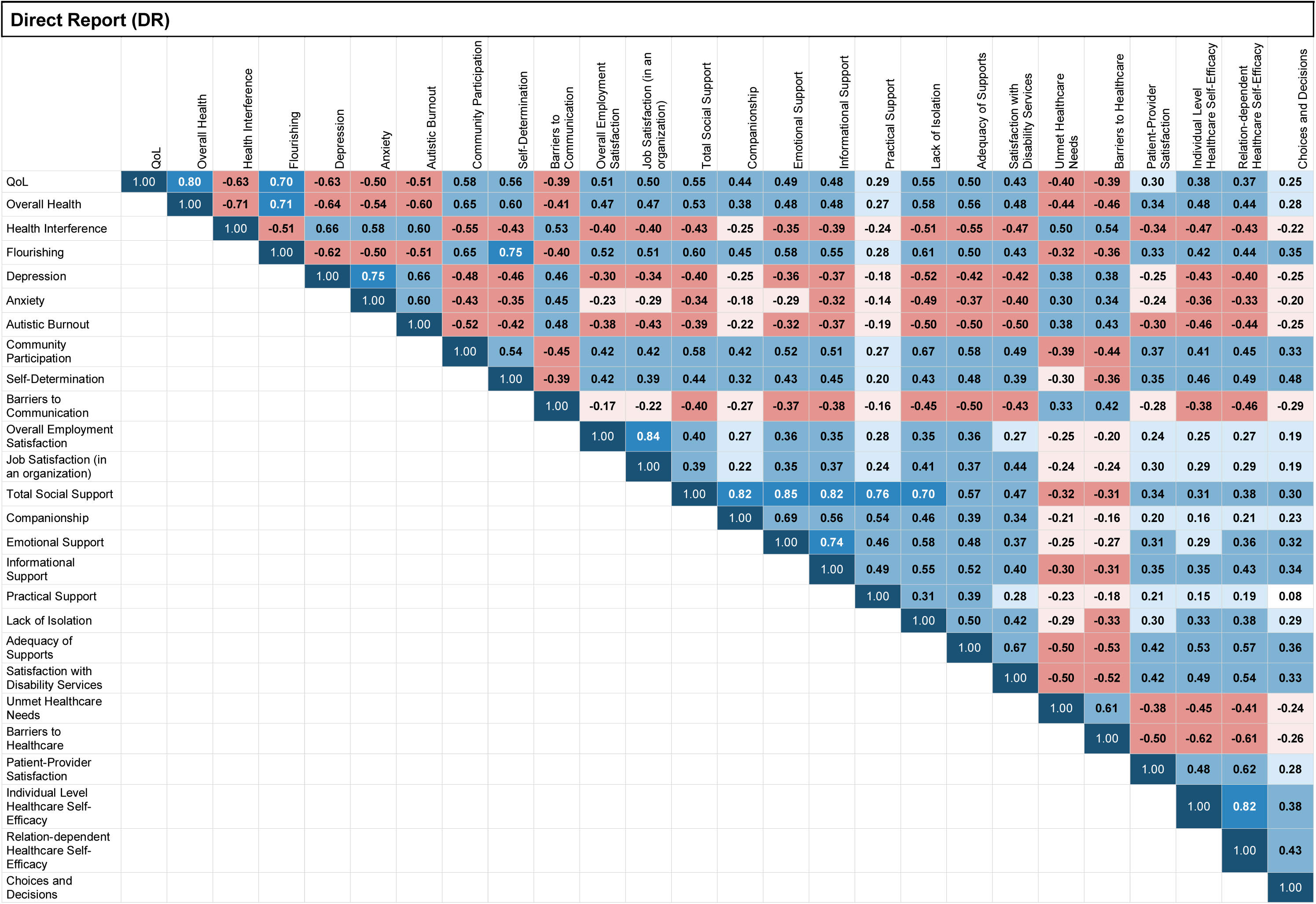
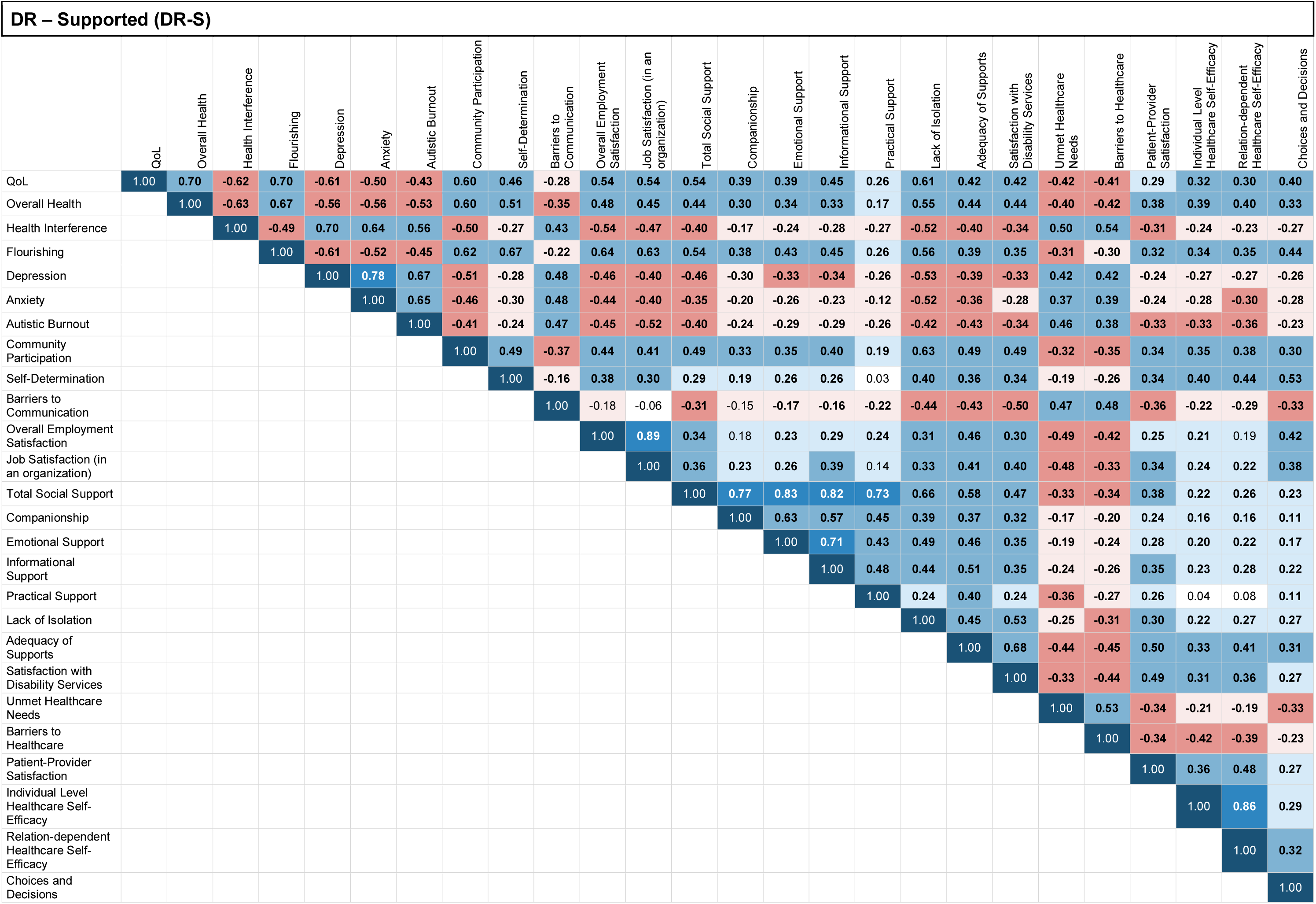
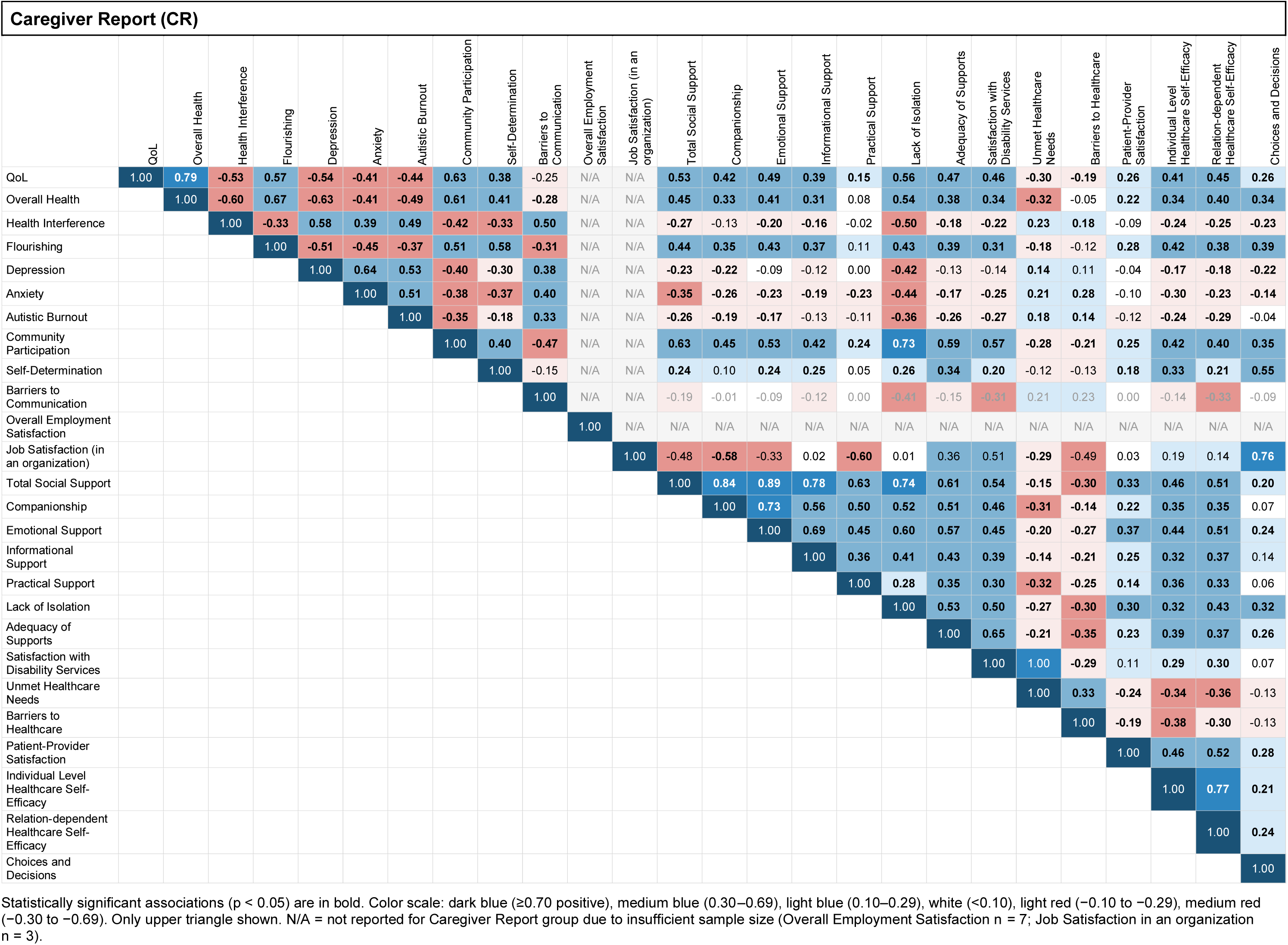
Correlations Between Outcomes, Divided by Reporter Type.

**Supplemental Table F1:** Responsiveness to Change by Subcohort.

| **Instrument** | **Disability Services (DDS)** |  |  |  | **Healthcare** |  |  |  | **Community** |  |  |  |
|---|---|---|---|---|---|---|---|---|---|---|---|---|
|  | **F** | **p** | **B Got Worse** | **B Got Better** | **F** | **p** | **B Got Worse** | **B Got Better** | **F** | **p** | **B Got Worse** | **B Got Better** |
| **Health and Quality of Life** |  |  |  |  |  |  |  |  |  |  |  |  |
| **Overall Health** | **9.93** | **<0.0001** | **-0.27** | **0.21** | **18.91** | **<0.0001** | **-0.24** | **0.17** | **56.13** | **<0.0001** | **-0.30** | **0.25** |
| **Quality of Life** | **11.29** | **<0.0001** | **-0.89** | **0.50** | **22.69** | **<0.0001** | **-0.93** | **0.48** | **26.85** | **<0.0001** | **-0.81** | **0.47** |
| **Social Engagement** |  |  |  |  |  |  |  |  |  |  |  |  |
| **Community Participation** | **15.17** | **<0.0001** | **-0.29** | **0.37** | **15.67** | **<0.0001** | **-0.32** | **0.30** | **39.95** | **<0.0001** | **-0.26** | **0.58** |
| **Choices and Decisions** | **8.23** | **0.0004** | **-0.60** | **0.15** | 2.01 | 0.1357 | -0.13 | 0.06 | **16.86** | **<0.0001** | **-0.43** | **0.27** |
| **Self Determination** | **13.37** | **<0.0001** | **-0.35** | **0.19** | **8.62** | **0.0002** | **-0.15** | **0.18** | **11.23** | **<0.0001** | **-0.27** | **0.12** |
| **Employment** |  |  |  |  |  |  |  |  |  |  |  |  |
| **Job Satisfaction Survey** | **3.34** | **0.0433** | **-0.58** | **0.20** | **8.25** | **0.0005** | **-0.33** | **0.06** | **18.20** | **<0.0001** | **-0.22** | **0.35** |
| **Overall Employment Satisfaction** | **6.69** | **0.0024** | **-1.01** | **0.63** | **13.71** | **<0.0001** | **-0.54** | **0.30** | **27.01** | **<0.0001** | **-0.38** | **0.66** |
| **Mental Health** |  |  |  |  |  |  |  |  |  |  |  |  |
| **Anxiety** | **16.68** | **<0.0001** | **0.43** | **-0.25** | **20.52** | **<0.0001** | **0.39** | **-0.10** | **32.95** | **<0.0001** | **0.27** | **-0.36** |
| **Depression** | **12.96** | **<0.0001** | **0.34** | **-0.26** | **22.41** | **<0.0001** | **0.28** | **-0.18** | **27.33** | **<0.0001** | **0.26** | **-0.25** |
| **Autistic Burnout** | **17.13** | **<0.0001** | **0.34** | **-0.36** | **24.65** | **<0.0001** | **0.34** | **-0.36** | **40.99** | **<0.0001** | **0.43** | **-0.49** |
| **Flourishing** | **14.64** | **<0.0001** | **-0.42** | **0.26** | **8.90** | **0.0002** | **-0.24** | **0.13** | **19.58** | **<0.0001** | **-0.34** | **0.26** |
| **Social Support** |  |  |  |  |  |  |  |  |  |  |  |  |
| **Social Support: Companionship** | **18.79** | **<0.0001** | **-0.46** | **0.25** | **13.83** | **<0.0001** | **-0.36** | **0.34** | **19.39** | **<0.0001** | **-0.31** | **0.45** |
| **Social Support: Emotional Support** | **6.32** | **0.0021** | **-0.35** | **0.25** | **11.69** | **<0.0001** | **-0.37** | **0.20** | **10.34** | **<0.0001** | **-0.47** | **0.24** |
| **Social Support: Informational Support** | **5.13** | **0.0066** | **-0.43** | **0.26** | **7.18** | **0.0009** | **-0.22** | **0.31** | **7.41** | **0.0007** | **-0.47** | **0.23** |
| **Social Support: Practical Support** | **5.46** | **0.0049** | **-0.84** | **0.20** | 2.61 | 0.0755 | -0.21 | 0.21 | **20.07** | **<0.0001** | **-0.43** | **0.62** |
| **Social Support: Isolation** | **8.90** | **0.0002** | **-0.38** | **0.03** | **19.87** | **<0.0001** | **-0.30** | **0.24** | **19.07** | **<0.0001** | **-0.27** | **0.25** |
| **Supports and Services** |  |  |  |  |  |  |  |  |  |  |  |  |
| **Barriers to Communication** | 0.74 | 0.4788 | 0.54 | -0.02 | **5.47** | **0.0047** | **0.34** | **-0.91** | **6.25** | **0.0022** | **0.64** | **-0.47** |
| **Satisfaction with Services: Support Staff** | **9.17** | **0.0002** | **-0.54** | **0.13** | **3.21** | **0.0468** | **-0.62** | **0.55** | 1.03 | 0.3716 | -0.49 | 0.02 |
| **Satisfaction with Services: Access** | **8.60** | **0.0003** | **-0.28** | **0.60** | 2.36 | 0.1023 | -0.37 | -0.01 | **4.15** | **0.0273** | **-0.82** | **0.20** |
| **Adequacy of Supports and Services** | 1.77 | 0.1722 | -0.09 | 0.21 | 1.98 | 0.1400 | -0.17 | 0.15 | **9.33** | **0.0001** | **-0.27** | **0.36** |
| **Healthcare Quality** |  |  |  |  |  |  |  |  |  |  |  |  |
| **Patient-Provider Communication** | **5.18** | **0.0064** | **-0.51** | **0.33** | **17.26** | **<0.0001** | **-0.32** | **0.54** | **19.42** | **<0.0001** | **-0.68** | **0.42** |
| **Healthcare Self-Efficacy: Individual Level** | 2.50 | 0.0845 | -0.42 | 0.37 | **5.96** | **0.0029** | **-0.30** | **0.51** | 2.72 | 0.0679 | -0.23 | 0.33 |
| **Healthcare Self-Efficacy: Relationship-Dependent** | **10.28** | **<0.0001** | **-0.95** | **0.37** | **9.02** | **0.0002** | **-0.37** | **0.79** | **4.73** | **0.0096** | **-0.43** | **0.41** |
| **Barriers to Healthcare** | **4.41** | **0.0134** | **1.41** | **-0.64** | 0.32 | 0.7256 | 0.22 | -0.19 | 0.23 | 0.7923 | 0.20 | 0.11 |
| **Unmet Healthcare Needs** | 1.39 | 0.2520 | 0.22 | -0.35 | 2.45 | 0.0883 | 0.44 | -0.18 | **7.06** | **0.0010** | **0.41** | **-0.74** |

DDS = Disability Services subcohort; HC = Healthcare subcohort; Comm = Community subcohort. F = F-statistic from OLS regression with cluster-robust standard errors. B (Got Worse) = mean difference in change score between participants who reported getting worse vs. those who stayed the same; B (Got Better) = same comparison for those who got better. Analyses used pooled longitudinal observations across two follow-up intervals; each participant contributes up to two observations per analysis. Bold values indicate statistical significance at p < 0.05. b Category absent in this subsample.

**Supplemental Table F2:** Responsiveness to Change by Support Needs.

| **Instrument** | **No Assistance Needed** |  |  |  | **IADLs Only** |  |  |  | **ADLs** |  |  |  |
|---|---|---|---|---|---|---|---|---|---|---|---|---|
|  | **F** | **p** | **B Got Worse** | **B Got Better** | **F** | **p** | **B Got Worse** | **B Got Better** | **F** | **p** | **B Got Worse** | **B Got Better** |
| **Health and Quality of Life** |  |  |  |  |  |  |  |  |  |  |  |  |
| **Overall Health** | **26.09** | **<0.0001** | **-0.38** | **0.15** | **47.64** | **<0.0001** | **-0.28** | **0.29** | **6.23** | **0.0023** | **-0.12** | **0.17** |
| **Quality of Life** | **25.49** | **<0.0001** | **-1.00** | **0.48** | **23.05** | **<0.0001** | **-0.89** | **0.43** | **15.01** | **<0.0001** | **-0.82** | **0.46** |
| **Social Engagement** |  |  |  |  |  |  |  |  |  |  |  |  |
| **Community Participation** | **17.35** | **<0.0001** | **-0.20** | **0.52** | **31.44** | **<0.0001** | **-0.31** | **0.38** | **13.03** | **<0.0001** | **-0.35** | **0.35** |
| **Choices and Decisions** | 1.46 | 0.2348 | -0.15 | 0.09 | **13.63** | **<0.0001** | **-0.28** | **0.20** | **9.25** | **0.0001** | **-0.52** | **0.14** |
| **Self Determination** | **18.30** | **<0.0001** | **-0.38** | **0.11** | **18.44** | **<0.0001** | **-0.19** | **0.18** | **3.19** | **0.0431** | **-0.16** | **0.19** |
| **Employment** |  |  |  |  |  |  |  |  |  |  |  |  |
| **Job Satisfaction Survey** | **14.31** | **<0.0001** | **-0.45** | **0.19** | **15.16** | **<0.0001** | **-0.20** | **0.32** | 1.01 | 0.3698 | -0.20 | 0.09 |
| **Overall Employment Satisfaction** | **21.56** | **<0.0001** | **-0.64** | **0.62** | **22.25** | **<0.0001** | **-0.46** | **0.56** | **4.28** | **0.0183** | **-0.25** | **0.40** |
| **Mental Health** |  |  |  |  |  |  |  |  |  |  |  |  |
| **Anxiety** | **20.45** | **<0.0001** | **0.36** | **-0.20** | **25.26** | **<0.0001** | **0.32** | **-0.27** | **21.33** | **<0.0001** | **0.41** | **-0.19** |
| **Depression** | **21.98** | **<0.0001** | **0.30** | **-0.22** | **23.80** | **<0.0001** | **0.26** | **-0.23** | **18.61** | **<0.0001** | **0.32** | **-0.22** |
| **Autistic Burnout** | **25.71** | **<0.0001** | **0.39** | **-0.30** | **40.69** | **<0.0001** | **0.36** | **-0.52** | **16.99** | **<0.0001** | **0.31** | **-0.44** |
| **Flourishing** | **8.71** | **0.0002** | **-0.25** | **0.11** | **31.60** | **<0.0001** | **-0.33** | **0.29** | **5.91** | **0.0032** | **-0.30** | **0.25** |
| **Social Support** |  |  |  |  |  |  |  |  |  |  |  |  |
| **Social Support: Companionship** | **22.19** | **<0.0001** | **-0.45** | **0.46** | **23.40** | **<0.0001** | **-0.36** | **0.32** | **3.61** | **0.0287** | **-0.32** | **0.23** |
| **Social Support: Emotional Support** | **11.64** | **<0.0001** | **-0.37** | **0.30** | **21.47** | **<0.0001** | **-0.47** | **0.25** | **3.29** | **0.0390** | **-0.40** | **0.10** |
| **Social Support: Informational Support** | **10.05** | **<0.0001** | **-0.52** | **0.12** | **8.35** | **0.0003** | **-0.26** | **0.28** | **5.32** | **0.0055** | **-0.34** | **0.39** |
| **Social Support: Practical Support** | **7.54** | **0.0006** | **-0.54** | **0.37** | **12.49** | **<0.0001** | **-0.52** | **0.37** | 3.00 | 0.0519 | -0.26 | 0.23 |
| **Social Support: Isolation** | **18.87** | **<0.0001** | **-0.39** | **0.17** | **17.73** | **<0.0001** | **-0.29** | **0.19** | **6.49** | **0.0018** | **-0.28** | **0.14** |
| **Supports and Services** |  |  |  |  |  |  |  |  |  |  |  |  |
| **Barriers to Communication** | **3.60** | **0.0285** | **0.52** | **-0.53** | **5.66** | **0.0038** | **0.57** | **-0.30** | 2.22 | 0.1115 | 0.47 | -0.66 |
| **Satisfaction with Services: Support Staff** | 2.09 | 0.1309 | -0.78 | 0.52 | **5.00** | **0.0080** | **-0.65** | **0.18** | **4.28** | **0.0161** | **-0.42** | **0.03** |
| **Satisfaction with Services: Access** | 0.33 | 0.7174 | 0.27 | 0.21 | **9.13** | **0.0002** | **-0.41** | **0.70** | **3.57** | **0.0313** | **-0.48** | **0.11** |
| **Adequacy of Supports and Services** | 0.97 | 0.3815 | -0.19 | 0.10 | **9.00** | **0.0002** | **-0.27** | **0.38** | 0.87 | 0.4216 | -0.09 | 0.08 |
| **Healthcare Quality** |  |  |  |  |  |  |  |  |  |  |  |  |
| **Patient-Provider Communication** | **10.70** | **<0.0001** | **-0.58** | **0.53** | **12.18** | **<0.0001** | **-0.49** | **0.38** | **16.03** | **<0.0001** | **-0.61** | **0.47** |
| **Healthcare Self-Efficacy: Individual Level** | 2.90 | 0.0569 | -0.45 | 0.29 | 1.32 | 0.2695 | -0.13 | 0.29 | **4.02** | **0.0193** | **-0.36** | **0.65** |
| **Healthcare Self-Efficacy: Relationship-Dependent** | **4.77** | **0.0092** | **-0.66** | **0.26** | **3.82** | **0.0228** | **-0.16** | **0.62** | **12.37** | **<0.0001** | **-0.88** | **0.65** |
| **Barriers to Healthcare** | 2.36 | 0.0963 | 0.41 | -0.68 | 1.06 | 0.3471 | 0.47 | 0.13 | 1.17 | 0.3128 | 0.24 | -0.82 |
| **Unmet Healthcare Needs** | 2.75 | 0.0656 | 0.42 | -0.28 | **5.85** | **0.0032** | **0.37** | **-0.67** | 1.90 | 0.1520 | 0.39 | -0.13 |

None = does not need assistance with ADLs or IADLs; IADLs Only = needs assistance with instrumental activities of daily living but not basic self-care; ADLs = needs assistance with activities of daily living. F = F-statistic from OLS regression with cluster-robust standard errors. B (Got Worse) = mean difference in change score between participants who reported getting worse vs. those who stayed the same; B (Got Better) = same comparison for those who got better. Analyses used pooled longitudinal observations across two follow-up intervals; each participant contributes up to two observations per analysis. Bold values indicate statistical significance at p < 0.05. b Category absent in this subsample.

**Supplemental Table F3:**
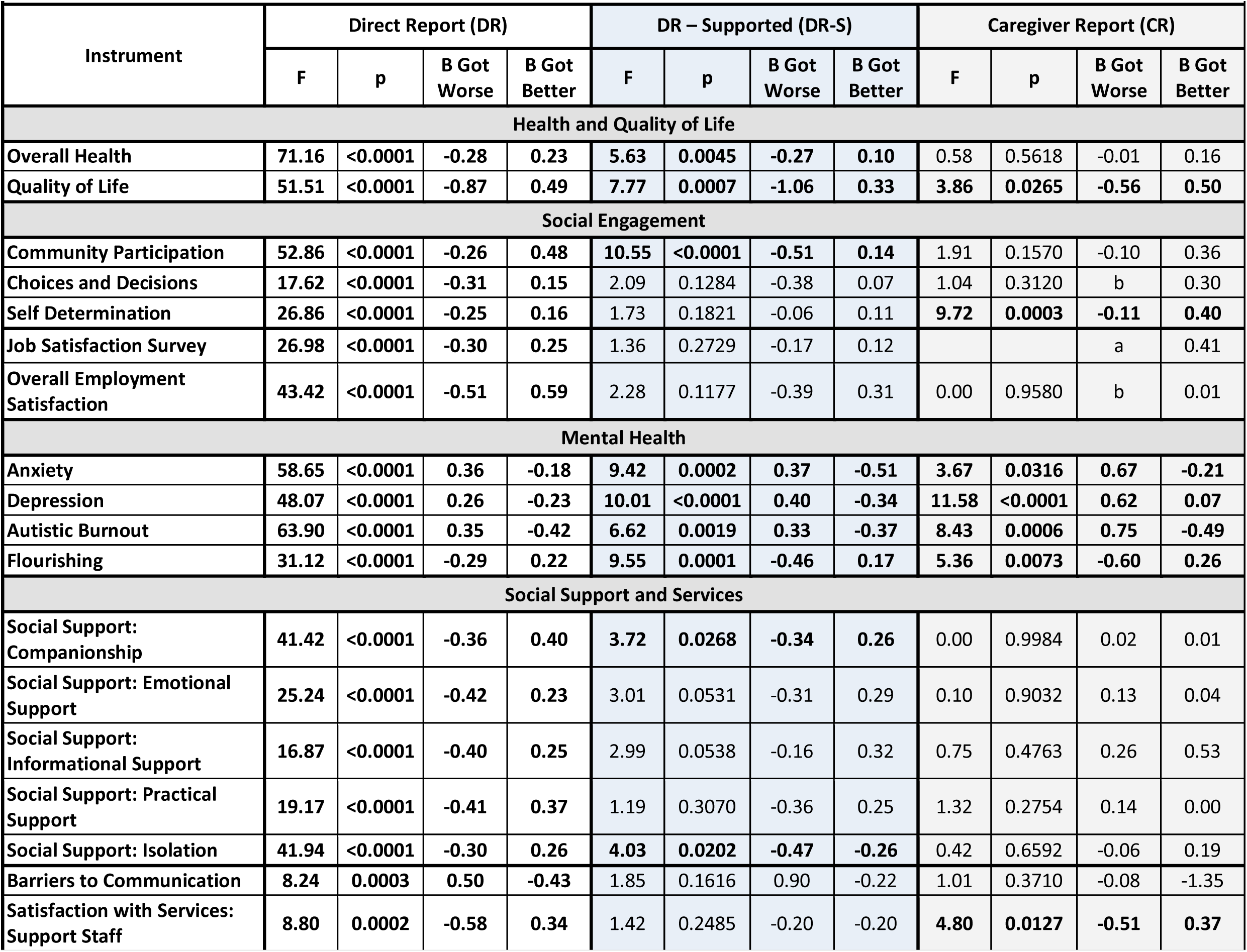
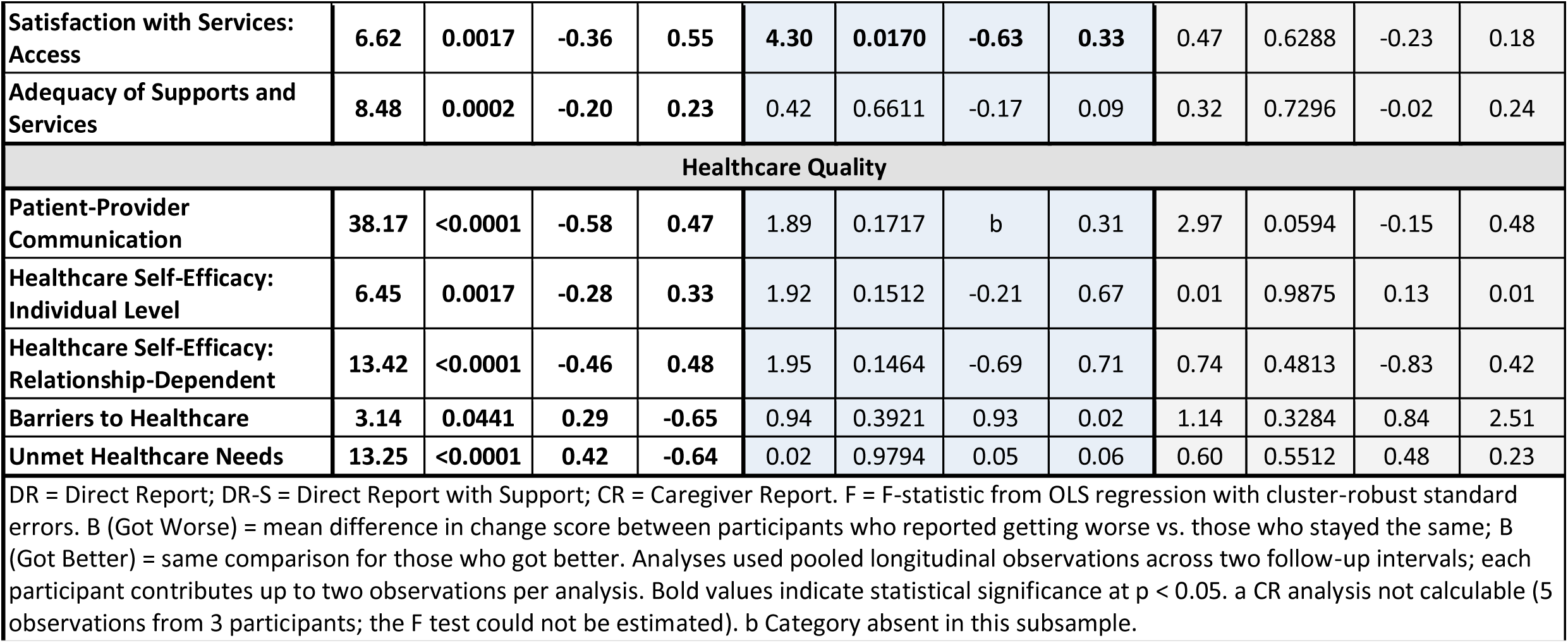
Responsiveness to Change by Reporter Type.

| **Instrument** | **Direct Report (DR)** |  |  |  | **DR – Supported (DR-S)** |  |  |  | **Caregiver Report (CR)** |  |  |  |
|---|---|---|---|---|---|---|---|---|---|---|---|---|
|  | **F** | **p** | **B Got Worse** | **B Got Better** | **F** | **p** | **B Got Worse** | **B Got Better** | **F** | **p** | **B Got Worse** | **B Got Better** |
| **Health and Quality of Life** |  |  |  |  |  |  |  |  |  |  |  |  |
| **Overall Health** | **71.16** | **<0.0001** | **-0.28** | **0.23** | **5.63** | **0.0045** | **-0.27** | **0.10** | 0.58 | 0.5618 | -0.01 | 0.16 |
| **Quality of Life** | **51.51** | **<0.0001** | **-0.87** | **0.49** | **7.77** | **0.0007** | **-1.06** | **0.33** | **3.86** | **0.0265** | **-0.56** | **0.50** |
| **Social Engagement** |  |  |  |  |  |  |  |  |  |  |  |  |
| **Community Participation** | **52.86** | **<0.0001** | **-0.26** | **0.48** | **10.55** | **<0.0001** | **-0.51** | **0.14** | 1.91 | 0.1570 | -0.10 | 0.36 |
| **Choices and Decisions** | **17.62** | **<0.0001** | **-0.31** | **0.15** | 2.09 | 0.1284 | -0.38 | 0.07 | 1.04 | 0.3120 | b | 0.30 |
| **Self Determination** | **26.86** | **<0.0001** | **-0.25** | **0.16** | 1.73 | 0.1821 | -0.06 | 0.11 | **9.72** | **0.0003** | **-0.11** | **0.40** |
| **Job Satisfaction Survey** | **26.98** | **<0.0001** | **-0.30** | **0.25** | 1.36 | 0.2729 | -0.17 | 0.12 |  |  | a | 0.41 |
| **Overall Employment Satisfaction** | **43.42** | **<0.0001** | **-0.51** | **0.59** | 2.28 | 0.1177 | -0.39 | 0.31 | 0.00 | 0.9580 | b | 0.01 |
| **Mental Health** |  |  |  |  |  |  |  |  |  |  |  |  |
| **Anxiety** | **58.65** | **<0.0001** | **0.36** | **-0.18** | **9.42** | **0.0002** | **0.37** | **-0.51** | **3.67** | **0.0316** | **0.67** | **-0.21** |
| **Depression** | **48.07** | **<0.0001** | **0.26** | **-0.23** | **10.01** | **<0.0001** | **0.40** | **-0.34** | **11.58** | **<0.0001** | **0.62** | **0.07** |
| **Autistic Burnout** | **63.90** | **<0.0001** | **0.35** | **-0.42** | **6.62** | **0.0019** | **0.33** | **-0.37** | **8.43** | **0.0006** | **0.75** | **-0.49** |
| **Flourishing** | **31.12** | **<0.0001** | **-0.29** | **0.22** | **9.55** | **0.0001** | **-0.46** | **0.17** | **5.36** | **0.0073** | **-0.60** | **0.26** |
| **Social Support and Services** |  |  |  |  |  |  |  |  |  |  |  |  |
| **Social Support: Companionship** | **41.42** | **<0.0001** | **-0.36** | **0.40** | **3.72** | **0.0268** | **-0.34** | **0.26** | 0.00 | 0.9984 | 0.02 | 0.01 |
| **Social Support: Emotional Support** | **25.24** | **<0.0001** | **-0.42** | **0.23** | 3.01 | 0.0531 | -0.31 | 0.29 | 0.10 | 0.9032 | 0.13 | 0.04 |
| **Social Support: Informational Support** | **16.87** | **<0.0001** | **-0.40** | **0.25** | 2.99 | 0.0538 | -0.16 | 0.32 | 0.75 | 0.4763 | 0.26 | 0.53 |
| **Social Support: Practical Support** | **19.17** | **<0.0001** | **-0.41** | **0.37** | 1.19 | 0.3070 | -0.36 | 0.25 | 1.32 | 0.2754 | 0.14 | 0.00 |
| **Social Support: Isolation** | **41.94** | **<0.0001** | **-0.30** | **0.26** | **4.03** | **0.0202** | **-0.47** | **-0.26** | 0.42 | 0.6592 | -0.06 | 0.19 |
| **Barriers to Communication** | **8.24** | **0.0003** | **0.50** | **-0.43** | 1.85 | 0.1616 | 0.90 | -0.22 | 1.01 | 0.3710 | -0.08 | -1.35 |
| **Satisfaction with Services: Support Staff** | **8.80** | **0.0002** | **-0.58** | **0.34** | 1.42 | 0.2485 | -0.20 | -0.20 | **4.80** | **0.0127** | **-0.51** | **0.37** |
| **Satisfaction with Services: Access** | **6.62** | **0.0017** | **-0.36** | **0.55** | **4.30** | **0.0170** | **-0.63** | **0.33** | 0.47 | 0.6288 | -0.23 | 0.18 |
| **Adequacy of Supports and Services** | **8.48** | **0.0002** | **-0.20** | **0.23** | 0.42 | 0.6611 | -0.17 | 0.09 | 0.32 | 0.7296 | -0.02 | 0.24 |
| **Healthcare Quality** |  |  |  |  |  |  |  |  |  |  |  |  |
| **Patient-Provider Communication** | **38.17** | **<0.0001** | **-0.58** | **0.47** | 1.89 | 0.1717 | b | 0.31 | 2.97 | 0.0594 | -0.15 | 0.48 |
| **Healthcare Self-Efficacy: Individual Level** | **6.45** | **0.0017** | **-0.28** | **0.33** | 1.92 | 0.1512 | -0.21 | 0.67 | 0.01 | 0.9875 | 0.13 | 0.01 |
| **Healthcare Self-Efficacy: Relationship-Dependent** | **13.42** | **<0.0001** | **-0.46** | **0.48** | 1.95 | 0.1464 | -0.69 | 0.71 | 0.74 | 0.4813 | -0.83 | 0.42 |
| **Barriers to Healthcare** | **3.14** | **0.0441** | **0.29** | **-0.65** | 0.94 | 0.3921 | 0.93 | 0.02 | 1.14 | 0.3284 | 0.84 | 2.51 |
| **Unmet Healthcare Needs** | **13.25** | **<0.0001** | **0.42** | **-0.64** | 0.02 | 0.9794 | 0.05 | 0.06 | 0.60 | 0.5512 | 0.48 | 0.23 |

DR = Direct Report; DR-S = Direct Report with Support; CR = Caregiver Report. F = F-statistic from OLS regression with cluster-robust standard errors. B (Got Worse) = mean difference in change score between participants who reported getting worse vs. those who stayed the same; B (Got Better) = same comparison for those who got better. Analyses used pooled longitudinal observations across two follow-up intervals; each participant contributes up to two observations per analysis. Bold values indicate statistical significance at p < 0.05. a CR analysis not calculable (5 observations from 3 participants; the F test could not be estimated). b Category absent in this subsample.

**Supplemental Table G1:** Distributional Properties of Outcome Variable Scores.

| Instrument | N | Mean | SD | Min | Max | Skewness | Kurtosis | Note |
|---|---|---|---|---|---|---|---|---|
| **Health and Quality of Life** |  |  |  |  |  |  |  |  |
| AASPIRE Quality of Life Item | 862 | 6.11 | 1.99 | 0.00 | 10.00 | -0.529 | 2.964 | † |
| AASPIRE Overall Health Scale | 870 | 2.97 | 0.85 | 1.00 | 5.00 | 0.112 | 2.521 |  |
| Overall Health: Interference | 869 | 2.37 | 0.79 | 1.00 | 5.00 | 0.482 | 2.777 |  |
| PHQ-9 (Depression) | 836 | 1.00 | 0.74 | 0.00 | 3.00 | 0.595 | 2.555 |  |
| GAD-7 (Anxiety) | 836 | 1.31 | 0.86 | 0.00 | 3.00 | 0.287 | 2.022 |  |
| Flourishing Scale | 836 | 3.60 | 0.79 | 1.00 | 5.00 | -0.555 | 3.229 |  |
| Autistic Burnout Scale – Revised | 835 | 2.91 | 1.03 | 1.00 | 5.00 | -0.088 | 2.212 |  |
| **Social Engagement** |  |  |  |  |  |  |  |  |
| Community Participation Scale | 845 | 3.03 | 0.85 | 1.00 | 5.00 | 0.025 | 2.324 |  |
| Self-Determination Inventory | 830 | 3.79 | 0.65 | 1.19 | 5.00 | -0.351 | 3.237 |  |
| Choices and Decisions Scale | 839 | 4.22 | 0.73 | 1.00 | 5.00 | -1.142 | 4.385 |  |
| Job Satisfaction Survey | 331 | 3.57 | 0.66 | 1.64 | 4.95 | -0.379 | 2.857 |  |
| Overall Employment Satisfaction | 413 | 3.46 | 0.87 | 1.00 | 5.00 | -0.461 | 2.991 |  |
| **Supports and Services** |  |  |  |  |  |  |  |  |
| Social Support Scale | 859 | 3.34 | 0.77 | 1.05 | 5.00 | -0.273 | 2.710 |  |
| Social Support: Companionship | 857 | 3.35 | 0.97 | 1.00 | 5.00 | -0.249 | 2.429 |  |
| Social Support: Emotional Support | 856 | 3.56 | 0.94 | 1.00 | 5.00 | -0.408 | 2.628 |  |
| Social Support: Informational Support | 857 | 3.53 | 0.93 | 1.00 | 5.00 | -0.323 | 2.619 |  |
| Social Support: Practical Support | 859 | 3.46 | 1.15 | 1.00 | 5.00 | -0.327 | 2.102 |  |
| Social Support: Lack of Isolation | 857 | 2.75 | 0.95 | 1.00 | 5.00 | 0.253 | 2.620 |  |
| Barriers to Communication Scale | 870 | 72.93 | 54.29 | 0.00 | 375.00 | 1.615 | 6.498 |  |
| Satisfaction with Disability Services | 302 | 3.81 | 0.84 | 1.00 | 5.00 | -0.758 | 3.268 |  |
| Adequacy of Supports and Information | 798 | 3.59 | 0.88 | 1.00 | 5.00 | -0.552 | 2.831 |  |
| **Healthcare Quality** |  |  |  |  |  |  |  |  |
| Unmet Healthcare Needs Checklist | 794 | 1.59 | 2.05 | 0.00 | 11.00 | 1.592 | 5.364 | ‡ |
| Barriers to Healthcare Checklist | 794 | 3.53 | 3.69 | 0.00 | 17.00 | 1.164 | 3.857 | † |
| Patient-Provider Communication Scale | 793 | 3.96 | 0.88 | 1.00 | 5.00 | -0.877 | 3.642 |  |
| HC Self-Efficacy: Individual Level | 792 | 7.28 | 1.92 | 1.00 | 10.00 | -0.725 | 2.960 |  |
| HC Self-Efficacy: Relationship-Dependent | 792 | 7.36 | 1.92 | 1.00 | 10.00 | -0.711 | 3.022 |  |
| Multi-item Likert scale range |  |  |  |  |  | \|0.03\|–\|1.14\| | 2.02–4.39 |  |

† Skewness or kurtosis at the upper margin of the range observed for multi-item Likert-scale scores but not substantially outside it.
‡ Skewness and kurtosis notably exceed the range observed for multi-item Likert-scale scores, consistent with a right-skewed count distribution with many zero values.
Note: The shaded reference row shows the range of skewness (absolute value) and kurtosis observed across the 23 multi-item Likert-scale mean scores (excluding the Quality of Life single-item measure and the two checklist-based counts). The Barriers to Communication Scale uses a product score (frequency × impact) rather than a simple mean; its SD and range are not directly comparable to other instruments. N varies across instruments due to skip patterns (e.g., employment scales administered only to employed participants; Satisfaction with DD Services administered only to those receiving services).

### Supplement G2: Sensitivity Analyses Related to Non-Parametric Data

Supplemental Table G1 presents the skewness, kurtosis, standard deviation, and score range for all 26 outcome variables. We examined these statistics to assess the distributional comparability of the Quality of Life single-item measure, the Unmet Healthcare Needs Checklist, and the Barriers to Healthcare Checklist to the multi-item Likert-scale scores, given that single-item and checklist-derived count scores may not meet the distributional assumptions underlying intraclass correlation coefficients (ICCs) and regression-based responsiveness analyses.

The Quality of Life item (skewness = −0.53, kurtosis = 2.96, SD = 1.99) and Barriers to Healthcare Checklist (skewness = 1.16, kurtosis = 3.86, SD = 3.69) showed skewness and kurtosis within or near the range observed for the 23 multi-item Likert-scale mean scores (|skewness| range: 0.03–1.14; kurtosis range: 2.02–4.39), supporting the use of ICC and continuous-variable methods for these instruments. The Unmet Healthcare Needs Checklist showed greater departure from this range (skewness = 1.59, kurtosis = 5.36), consistent with its right-skewed count distribution and high proportion of zero scores.

We also examined distributional properties of the change scores used in the responsiveness analyses. Change score distributions showed elevated kurtosis across all 26 outcome variables (range: 4.18–9.65), which is expected for longitudinal change data in which most participants show little change between waves. The change score distributions of the Quality of Life item (skewness = −0.06, kurtosis = 5.37), Unmet Healthcare Needs Checklist (skewness = −0.48, kurtosis = 9.40), and Barriers to Healthcare Checklist (skewness = −0.32, kurtosis = 8.57) were not meaningfully more departing from normality than those of several multi-item Likert-scale change scores (e.g., Flourishing kurtosis = 8.08, Choices and Decisions kurtosis = 8.30, Barriers to Communication kurtosis = 9.65). The use of cluster-robust standard errors in the regression-based responsiveness analyses provides valid inference under non-normal residual distributions without requiring normality assumptions.

As sensitivity analyses, we calculated Spearman’s rho as a non-parametric test-retest reliability coefficient for the three flagged instruments (n = 129–132). Results were fully consistent with the ICC-based estimates: Quality of Life ρ = 0.85, Barriers to Healthcare ρ = 0.79, Unmet Healthcare Needs ρ = 0.73 (all p < 0.0001), confirming adequate two-week test-retest reliability under non-parametric assumptions.

We also replicated the responsiveness analyses for both checklist instruments using Wilcoxon signed-rank tests with rank-biserial correlation as a non-parametric effect size. For the Unmet Healthcare Needs Checklist, results were fully consistent with the primary regression-based findings: participants reporting worsening healthcare needs showed significantly increased scores (z = 3.06, p = 0.002, r = 0.25), those reporting improvement showed significantly decreased scores (z = −2.67, p = 0.008, r = −0.37), and those reporting no change showed no significant difference from zero (z = −1.69, p = 0.09, r = −0.08). For the Barriers to Healthcare Checklist, no consistent pattern of significant change emerged across self-perceived change categories (Got Worse: z = 1.29, p = 0.20, r = 0.11; Stayed Same: z = −1.50, p = 0.13, r = −0.06; Got Better: z = −1.97, p = 0.049, r = −0.26), replicating the null responsiveness finding from the primary analysis. This suggests the null finding reflects the smaller magnitude of change in healthcare barriers in an observational cohort rather than a distributional artifact — the instrument has previously demonstrated responsiveness in intervention contexts.

